# Adaptive Responses to PARP Inhibition Predict Response to Olaparib and Durvalumab: Multi-omic Analysis of Serial Biopsies in the AMTEC Trial

**DOI:** 10.1101/2024.08.29.24312245

**Authors:** Zahi I. Mitri, Allison L. Creason, Jayne M. Stommel, Tugba Y. Ozmen, Matthew J. Rames, Furkan Ozmen, Daniel Bottomly, Boyoung Jeong, Jeong Youn Lim, Shamilene Sivagnanam, Konjit Betre, Jinho Lee, Marilyne Labrie, Lisa M. Coussens, Christopher L. Corless, Shannon K. McWeeney, Gordon B. Mills

## Abstract

In syngeneic murine breast cancer models, poly(ADP-ribose) polymerase inhibitor (PARPi) and anti-PD-L1 combinations induce deep, sustained responses independent of *BRCA1* or *BRCA2* mutation (BRCAm) status. We therefore investigated this combination in the AMTEC clinical trial, in which a one-month olaparib run-in was followed by combined olaparib and durvalumab in participants with non-BRCAm metastatic triple negative breast cancer. To characterize adaptive responses to olaparib monotherapy, paired biopsies taken before and during the PARPi lead-in were deeply characterized by DNA, RNA, and protein multi-omic analyses, including spatially resolved single-cell proteomics for tumor and immune contexture. We identified multiple potential tumor-intrinsic and microenvironmental biomarkers from pre-treatment and on-olaparib biopsies that robustly predicted participant response to combined olaparib and durvalumab. Notably, the on-olaparib biopsy provided the greatest information content, suggesting that early adaptations of malignant and immune cells to PARPi can serve as a predictor of potential benefit from combined PARPi and anti-PD-L1 therapy.

## INTRODUCTION

Triple negative breast cancer (TNBC), comprising 15-20% of all breast cancers, predominantly affects younger women and black and Hispanic ethnic groups. Compared to other breast cancer subtypes, TNBC is characterized by aggressive features, a higher propensity for metastasis, and an overall worse prognosis^1^. Despite advancements in understanding TNBC biology, outcomes for metastatic TNBC (mTNBC) remain poor, with an overall survival of 1-2 years with standard therapies^2–4^.

The DNA damage response (DDR) pathway is frequently dysregulated in TNBC, with nearly half of all cases exhibiting aberrations. *BRCA1* or *BRCA2* mutations (BRCAm) are observed in about 15% of TNBC, while another 45% of TNBC with genomic instability are non-BRCAm, suggesting that other aberrations can lead to defects in this pathway^1,5–7^. DDR dysregulation renders TNBC sensitive to DNA damaging therapies such as chemotherapy, radiation therapy, and more recently, poly(ADP-ribose) polymerase (PARP) inhibitors (PARPi). PARPi are of particular interest in mTNBC because of the high frequency of defects in homologous recombination (HR) repair, the high-fidelity repair pathway for double-stranded DNA breaks^8^.

In mTNBC, the PARPi olaparib and talazoparib, FDA-approved for patients with germline BRCAm, improve clinical outcomes primarily by exploiting synthetic lethality with defective HR (HRD). However, even in this population, responses to PARPi monotherapy are often short-lived, with most patients eventually experiencing disease progression^9,10^. Moreover, PARPi monotherapy has limited activity in non-BRCAm tumors, which constitute the majority of mTNBC^11^, with the VIOLETTE trial showing minimal benefit in these patients^12^. Preclinical and early clinical data suggest that PARPi combination therapies are safe and more effective in non-BRCAm TNBC, highlighting potential opportunities for these treatments for this population^13–15^.

Immune checkpoint blockade (ICB) has improved outcomes for various tumor types, especially those with high tumor mutational burden (TMB) and neoantigenic propensity, which can bolster anti-tumor immunity^16,17^. In mTNBC, the combination of the programmed cell death 1 (PD-1) inhibitor pembrolizumab and chemotherapy is FDA-approved for patients with tumors expressing programmed cell death ligand 1 (PD-L1). However, this indication excludes the majority of mTNBCs, which are frequently PD-L1 negative, and clinical benefit is often brief even in PD-L1 positive patients^18^. Additionally, TMB in TNBC is usually below the generally accepted 10 mutations/Mb that predicts ICB efficacy in other cancer types^19^.

Extensive preclinical data demonstrate that PARPi induce adaptive responses in malignant cells and the tumor ecosystem, including immune cells, in models of HR-deficient and HR-competent tumors^20–24^. Similar adaptive changes have been observed in human tumors, such as breast cancer, under therapeutic stress induced by PARPi^14,15,25,26^. Although adaptive responses to PARP inhibition vary among patients, they appear to involve a limited set of stress response pathways, suggesting that identification of patient-specific adaptive responses is feasible. Targeting these adaptive responses – for example, by increasing PARPi-induced stress, blocking protective responses, or capitalizing on the rewiring of malignant cells and the immune microenvironment^27^ – has the potential to improve responses to PARPi, including in non-BRCAm mTNBC. The identification of the specific adaptive response to target may require collecting tumor biopsies before and during PARPi monotherapy, then cataloguing drug-induced changes that could be exploited by a combined treatment regimen.

Combining PARPi with ICB is a promising therapeutic approach to leverage PARPi-induced adaptive changes in immune signaling. PARP inhibition enhances antigen presentation, recruits cytotoxic T lymphocytes, and activates the stimulator of interferon genes (STING) pathway^21,22,28^, all of which could increase the efficacy of ICB. Notably, PARPi can activate the STING pathway in non-BRCAm syngeneic cancer models that are DNA repair competent^23^, potentially leading to an infiltration of adaptive immune cells that would recognize neoantigens generated through low fidelity DNA repair. Indeed, clinical trials combining ICB and PARPi showed activity in both BRCAm and non-BRCAm mTNBC, although response rates are lower in the wild-type group^14,29–31^.

Building on these findings, we initiated the Adaptive Molecular Therapy of Evolving Cancers (AMTEC) trial to evaluate the efficacy of combining the PARPi olaparib with the PD-L1 inhibitor durvalumab in non-BRCAm mTNBC. This trial includes an olaparib monotherapy phase followed by a combined olaparib and durvalumab phase, with biopsies collected at baseline and during olaparib monotherapy. Both biopsies were analyzed with a suite of multi-omic assays to provide an in-depth examination of changes in malignant cells and the tumor ecosystem in response to PARPi-mediated therapeutic stress. Using this approach, the AMTEC trial was designed to identify adaptive changes induced by PARPi monotherapy that could serve as biomarkers for selecting patients likely to benefit from PARPi/ICB combination therapy in follow-on trials. An additional goal was to discover resistance mechanisms to combined PARPi/ICB, which could be targeted with alternative PARPi combinations. AMTEC closed to enrollment in 2024 with n=24 participants enrolled. Here we report on the final clinical and initial translational results for the AMTEC trial.

## RESULTS

AMTEC is a single-arm, open-label phase II trial assessing the efficacy of olaparib combined with durvalumab in participants with non-BRCAm mTNBC (Fig. 1a). Eligible participants undergo a pre-treatment biopsy upon enrollment, followed by a 4-week olaparib treatment. At the end of this olaparib lead-in, all participants undergo a second biopsy on olaparib, and then durvalumab is added to the treatment regimen. The combination of olaparib and durvalumab continues until progression or toxicity-required termination.

**Figure 1.**
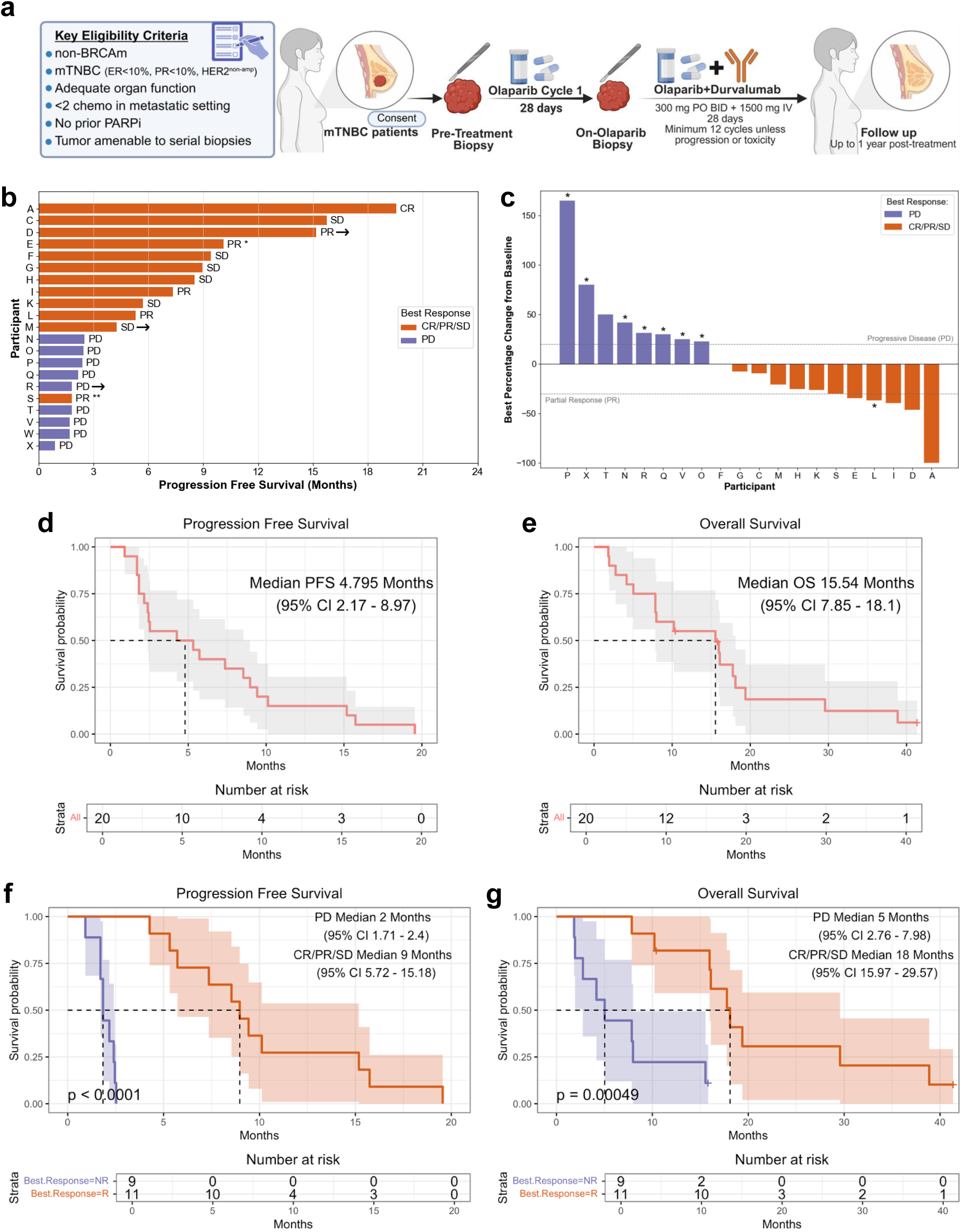
Overview of the AMTEC trial and clinical outcomes. **a)** *Trial design.* Schematic of the AMTEC trial showing key eligibility criteria, treatment schedule, biopsy timing, treatment discontinuation, and post-treatment follow-up. Created in BioRender. Yildiran Ozmen, T. (2025) https://BioRender.com/vjju338 **b)** *Progression free survival (PFS).* Swimmer plot for 21 efficacy-evaluable participants (see Extended Data Fig. 1a). PFS measured from the start of olaparib treatment until the end of olaparib plus durvalumab. Best response by RECIST v1.1^32^ is adjacent to each treatment duration bar. Orange bars: participants with Complete Response (CR), Partial Response (PR), or Stable Disease (SD); purple bars: Progressive Disease (PD). * = passed away from a trial-unrelated accident; ** = voluntarily withdrew from the study. Arrows = alive at time of data cutoff. **c)** *Best percentage change from baseline.* Waterfall plot showing best change in the sum of all target lesions for 20 efficacy-evaluable participants. Lesions from Participant W were not evaluable. Dashed lines indicate RECIST thresholds for PD (+20%) and PR (−30%). * = new lesions. **d-g)** *Kaplan-Meier analyses.* PFS and Overall Survival (OS) for 20 survival-evaluable participants (see Extended Data Fig. 1a). Shaded areas indicate 95% confidence intervals (CI); median time shown by dashed line. PFS measured from start of olaparib until end of olaparib plus durvalumab. OS measured from olaparib start until death or data cutoff; right-censoring denoted by “+”. **d)** PFS, all. **e)** OS, all. **f)** PFS, by RECIST best response. Non-responders (NR; purple) = PD. Responders (R; orange) = CR/PR/SD. P-values from log-rank tests. **g)** OS, by RECIST best response.

The primary endpoint of the study is the overall response rate (ORR) for olaparib plus durvalumab. Secondary endpoints include clinical benefit rate (CBR), duration of response, progression-free survival (PFS), and overall survival (OS), as well as safety and tolerability of the treatment.

### Participant and Disease Characteristics

Twenty-four participants with biopsy-proven mTNBC were enrolled in the AMTEC trial between January 2019 and May 2024. Participant characteristics are summarized in Table 1. All participants had an ECOG performance status of 0 or 1. Similar numbers of participants had 0, 1, or 2 lines of prior therapy in the metastatic setting, including seven participants who had received platinum-based regimens. Seven participants had controlled brain metastases at the time of enrollment.

**Table 1.** Summary of clinical features of AMTEC trial participants at baseline.

|  |  | **N = 24 (%)** |
| --- | --- | --- |
| **Median age (range)** | 54.0 (25.3 – 78.9) |  |
| **ECOG performance status** | 0 | 15 (62.5%) |
|  | 1 | 9 (37.5%) |
| **Race** | Caucasian | 21 (87.5%) |
|  | African American | 1 (4.2%) |
|  | Undisclosed | 2 (8.3%) |
| **Menopausal status** | Pre-menopausal | 8 (33.3%) |
|  | Post-menopausal | 16 (66.7%) |
| **Lines of therapy in metastatic setting** | 0 | 8 (33.3%) |
|  | 1 | 10 (41.7%) |
|  | 2 | 6 (25.0%) |
| **Prior platinum chemotherapy in metastatic setting** | Yes | 7 (29.2%) |
|  | No | 17 (70.8%) |
| **History of brain metastases** | Yes | 7 (29.2%) |
|  | No | 17 (70.8%) |

Of the 24 enrolled participants, three participants were excluded, resulting in a set of 21 participants in the primary endpoint ORR analysis (Extended Data Fig. 1a). An additional participant voluntarily withdrew during olaparib/durvalumab cycle 3 per personal preference. This participant (“S”; Fig. 1b) achieved a RECIST best response of partial response (PR)^32^ on first restaging and was included in the ORR analysis but omitted from survival analyses, resulting in a total of 20 participants evaluable for the survival efficacy endpoints (PFS and OS).

### Efficacy Endpoints

Among the 21 participants who received the combination therapy and were evaluable for the primary ORR efficacy endpoint, there were 5 PRs and 1 complete response (CR), for an ORR of 29% (Fig. 1b and Extended Data Table 1a). Four additional participants had stable disease (SD) ≥6 months, for a CBR of 48%. Two participants had SD as best response on first restaging scan but progressed prior to the 6-month timeline. Of note, two participants with PR (Participant D) and CR (Participant A) as best response on study were off therapy for 26 and 19 months respectively (Extended Data Fig. 1b). Participant D remains without evidence of disease progression and has not required additional systemic therapy, whereas Participant A eventually recurred and died of progressive metastatic disease. These outcomes mirror findings from our prior three-patient pilot study with the same design as AMTEC, which included one prolonged PR, one SD, and one PD^14^.

The best percentage change from baseline for the sum of all target lesion diameters was calculated for each efficacy-evaluable participant (Fig. 1c). Among the six participants with SD as best response, percentage changes ranged from 0% to −27%. The four participants with PR as the best response showed reductions from −30% to −47%, and the single participant with CR exhibited a −100% reduction. In contrast, the five participants with progressive disease (PD) demonstrated increases ranging from +23% to +165%. Participant W lacked target lesion size measurements at the time of progression, and percent change could not be determined.

The median PFS for the entire cohort was 4.8 months (95% CI 2.17 – 8.97 months), and median OS was 15.5 months (95% CI 7.85 – 18.1 months; Fig. 1d,e). Participants with a best response of SD vs. PR did not show statistically significant differences in PFS or OS (PFS for PR vs. SD, p=0.97; OS, p=0.68). Therefore, for subsequent analyses, cases with SD, PR, or CR (n= 12) were grouped as participants who derived clinical benefit from the study therapy (“Responders”; R). These were compared to participants with PD as best response (n=9), who were considered to have primary resistance to the study therapy (“Non-Responders”; NR).

Participants with CR, PR, or SD as RECIST best response had significantly better survival outcomes than those with PD. Specifically, participants with CR/PR/SD had improved PFS (9 vs. 2 months, p<0.0001) and OS (18 vs. 5 months, p=0.00049) compared to PD patients (Fig. 1f,g). The marked survival differences between response groups justified efforts to uncover molecular markers that differentiate responders from non-responders and predict clinical benefit, with the potential to guide biomarker evaluation in subsequent trials.

Clinical outcomes were not associated with the total number of prior therapies (p=0.8, R vs. NR) or with prior platinum treatment in the metastatic setting (PFS, p=0.85; OS, p=0.26). Among participants with a history of CNS metastases, CNS progression occurred concurrently with the progression of extra-cranial systemic disease.

### Safety Results

All 24 participants who received at least one dose of study-directed therapy were evaluable for safety endpoints (Extended Data Table 1a,b). Most treatment-related adverse events (AEs) were mild to moderate (Grade 1-2 = 190/210; 90%). Thirteen participants (54%) experienced severe AEs (Grade 3), with 20/41 (49%) considered as possibly related to the study treatment. No Grade 4 or 5 AEs attributed to study interventions or treatment were noted.

Among the Grade 3 AEs, two consisted of immune-related elevated liver enzymes attributed to durvalumab, which resolved following dose hold and supportive medications (Extended Data Table 1b). The most common likely olaparib-related Grade 3 AE was anemia in 6/24 participants (25%), which resolved with a dose hold and adjustment per protocol. The two Grade 5 AEs included a motor vehicle accident (“Investigations – other*”) and respiratory failure, both unrelated to treatment.

### Clinical and Exploratory Research Analytics

To identify patients likely to benefit and to uncover targetable mechanisms of response and resistance, we conducted a comprehensive exploratory analysis of tumor-intrinsic and microenvironmental features in both the pre-treatment and on-olaparib tumor biopsies. This included a focus on signatures that emerged as adaptive responses to the olaparib monotherapy lead-in. In addition to the 21 participants treated on the AMTEC trial, we included three mTNBC participants from our previously reported pilot study who received the same sequence of biopsies and therapy as in AMTEC^14^. This approach serves to increase our power to identify potential biomarkers predictive of response to the olaparib and durvalumab combination (n=24).

Given the relatively small sample size and the absence of an independent test set with comparable study design and multi-omic profiling, we implemented a conservative analytical approach for the high-dimensional omics data (RNAseq, RPPA, mIHC, DSP). We employed the Boruta algorithm for feature selection, which incorporates Bonferroni correction to control for false discovery rate in multiple hypothesis testing. Predictive performance was then validated using F1 scores calculated across iterative stratified train-test splits, ensuring a balanced precision-recall trade-off and minimizing the risks of artifacts from feature selection (see Methods for details).

The clinical and exploratory analytical methods included: 1) clinical immunohistochemistry (IHC); 2) *GeneTrails®* DNA sequencing of 225 cancer-relevant genes; 3) whole exome DNA sequencing (WES); 4) bulk transcriptomic sequencing (RNAseq) for gene rearrangements and expression levels; 5) NanoString GeoMx*®* Digital Spatial Profiling (DSP) of 46 proteins and phospho-proteins; 6) reverse phase protein arrays (RPPA) of 480 proteins, including post-translational modifications; 7) RAD51 foci analysis to measure homologous recombination deficiency, and 8) multiplexed immunohistochemistry (mIHC) using 23 antibodies to quantify infiltrating immune cells and their functional states. Due to sample availability and assay-specific technical or biological requirements, such as minimum tumor content and tissue adherence, not all assays were performed on every sample.

Extended Data Fig. 1c and Extended Data Tables 1c,d summarize the results of these clinical and exploratory research analyses, discussed in detail below. Extended Data Fig. 1c shows clinical metadata and assay results for each sample, noting statistically significant associations with best response (R), PFS (P), OS (O), as well as two paired-biopsy analyses. “B” denotes within-group paired comparisons, showing whether samples from responders and non-responders changed from pre-treatment to on-olaparib when each group is analyzed separately. “D” denotes between-group differences in olaparib-induced changes, testing whether the magnitude of change from pre-treatment to on-olaparib differs between responders and non-responders. Extended Data Tables 1c,d list the features that were statistically significant predictors of response to the olaparib-durvalumab combination, including candidate biomarkers selected by Boruta with high F1 scores within each data type (see Methods for details of the statistical analysis).

### Malignant Cell-Intrinsic Analytes

The following measurements did not show statistically significant correlations with participant outcomes:1) mutations in *TP53*, the HRD pathway, or the MAPK pathway; 2) TMB (no sample exceeded 10 mutations/Mb); 3) percentage of malignant cells expressing PD-L1 or Ki-67 by clinical IHC; 4) biopsy site; or 5) tumor content, although this tended to be higher in participants with PD.

The most frequently mutated gene was *TP53*, observed in 22 out of 24 participants (92%; Extended Data Fig. 1c). Of the genes known to confer sensitivity to PARPi^33,34^, pilot study Participant B was PR and had an unusual germline deletion of exons 13-15 of *BRCA1* that was not detected on initial screening. In contrast, Participant V was PD with somatic *BRCA1* copy loss (0.5 copies). Notably, this participant also had copy loss of *PALB2* (0.35 copies), homozygous deletions of *ATM* and *RAD51,* and 15-copy gain of AKT2, which may have contributed to their poor response. No other deleterious mutations in known PARPi-sensitizing genes *BRCA1*, *BRCA2, BARD1, PALB2, RAD51C, or RAD51D* were noted. Alterations in other genes implicated in homologous recombination (HR) included germline *ATM* mutations in Participants C (SD; p.R3008C) and R (PD; p.S99_I105dup) and a *CDK12* mutation in Participant K (SD; p.L123fs*3). Regarding mutations possibly related to durvalumab response, responding Participants I and H had deleterious *B2M* mutations (splice site and p.C45_N62del), and non-responding Participant P had low frequency compound *JAK1* mutations (D1003Y and A1005V) detected only in the on-olaparib biopsy.

Defects in HR are well-established predictive biomarkers for PARPi response, even in the absence of *BRCA1/2* mutations^35–38^. To assess HR deficiency in our predominantly non-BRCAm samples, we used an immunofluorescence-based assay that we developed previously to distinguish HR-competent (HRC) from HR-deficient (HRD) cells^39^. Cells in G2 phase were evaluated for the presence of functional HR repair complexes (RAD51 foci) and double-strand DNA breaks (γH2AX), then samples were categorized as HRC, HRD, or Intermediate (with a mixture of HRC and HRD cells) based on the ratio of HRD to HRC nuclei (Extended Data Fig. 2a-c). Strikingly, all participants with tumors classified as HRD (including participant (B) from the pilot study with an unusual *BRCA1* mutation) achieved a best response of CR, PR or SD, including those that shifted from Intermediate to HRD in the second biopsy (Extended Data Fig1c, 2d). In contrast, no samples from participants with PD were classified as HRD in either biopsy.

Genomic scarring is a common indicator of HRD due to its strong association with *BRCA1/2* mutations and other DNA damage mechanisms^40^. Given its association with HRD, we assessed the predictive value of the MutSig3 signature in the WES from the pre-therapy biopsy. MutSig3 was significantly associated with OS (p=0.03; Fig. 2a) but not with best response or PFS (note only limited samples had WES). Interestingly, there was no clear correspondence between the HRD/HRC ratio and the MutSig3 signature in samples with both assays (Extended Data Fig. 1c). This discrepancy may reflect the different aspects of DNA repair biology captured by each assay: the HRD immunofluorescence assay measures ongoing DNA repair deficiency, whereas the MutSig3 signature captures the historical mutational footprint or “scar” of prior homologous recombination defects.

**Figure 2.**
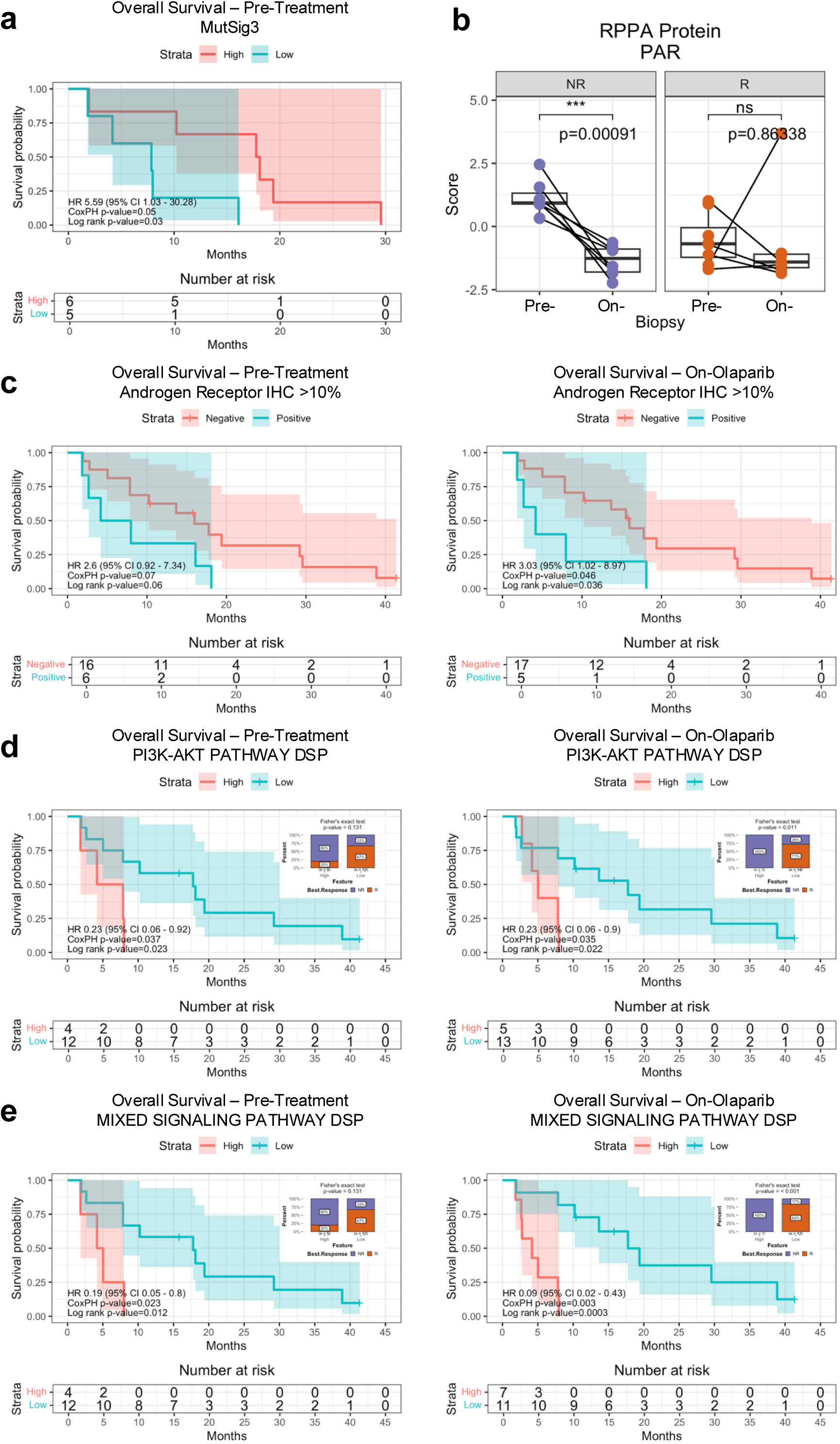
Malignant cell-intrinsic signatures of response to olaparib and durvalumab. **a)** *Mutation Signature 3 (MutSig3).* Kaplan-Meier analysis of Overall Survival (OS) by High (red) vs. Low (blue) MutSig3 status from whole-exome sequencing of pre-treatment biopsies. Hazard ratios (HRs) and p-values from Cox proportional hazards (CoxPH) and log-rank tests are shown. Shaded areas indicate 95% confidence intervals (CI) for the log survival probability. OS was measured from the start of olaparib until death. **b)** *PAR protein levels.* Boxplots comparing PAR protein levels from RPPA in Pre-Treatment (Pre-) and On-Olaparib (On-) biopsies, separated by RECIST non-responders (NR; purple) and responders (R; orange). Lines connect paired Pre- and On- samples from the same participant. P-values from paired Student’s t-test comparing Pre- versus On- biopsies within each group. **c)** *Androgen Receptor (AR) IHC.* Kaplan-Meier analysis of OS stratified by AR IHC (≥10% positive cells, blue; <10%, red) in Pre-Treatment (left) and On-Olaparib (right) biopsies. OS measured from olaparib start until death or data cutoff; right-censoring denoted by “+”. **d-e)** *Signaling pathway activity.* Kaplan-Meier analysis of OS stratified by High (z-score percentile ≥50, red) or Low (<50, blue) DSP signaling pathway protein expression. Insets: stacked bar plots of percent NR (purple) and R (orange) in High (left) and Low (right) groups; p-values from Fisher’s exact test for High vs. Low. **d)** PI3K-AKT pathway activity. Positive: p-AKT1 and p-GSK3B ≥50 percentile. **e)** Mixed signaling pathway activity. Positive: at least 3 of p-GSK3B, p-PRAS40, AKT, or p-HER2 ≥50 percentile.

RPPA proteomics data were generated from a subset of samples, including 15 pre-treatment biopsies, 15 on-treatment biopsies, and 12 matched pairs. Protein PARylation (PAR), catalyzed by PARP upon binding single-strand DNA breaks, was elevated in all pre-treatment biopsies from non-responders but in only 2 responder samples (Participant B and I; Extended Data Fig. 1c). Although PAR levels were significantly higher at baseline in non-responders (p=0.0023 NR vs. R, Extended Data Fig. 2e), they decreased on olaparib to levels similar to those in the responders (Pre- vs. On- paired t-test p=0.00091, Fig. 2b; NR vs. R on-olaparib p=ns, Extended Data Fig. 2e), indicating that lack of clinical benefit was not due to insufficient PARP inhibition.

Androgen Receptor (AR) expression has been shown to suppress immune activity across multiple tumor types and attenuate the response to ICB in prostate cancer^41–43^. In the AMTEC study, participants with nuclear AR expression >10% by clinical IHC were similarly less responsive to olaparib/durvalumab, with AR positivity predictive for shorter OS in on-olaparib biopsies (p=0.036; Fig. 2c). High AR expression (>70% positive) corresponded perfectly with Luminal Androgen Receptor subtype (LAR; see below) in non-responders as previously observed^44^, but was not predictive of response across the entire cohort, suggesting the presence of multiple, distinct mechanisms of resistance to this therapeutic combination.

PI3K-AKT and Receptor Tyrosine Kinase (RTK)-MAPK pathway activation is associated with therapeutic resistance, including to PARPi and ICB^15,45,46^. To evaluate whether activation of these pathways influenced the response to olaparib/durvalumab, we catalogued PI3K-AKT and RTK-MAPK gene alterations and pathway activation via NanoString DSP phosphoprotein profiling. PI3K-AKT gene alterations were present in biopsies from 3 of 14 responders (21%) participants and 7 of 10 non-responders (70%) (Extended Data Fig. 1c). These associations were statistically significant for response and PFS but not OS (pre-treatment p=0.013, 0.018, and ns; on-olaparib ns; Extended Data Fig. 2f). *PTEN* copy number loss alone was significantly associated with best response, PFS, and OS (on-olaparib p=0.029, 0.008, and 0.009; Extended Data Fig. 2g), indicating that some PI3K-AKT pathway mutations may be passengers with negligible impact on therapeutic outcome. RTK-MAPK pathway aberrations were present in only two participants: one with SD and one with PD as best response.

Samples with elevated phosphoproteins in the PI3K-AKT signaling pathway (p-AKT1 plus p-GSK3B) frequently harbored concurrent pathway gene alterations: 9 of the 10 pathway-activated samples had a PI3K-AKT pathway gene alteration, including all 9 non-responders (Extended Data Fig. 1c). Notably, however, 8 samples with PI3K–AKT gene aberrations lacked detectable pathway activation, demonstrating the importance of phosphoprotein profiling in determining relevant downstream effects of gene alterations. Elevated protein phosphorylation accurately predicted poor therapeutic response, particularly in the on-olaparib biopsies: p-GSK3B alone was significantly associated with best response and PFS (p= 0.008 and 0.047; Extended Data Fig. 2h), while PI3K-AKT pathway activation correlated with response, PFS, and OS (p=0.011, 0.026, 0.022; Fig. 2d, Extended Data Fig. 2i)

Although RTK-MAPK pathway activity alone was not associated with clinical outcomes, a combined PI3K-AKT/RTK-MAPK pathway protein signature (AKT, p-GSK3B, p-PRAS40, and p-HER2) was associated with PD and reduced PFS and OS (“Mixed Signaling Pathway”; Fig. 2e, Extended Data Fig. 1c, 2j). Notably, activation of this combined signaling pathway in the on-olaparib biopsy was strongly associated with poor clinical outcomes, including best response, PFS, and OS (on-olaparib p<0.001, p=0.001, and p=0.0003), suggesting these signaling proteins may be activated as an adaptive response to olaparib stress. Indeed, the mixed signaling signature demonstrated high predictive accuracy for identifying non-responders, with a positive predictive value (PPV) of 0.80 in pre-treatment biopsies and 1.00 in on-olaparib samples.

### Immune Contexture Analyses

In parallel with our analyses of gene and protein expression within malignant cells, we deeply evaluated the tumor microenvironment to identify features predictive of benefit from the olaparib/durvalumab combination. Previous work has shown that olaparib alters the immune landscape in preclinical models and patient tumors, and that immune contexture is a key determinant of durvalumab activity^47^. We therefore emphasized immune contexture in our microenvironment analyses.

As expected, analysis of the RNAseq, mIHC, and DSP data identified multiple immune features and markers associated with PFS, OS, and RECIST best response, including gene expression signatures, cell phenotypes, and protein markers, (summarized in Extended Data Fig. 1c and Extended Data Table 1d; only features that passed Boruta feature selection with Bonferroni adjustment and showed robust performance based on the harmonic mean of precision/recall (F1 score) are shown). Notably, the specific predictors varied from platform-to-platform, likely reflecting technical and assay-specific differences we present in the Discussion below.

To identify cross-platform response predictors that minimize the technical nuances of each assay platform, we first summarized the overall immune activation state within each platform by combining the most predictive individual markers and signatures (see Methods and below). Then, we integrated these single-platform summaries to generate a multi-omic immune consensus signature across platforms.

For the single platform summaries, the RNAseq summary signature was defined as high enrichment scores in at least 10 of the 17 significant immune RNA signatures (“IMMUNE RNA SUMMARY”; Extended Data Fig. 1c), the mIHC summary signature was defined as high levels of 9 or 10 mIHC cell types (“IMMUNE mIHC SUMMARY”), and the DSP consensus immune signature was defined as having high levels of at least 40% of the Immune DSP proteins (“IMMUNE DSP SUMMARY”; 40% was used due to variability in the number of successful DSP assays per sample). Each of these immune summary signatures was significantly associated with favorable PFS, OS, and RECIST best response, with stronger associations observed in the on-olaparib biopsies across the platforms (Fig. 3a-c; Extended Data Fig. 3a-c). Notably, only the immune DSP summary signature was predictive in untreated samples.

**Figure 3.**
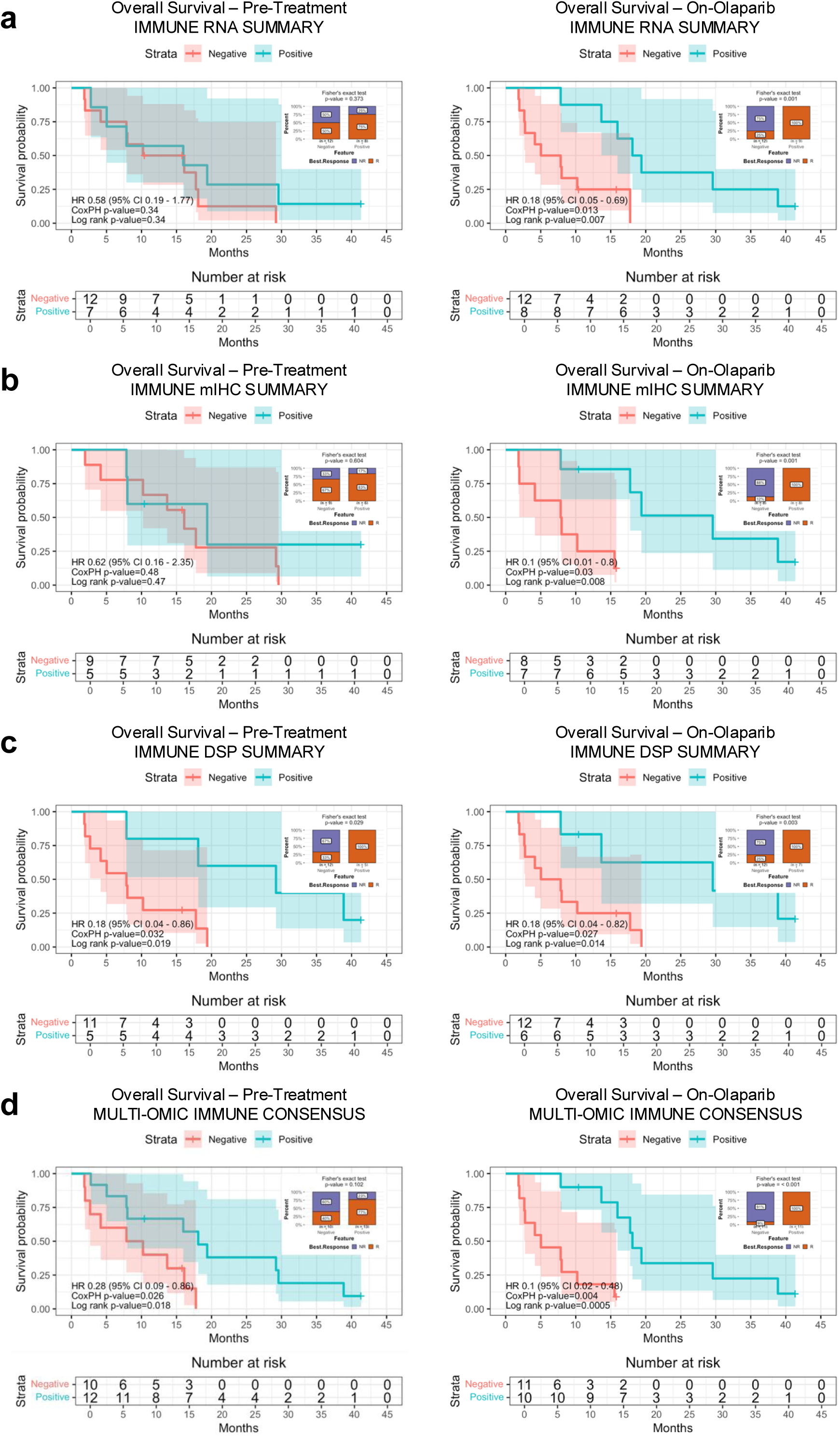
Microenvironment signatures of response to olaparib and durvalumab: Immune Consensus signatures. **a-d)** *Multi-omic immune summary signatures.* Kaplan-Meier OS in Pre-Treatment (left) and On-Olaparib (right) biopsies. Samples classified as Positive (blue) or Negative (red) based on predefined signature thresholds. HRs and p-values from CoxPH and log-rank tests are shown; shaded areas indicate 95% CI for the log survival probability. OS measured from olaparib start until death or data cutoff; right-censoring denoted by “+”. Insets: stacked bar plots of percent NR (purple) and R (orange) in Negative (left) and Positive (right) groups; p-values from Fisher’s exact test for Negative vs. Positive. Features shown achieved F1 >0.7 from random train/test split analysis and were significant (p <0.05) in pre-treatment and/or on-olaparib analyses. **a)** Immune RNA summary signature. Positive: ≥10 of 17 Immune RNA Signatures (Extended Data Figure 1c). **b)** Immune mIHC summary signature. Positive: ≥9 of 10 mIHC Cell Types (Extended Data Figure 1c). **c)** Immune DSP summary signature. Positive: ≥40% of Immune DSP proteins (Extended Data Figure 1c). **d)** Multi-Omic Immune Consensus signature. Positive: at least one positive summary signature (Extended Data Figure 1c).

The sample-level summary calls were highly concordant across platforms: 83% for RNA vs. mIHC, 79% for RNA vs. DSP, and 77% for mIHC vs. DSP (Extended Data Fig. 1c: compare “IMMUNE RNA SUMMARY”, “IMMUNE mIHC SUMMARY”, and “IMMUNE DSP SUMMARY”; see Methods for threshold selection). This demonstrates that combining predictors within each assay platform can effectively compensate for technical differences across modalities, as detailed in Discussion.

Importantly, the multi-omic immune consensus signature – defined as positive for at least one of the single-platform immune summaries – strongly predicted clinical outcomes, including best response (p<0.001), OS (p=0.0005), and PFS (p=0.002) in the on-olaparib biopsy (“MULTI-OMIC IMMUNE CONSENSUS”; Fig. 3d, Extended Data Fig. 3d). The predictive value in the on-therapy biopsy was greater than the pre-therapy biopsy (PPV: 0.91 on- vs. 0.77 pre-); however, the pre-therapy biopsy was still predictive of OS but not best response or PFS. Taken together, evaluation of immune contexture, particularly within the on-olaparib biopsy, provides a compelling ability to predict clinical response to combined olaparib/durvalumab therapy.

The pathways and cell types identified by the combined Boruta/F1 analysis and incorporated into the consensus signatures are presented in Extended Data Figure 3. Several RNA-based GSEA Hallmarks – Inflammatory Response, Allograft Rejection, Interferon Gamma Response, IL6/JAK/STAT3 Signaling, IL2/STAT5 Signaling, and Interferon Alpha Response – were variably associated with RECIST best response, PFS, and OS. Multiple adaptive immune cell populations inferred by GSVA were also variably associated with clinical outcomes. Among T cells, Activated CD8^+^ T Cells, Effector Memory T Cells, T Helper Cells, and Cytotoxic Cells, were most prominent. Regulatory T Cells, typically viewed as immunosuppressive, were unexpectedly associated with favorable RECIST best response in the on-olaparib biopsy (Extended Data Fig. 3n). B Cells were also predictive of response in this analysis (Extended Data Fig. 3u).

Among innate immune populations, Immature Dendritic Cells and NK CD56^dim^ natural killer cells were identified as potential predictors (Extended Data Fig. 3l,s), suggesting that activation of the innate immune response may contribute to benefit from olaparib/durvalumab. This is particularly noteworthy in participants H and I, who despite having a good response to therapy, had *beta-2 microglobulin (B2M)* mutations that are predicted to significantly impact B2M protein function, potentially rendering tumor cells resistant to adaptive immune cell killing. Indeed, the only significant RNA-based immune cell signature in participant H was NK CD65^dim^ natural killer cells.

Potential biomarkers on the DSP and mIHC platforms were generally consistent with those identified on the mRNA platform, including associations in the on-olaparib but not pre-therapy biopsy. These include Activated CD8^+^ T Cells, Ki-67^+^ CD8^+^ T Cells, FOXP3^-^ CD4^+^ T Cells (but not FOXP3^+^ CD4^+^ T cells) and CD20^+^ B Cells on the mIHC platform, and high CD45 and CD3 protein levels in both the tumor and stroma compartments on the DSP platform. Interestingly, CD8 and granzyme A (GZMA) were significant only in the stroma compartment by the Boruta/F1 approach. Whether this reflects differential localization of these populations or a technical challenge with the small number of samples remains to be elucidated.

PD-L1 tumor cell positivity was assessed by IHC using two different cutoffs due to discrepancies in the literature^48^. PD-L1 tumor proportion scores ≥10% were observed in at least one biopsy from 7 of the 24 participants, including 5 of 14 (36%) demonstrating clinical benefit (CR/PR/SD) and 2 of 10 (20%) participants who did not (PD; Extended Data Fig. 1c). Using a lower cutoff of ≥1%, positive PD-L1was observed in at least one biopsy from 13 of the 24 participants, including 8 of 14 (57%) demonstrating benefit (CR/PR/SD) and 5 of 10 (50%) who did not (PD). Although not statistically significant, participants with PD-L1 ≥1% were more likely to achieve CR/PR/SD, aligning with other studies indicating an association between PD-L1 positivity and response to ICB in mTNBC^49,50^.

In contrast, PD-L1 assessed by DSP was significantly predictive of best response, with similar associations in the pre- and on-olaparib biopsies (p= 0.022 and 0.035). The greater predictive value of DSP PD-L1 may reflect the platform’s higher sensitivity and linear quantitative range compared with IHC, which is reported as a dichotomous variable (positive or negative). Differences may also reflect variability in pathologist-interpreted positivity thresholds, or slight differences in sample sets.

The only immune-related predictor identified by the combined Boruta/F1 analysis from the RPPA platform was ZAP-70, likely reflecting the presence of T cells in the samples (Extended Data Fig. 3mm). The absence of other immune markers may relate to the bulk nature of the RPPA tissue specimens, which included tumor, immune, and adjacent normal cells, possibly diluting immune-specific signals.

### Olaparib-induced changes

In the analyses above, biopsies from participants with better responses and longer survival tended to have higher levels of malignant cell-intrinsic and immune-related features than those from non-responders, suggesting that outcomes may be influenced by adaptive changes in both tumor and infiltrating immune cells in response to olaparib. To investigate this further, we examined olaparib-induced changes in matched pairs of pre- and on-olaparib biopsies to identify statistically significant changes correlated with RECIST best response.

As shown in Fig. 4 and Extended Data Fig. 4, olaparib treatment changed multiple immune-related markers in matched biopsy pairs. Markers that increased in responders but decreased in non-responders include Activated CD8^+^ T Cells RNA, Immune Cells by mIHC, and stroma HLA-DR by DSP (p=0.00086, p=0.0018, and p<0.001; Fig. 4a). CD3, CD4, and HLA-DR in tumor DSP increased in responders but remained unchanged in non-responders (p=0.002, p<0.001, and p<0.001; Fig. 4b). Effector Memory T Cells RNA and Granzyme A (GZMA) in tumor DSP were unchanged in responders but decreased in non-responders (p=0.0016 and p<0.001; Fig. 4c). Finally, the RPPA protein B7-H3, which suppresses T cell activation, decreased in responders but increased in non-responders (p<0.001; Fig. 4d). Although not significant in the comparison of change magnitude between response groups, PD-L1 significantly increased on olaparib in non-responders but remained unchanged in responders by paired biopsy t-test (Extended Data Fig. 4g).

**Figure 4.**
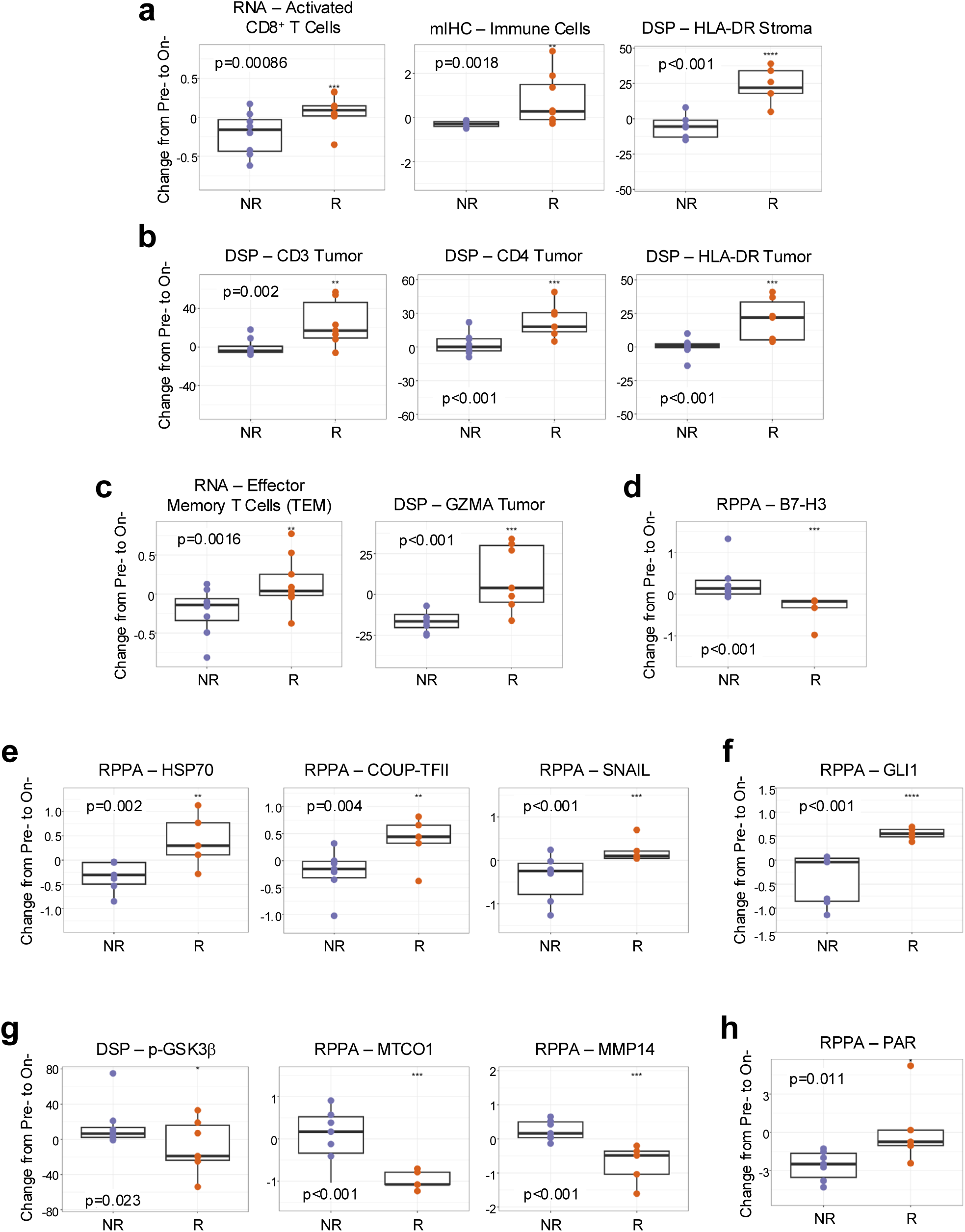
Olaparib-induced response markers and signatures. **a-h)** *Changes following olaparib treatment.* Boxplots showing changes in feature expression from Pre-Treatment (Pre-) to On-Olaparib (On-) for paired samples from the same participant. Positive values = higher expression on-olaparib; negative values = lower expression. NR (purple) and R (orange) changes were compared using Student’s t-test. Features shown were identified by Boruta analyses for each dataset: RNAseq, mIHC, DSP, and RPPA. All features significant for NR vs. R (p <0.05). **a)** Immune markers increased in responders, decreased in non-responders. **b)** Immune markers increased in responders, unchanged in non-responders. **c)** Immune markers unchanged in responders, decreased in non-responders. **d)** Immune markers decreased in responders, increased in non-responders. **e)** Malignant cell markers increased in responders, decreased in non-responders. **f)** Malignant cell markers increased in responders, unchanged in non-responders. **g)** Malignant cell markers decreased in responders, increased in non-responders. **h)** Malignant cell markers decreased in both responders and non-responders.

Protein markers associated with malignant cells also changed differentially between responders and non-responders. RPPA proteins HSP70, COUP-TFII, and SNAIL increased in responders and decreased in non-responders (p=0.002, p=0.004, and p<0.001; Fig. 4e), while GLI1 only increased in responders (p<0.001; Fig 4f). In contrast, p-GSK3β (DSP) and MTCO1 and MMP14 (RPPA) showed the opposite pattern, decreasing in responders and increasing in non-responders p=0.023, p<0.001, and p<0.001; Fig. 4g.). Similar to Fig. 2, PAR decreased in both groups but showed a significantly greater reduction in non-responders (p=0.011; Fig. 4h). Taken together, the association of olaparib-induced molecular changes with patient response underscores the importance of evaluating both pre- and on-treatment samples to better predict outcomes and guide patient selection for optimal therapy.

### Analyses using predefined biomarker algorithms

We next applied two TNBC marker classification algorithms derived from independent cohorts to the AMTEC pre- and on-olaparib samples to determine whether they could predict response. TNBC classifiers that use bulk RNAseq data to define luminal, basal, mesenchymal, and immune states have been refined based on our studies to include four subtypes: basal-like immune-activated (BLIA), basal-like immune-suppressed (BLIS), luminal androgen receptor (LAR), and mesenchymal (MES) ^51,52^. We have since implemented and validated this updated classification algorithm within the CLIA/CAP certified OHSU Knight Diagnostic Laboratories, adding an indeterminant (IND) category for samples with insufficient confidence to discriminate between BLIA and BLIS subtypes.

In the AMTEC cohort, tumors classified as BLIS or LAR in either the pre-treatment or on-olaparib biopsy had significantly poorer outcomes than those classified as BLIA or IND (non-BLIS/LAR), with decreased PFS and OS and a significant likelihood of RECIST best response of PD (on-olaparib biopsy p=0, p=0.005, and p=0.002; Fig. 5a, Extended Data Fig. 5a). Samples classified as BLIA were strongly associated with benefit: all five BLIA participants achieved a best response of CR/SD/PR, whereas all six BLIS/LAR participants had PD (Fig. 5b,c), indicating a 100% predictive value for these dichotomous predictors. IND samples were present in both PD and CR/PR/SD groups, though more frequently in responders. When the BLIS/LAR subset was compared with the combined BLIA/IND subset (non-BLIS/LAR), only three on-therapy biopsies (all IND) were misclassified as PD, corresponding to an 88% accuracy in predicting response. No MES tumors were present in the cohort.

**Figure 5.**
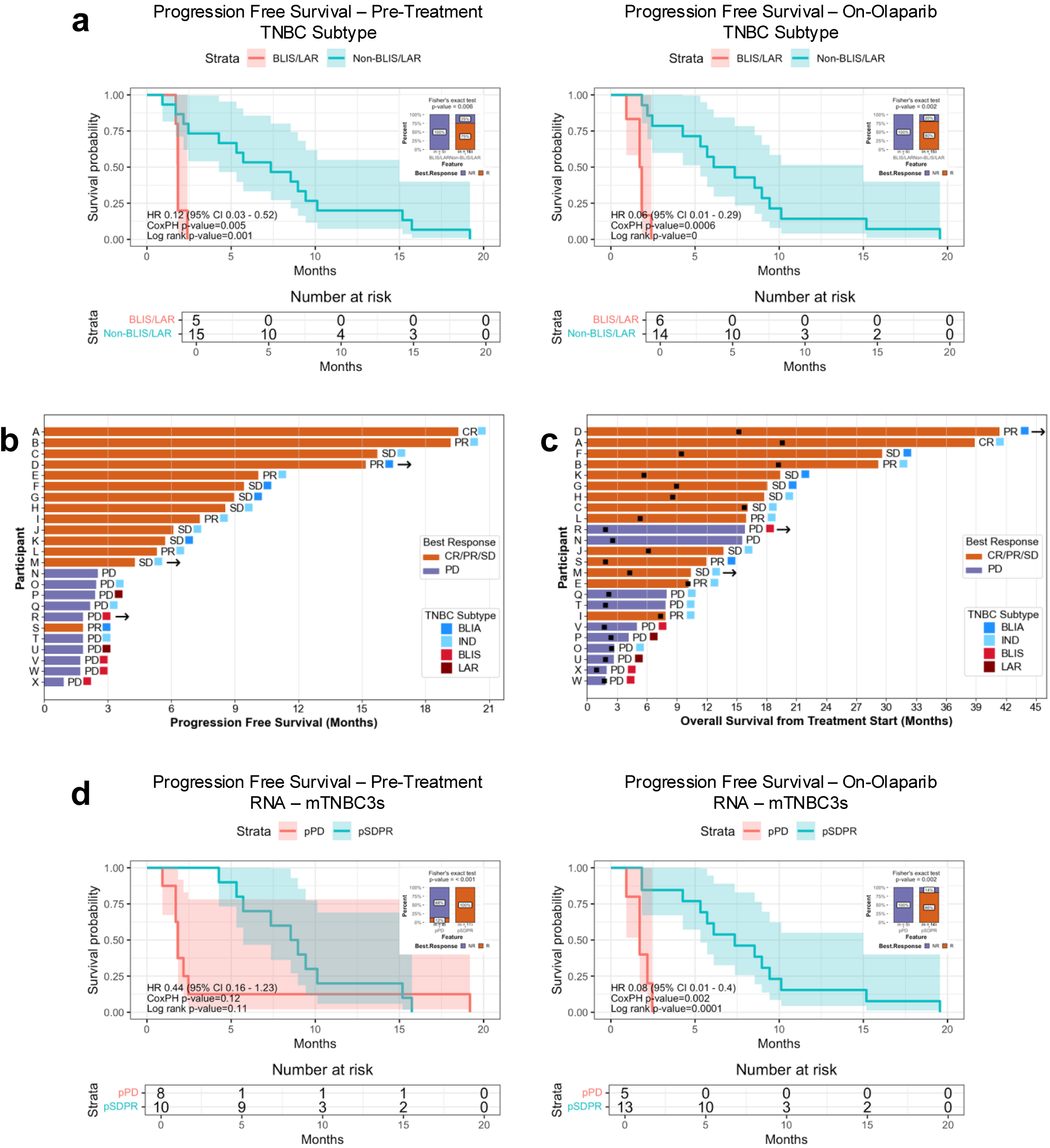
TNBC subtype correlates of response to olaparib and durvalumab. **a)** *TNBC subtypes*. Kaplan-Meier PFS by TNBC RNA subtype (BLIS/LAR, red; Non-BLIS/LAR, blue) in Pre-Treatment (left) and On-Olaparib (right) biopsies. HR and p-values from CoxPH and log-rank tests are shown; shaded areas indicate 95% CI. PFS measured from start of olaparib treatment until end of olaparib plus durvalumab. Insets: stacked bar plots of percent NR (purple) and R (orange) in BLIS/LAR (left) and Non-BLIS/LAR (right) groups; p-values from Fisher’s exact test for BLIS/LAR vs. Non-BLIS/LAR. **b)** *Progression free survival with TNBC subtypes.* Swimmer plot showing PFS (time on olaparib ± durvalumab) for 24 participants (21 efficacy + 3 pilot; Extended Data Figure 1c). TNBC subtype and RECIST best response are annotated. Subtyping was from the on-olaparib biopsy except for Participants A, C, and U (Pre-Treatment) and N (no RNA available). Arrows = alive at time of data cutoff. **c)** *Overall survival with TNBC subtypes.* Swimmer plot showing OS from treatment start until death or data cutoff for the same participants as in (b). Black squares = treatment end. **d)** *mTNBC3s gene signature.* Kaplan-Meier PFS stratified by mTNBC3s^54^predicted non-responders (pPD, red) and predicted responders (pSDPR, blue). Insets: stacked bar plots of percent NR (purple) and R (orange) in pPD (left) and pSDPR (right) groups; p-values from Fisher’s exact test for pPD vs. pSDPR.

Among the 19 participants with subtyping performed on both pre- and on-olaparib biopsies, four exhibited a change in TNBC subtype following treatment: two changed from IND to BLIS, one from BLIS to IND, and one from BLIA to IND (Extended Data Fig. 1c). All these changes involved at least one IND classification, reflecting cases where subtype assignment could not be confidently determined by the CLIA assay. Notably, these changes were clinically relevant, as the on-olaparib subtype had a higher predictive value for both survival and best response than the pre-treatment biopsy (Fig. 5a, and Extended Data Fig. 5a).

Given the strong immune activation associated with the BLIA subtype, we investigated whether its defining genes reflected an interferon-driven transcriptional program. A highly ranked subset of BLIA genes (centroid >0.5) was interferon-inducible, with a median fold change >2 based on data from interferome.org (Extended Data Fig. 5b). However, classical interferon-stimulated genes such as IRFs and IFITs were not among the top- ranked genes, suggesting that the BLIA classifier captures a novel interferon-induced gene set rather than the canonical interferon signature.

To determine which cell types express the genes defining the TNBC subtypes, we performed single cell RNAseq on a set of 13 advanced breast cancer specimens collected under an internal observational study^53^. BLIA-associated genes were predominantly expressed in immune cells, including various subtypes of T cells, NK cells, myelomonocytic subsets, and dendritic cells, consisting with their association with antigen presentation, T cell localization, and T cell killing (Extended Data Fig. 5c). Expression was minimal in other cell types, including cancer-associated fibroblasts, B cells, malignant cells, or normal epithelial cells. However, like the interferon-induced genes, classic T cell, macrophage, monocyte, or dendritic cell genes were not part of the BLIA signature.

The second TNBC marker classification algorithm evaluated was mTNBC3s, a biomarker signature we previously derived from TCGA basal breast cancers that identifies a tumor-infiltrating B cell population strongly associated with therapeutic response^54^. In our prior analyses of a subset of AMTEC samples, mTNBC3s demonstrated both predictive and prognostic ability, and resolved the IND subtype within the TNBC subtype classifier^54^. Applied here to the full AMTEC cohort, (excluding LAR samples, as mTNBC3s was not developed for this subtype), mTNBC3s correctly classified predicted responders (pSDPR) and non-responders (pPD) in 95% of pre-treatment samples (18/19) and 89% on-olaparib samples (17/19), and significantly predicted response in both biopsies (pre-treatment p<0.001 and on-olaparib p=0.002; Fig. 5d, Extended Data Fig. 1b). Among all pre-treatment markers evaluated, mTNBC3s provided the strongest predictive signal for RECIST best response; however, its performance for PFS and OS was surpassed by the on-olaparib biopsy (PFS pre-treatment p=0.11 vs. on-olaparib p=0.0001, Fig 5d; OS pre-treatment p=0.046 vs. on-olaparib p=0, Extended Data Fig. 5d), indicating that olaparib-induced immune remodeling adds additional predictive value.

### A reduced three-protein response signature with clinical translation potential

The potential biomarkers that emerged as the strongest predictors of benefit in the earlier analyses were derived from resource-intensive research platforms that use RNAseq, multi-gene expression signatures, multiplexed antibody panels, and complex computational analysis pipelines. While these approaches yield important biological insights, they are not suitable for the rapid, scalable, and regulated testing needed to guide treatment decisions in routine clinical practice because they require specialized infrastructure, technical expertise, long processing times, and most are not CLIA-compliant. To develop a more limited biomarker approach that could feasibly be implemented in standard workflows, we turned to the immune proteins quantified in our CLIA-validated DSP dataset to find markers that could be measured by routine IHC. Using a training/test framework with ANOVA-based feature selection applied to the on-olaparib samples, we identified a three-protein combination - CD3 (Tumor), CD45 (Tumor), and PD-L1 (Stroma) - as the strongest discriminator of response and used it to train a logistic regression classifier. This three-protein predictor achieved 100% accuracy in the on-olaparib cohort (Fisher’s exact test *p*<0.001; Fig. 6a; Extended Data Fig. 1c), and predicted responders (pR) exhibited significantly longer OS and PFS than predicted non-responders (pNR) (*p*=0.006 and *p*=0.0001; Fig. 6a,b). Pre-treatment samples were classified with lower, though still substantial, accuracy (84%) and showed weaker survival associations, consistent with the greater predictive value of on-therapy biopsies and olaparib-induced immune remodeling.

**Figure 6.**
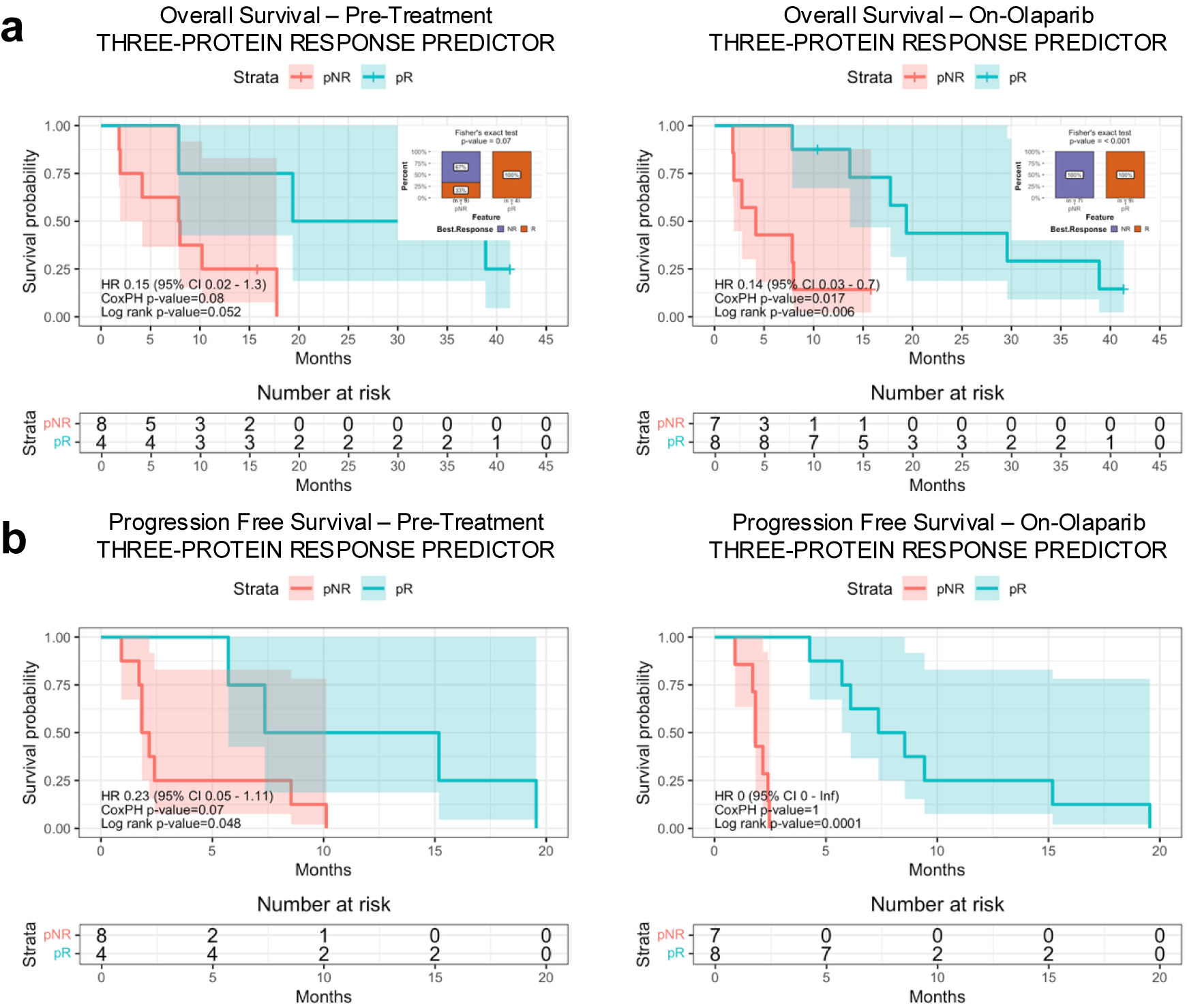
Three-Protein Response Predictor correlates of response to olaparib and durvalumab. **a-b)** *Survival and response correlates.* Kaplan-Meier OS and PFS stratified by combined weighted expression scores from three immune DSP proteins (CD45 tumor, CD3 tumor, and PD-L1 stroma): predicted non-responders (pNR score <0; red) and responders (pR ≥0; blue). HR and p-values from CoxPH and log-rank tests are shown; shaded areas indicate 95% CI. OS measured from olaparib start until death or data cutoff; right-censoring denoted by “+”. PFS measured from start of olaparib until end of olaparib plus durvalumab. Insets: stacked bar plots of percent NR (purple) and R (orange) in pNR (left) and pR (right) groups; p-values from Fisher’s exact test for pNR vs. pR. **a)** OS and best response distributions. **b)** PFS.

The three-protein biomarker set can be measured using a CLIA-compliant assay and could potentially be adapted to a simplified format suitable for broader clinical use, such as IHC. Importantly, when an on-therapy biopsy cannot be obtained, this three-protein combination could still provide actionable information for therapy selection from pre-treatment samples. Further validation in independent cohorts is needed to determine whether this three-protein signature is broadly prognostic of therapeutic response, or specifically predictive of patient benefit from olaparib–durvalumab therapy.

## DISCUSSION

The AMTEC trial demonstrated that the combination of olaparib and durvalumab provides meaningful clinical benefit in advanced mTNBC, even in patients without *BRCA1/2* mutations or other indicators of HRD. The study regimen - consisting of one month of olaparib monotherapy followed by combined olaparib and durvalumab - resulted in a median PFS of 4.8 months and OS of 15.5 months, with an overall response rate of 29% and a clinical benefit rate of 48%. Notably, participants achieving partial response or stable disease lasting ≥6 months had comparable PFS, indicating that CBR is a relevant measure of efficacy for this combination. Treatment was reasonably well-tolerated, with toxicities resolving through dose holds or reductions.

A major challenge in evaluating clinical trials with combination therapies is determining the relative “contribution of components” to overall efficacy. The AMTEC trial design, which incorporated an initial olaparib monotherapy window, enabled direct assessment of adaptive responses to PARP inhibition and their relationship to subsequent benefit from the olaparib-durvalumab combination. Because a subset of participants experienced marked and durable clinical benefit, we performed deep, integrated multi-omic analyses of pre-treatment and on-olaparib biopsies to identify potential biomarkers of response, with a focus on adaptive mechanisms induced by olaparib that might be leveraged by immune checkpoint blockade.

Using this multi-omic DNA, RNA, and protein profiling approach, we identified markers of both primary and adaptive resistance to PARPi, as well as predictors of clinical response to PARPi combined with anti-PD-L1 in pre- and on-therapy biopsies. Most markers showed limited predictive value in pre-treatment samples but were strongly associated with outcomes in on-olaparib biopsies, supporting the concept that PARPi-induced adaptive responses are key determinants of therapeutic outcome. These findings suggest that treatment strategies guided by evaluation of therapy-induced adaptive responses may optimize patient outcomes with this regimen and potentially other drug combinations. To advance clinical translation, we distilled the olaparib-induced response predictors into a set of three proteins measurable on the CLIA-compliant DSP platform, potentially enabling broad implementation of these markers in clinical practice.

Among the markers we analyzed, several were not predictive of response to olaparib/durvalumab, despite initial expectations. These include TMB; the percentage of Ki-67^+^ cells by IHC; mutations in *TP53*, *BRCA1/2* and other HRD pathway genes, or MAPK pathway genes; CD4^+^ or CD8^+^ T cell counts by IHC; and percent tumor content.

Notably, PD-L1 expression by clinical IHC also did not significantly predict outcomes. Although higher percentages of PD-L1^+^ tumor cells were more often observed in responding participants and less common in non-responders, this did not reach statistical significance. In contrast, PD-L1 protein levels quantified by DSP were significantly associated with RECIST best response in both pre- and on-olaparib biopsies. This improved performance likely reflects several advantages of DSP: (1) it is fully quantitative, reducing observer variability; (2) it captures protein expression in both tumor and infiltrating immune cells; and (3) it is sensitive to low-abundance protein, enabling detection of biologically meaningful differences that may be missed by standard IHC.

Although mutations in the HR pathway were not predictive of response, MutSig3, which reflects a genomic “scar” associated with HRD, was predictive of OS, consistent with HR status contributing to the response to olaparib plus durvalumab. Interestingly, similar findings were reported in ovarian cancer, where MutSig3, but not other HRD assays, was associated with outcomes following combined PARPi and ICB^25^. Using a functional HRD assay that counts RAD51 foci exclusively in G2-phase cells with DNA damage^39^, we found that all HRD samples in our study came from participants who benefited from the combination therapy, although HRD status did not significantly correlate with outcome, likely due to the limited number of samples definitively categorized as HRD. Importantly, this assay revealed that many tumors contained mixed populations of HRC and HRD cells, suggesting that intratumoral heterogeneity in HR competence may undermine the predictive value of HRD biomarkers and adversely affect clinical outcomes in PARPi-treated mTNBC patients.

In contrast, signaling pathway alterations showed significant associations with clinical outcomes. PI3K-AKT pathway gene alterations, particularly *PTEN* copy loss, were associated with poor outcomes in both pre-treatment and on-olaparib biopsies. Importantly, protein evidence of pathway activation through both PI3K-AKT and RTK-MAPK more strongly predicted non-response than gene alterations, particularly in the on-olaparib biopsies. These findings support the concept that signaling through these pathways is activated as an adaptive response to PARP inhibition, thereby limiting the efficacy of olaparib plus durvalumab. Consistent with this, these pathways have been implicated in responses to olaparib combination therapies in both preclinical studies and clinical trials^15,26,33,46,55^. Notably, signaling activity in our study was observed predominantly in samples lacking immune activation, suggesting that these participants may benefit from PARPi combined with an agent targeting the PI3K or MAPK pathways, rather than PARPi plus ICB.

The strongest predictors of response were related to the immune contexture, particularly immune features in the on-therapy biopsy. We and others have shown in preclinical studies that PARPi generates double-stranded DNA fragments that activate STING, leading to interferon production^22–24,28^. In the current study, although a baseline interferon signature was associated with response, a high “Interferon Gamma Response” RNA signature in the on-olaparib biopsy showed a stronger correlation with clinical benefit. Because interferon signals through the JAK-STAT pathway, the association of the “IL6/JAK/STAT3 Signaling” RNA signature with outcomes supports the concept that olaparib-induced interferon signaling contributes to therapeutic efficacy of the olaparib-durvalumab combination.

Interestingly, two participants had tumors with *B2M* mutations predicted to decrease class I HLA expression and impair antigen presentation. Loss of B2M and HLA expression has been linked to resistance to immune checkpoint blockade^56^. Despite this, both participants benefited from treatment, with one showing evidence of constitutive NK cell activation and the other exhibiting an olaparib-induced immune response consistent with the innate immune activation, in which type 1 interferons are potent inducers.

Many predictive markers were associated with immune status in the on-olaparib biopsy, supporting the contention that olaparib-mediated alterations in the immune contexture contribute to the response to the olaparib and durvalumab combination. However, not all analyzed markers showed response-dependent changes with olaparib treatment, indicating that the immune signature predictors likely reflect specific changes in immune contexture induced by olaparib rather than sample set differences or differential tumor heterogeneity between responders and non-responders. Importantly, the potential biomarkers identified in the AMTEC trial paralleled those that we identified in preclinical studies as well as in prior clinical trials^14,15,20–26^. This strongly supports the validity and mechanistic relevance of the identified biomarkers.

While immune indicators from the RNA, mIHC, and DSP platforms each predicted benefit from olaparib and durvalumab, the specific markers identified differed across platforms. This is not unexpected, as the mIHC and DSP platforms used different antibodies and were sometimes performed on different cores with varying degrees of tissue heterogeneity. Moreover, mIHC measures protein expression at the single-cell level, whereas DSP and RNA sequencing yield composite data from mixed cell populations within tumor-rich regions. Given that RNA and protein levels are often poorly correlated, especially for proteins regulated through post-translational modifications rather than gene expression^57^, discrepancies across platforms are anticipated. The limited number of samples, varying cut-off thresholds for positive versus negative calls, and lack of consensus on markers used for cell type assignments likely also influenced the statistical significance of individual markers. These factors highlight the need for further refinement of platform-specific predictors in larger cohorts. Nevertheless, by combining multiple markers into platform-specific summaries, our patient-by-patient analysis of immune activation states showed high concordance across platforms.

We used two RNAseq-based algorithms developed from external data sets to predict therapeutic response in our cohort: the BLIA/BLIS TNBC subtype classifier and the mTNBC3s outcome classifier^54^. The BLIA/BLIS algorithm is particularly notable because it has been validated and implemented in a CLIA environment, enabling its potential use for patient selection. TNBC subtype was a strong predictor of benefit on the pre-treatment biopsy with BLIA/IND vs. BLIS/LAR p-values of 0.001 and 0.04 for PFS and OS, respectively. Predictive accuracy further improved in on-olaparib biopsies, with p-values of 0 and 0.005 for PFS and OS, and PPV of pre-therapy biopsy 0.75 and on-olaparib biopsy 0.80. Thus, while not optimal, pre-treatment TNBC subtyping provides substantial predictive value for selecting patients likely to benefit from olaparib and durvalumab, especially when an on-treatment sample is unavailable.

However, clinical use of the TNBC subtyping classifier is confounded by the frequency of Indeterminant results on the CLIA platform. Among participants with Indeterminate subtype calls on the on-olaparib biopsy, most derived clinical benefit (7/10), though a subset did not (3/10). These findings underscore the need to refine this algorithm to improve its predictive value in the CLIA environment.

The previously developed mTNBC3s algorithm proved highly effective in predicting response to the olaparib and durvalumab combination in both pre-treatment and on-olaparib biopsies and, importantly, was strongly predictive in samples classified as Indeterminate by the TNBC subtype assay. This suggests that mTNBC3s may resolve a key limitation of the current CLIA-based BLIA/BLIS subtype test. Moving mTNBC3s into the CLIA environment could therefore substantially improve identification of patients most likely to benefit from PARPi and immune checkpoint inhibitor combinations. Further, because mTNBC3s predicted response in baseline biopsies in our study and has previously shown to have prognostic value^54^, this algorithm, pending additional validation, may have utility in predicting benefit beyond PARPi and immunotherapy.

Finally, we identified three proteins on the DSP platform – CD45, CD3, and PD-L1 – that, when combined, were highly predictive of response in on-olaparib biopsies. Because DSP is CLIA-compliant, this assay could be used for patient selection in future trials and warrants evaluation to determine whether it can predict response to other therapeutic regimens. Recognizing that DSP technology is not yet widely available, the small size of the analyte set also makes it feasible to implement alternative CLIA-compliant approaches, such as IHC, which could allow broader application in clinical practice.

Taken together, our analyses of pre- and on-olaparib biopsies provide insight into the relative prognostic and predictive value of the identified markers. Among the features we identified, those that were strongly associated with response in the pre-treatment biopsy, such as TNBC subtype and mTNBC3s, may reflect a mixture of prognostic and predictive information. Their performance before therapy suggests an inherent prognostic component, while their stronger associations in the on-olaparib biopsy indicate added predictive value driven by olaparib-induced biological changes. Indeed, both algorithms were developed using TNBC sample sets from patients treated with a variety of therapeutic regimens that did not include a PARPi or ICB. In contrast, our study design, incorporating both pre-treatment and on-therapy biopsies, allowed us to identify markers, such as the Three-Protein Response Predictor, that were not evident at baseline but became predictive only upon olaparib exposure. The emergence of such on-treatment markers supports the idea that some predictors specifically capture adaptive biological changes induced by PARP inhibition rather than underlying disease biology. Thus, a key strength of this study is its ability to distinguish markers with general prognostic significance from those with predictive value for the olaparib-durvalumab combination.

Despite these promising insights, our study has several important limitations. The foremost is the limited number of participants and the lack of an independent test set. While the Boruta algorithm with a Bonferroni adjustment and an F1 statistic helped mitigate false positives, prospective validation of our potential predictors in larger trials is necessary. In addition, some assays were completed on only a subset of participants due to insufficient material or tumor content, despite obtaining up to six cores per biopsy. Further, DSP data were unavailable in some samples because the assay was upgraded mid-trial, and there was insufficient material to re-run analyses. Finally, several immune characterization assays were not concordant, and as discussed above, there are likely multiple reasons for these discrepancies that require further investigation.

In summary, the AMTEC trial demonstrated marked clinical activity with acceptable toxicity. Importantly, we identified a set of potential signatures that strongly predicted participant outcomes, with a subset already validated in the CLIA environment. While some predictive biomarkers were evident in pre-therapy biopsies, the greatest predictive value was observed in on-therapy samples, consistent with adaptive responses to PARP inhibition contributing to the efficacy of the olaparib–durvalumab combination. Taken together, these results demonstrate that on-therapy biopsies capture adaptive tumor responses that are not present at baseline, underscoring their potential to improve treatment selection and provide optimal patient outcomes.

## Acknowledgements

This project was carried out with major support from the Oregon Health & Science University (OHSU) SMMART Clinical Trials Program, AstraZeneca, National Institutes of Health (NIH), National Cancer Institute (NCI) Human Tumor Atlas Network (HTAN) Research Center (U2CCA233280), Breast Cancer Research Foundation, and Prospect Creek Foundation. We thank all members of the SMMART Clinical Trials Program for support with sample collection and data management. We thank the OHSU Knight Cancer Institute BioLibrary for assistance with sample management.

## Author Contributions

GBM, ZIM, and CLC conceived and designed the study. ZIM enrolled and treated participants. ALC, JMS, ZIM, DB, TYO, and FO generated visualizations and figures. ALC, JMS, JYL, TYO, and DB performed data and statistical analyses. ALC developed software and pipelines for primary data processing. GBM, JMS, ALC, ZIM, TYO, FO, DB, and SKM interpreted the data. SS, KB, JL, DB, ML, TYO, MJR, FO, and BJ generated data. ALC and JMS managed data collection, cleaning, and curation. GBM, JMS, ZIM, and ALC wrote the manuscript. GBM, CLC, SKM, and LMC supervised the project. GBM acquired funding. All authors reviewed and revised the manuscript.

## Ethics Declaration

ZIM has received institutional grants/research support from AstraZeneca, Eli Lilly, and GSK; and received consulting fees / advisory boards from Gilead, Lilly, Merck, Novartis, AstraZeneca, and Daiichi Sankyo. LMC has received reagent support from Cell Signaling Technologies, Syndax Pharmaceuticals, Inc., ZielBio, Inc., and Hibercell, Inc.; holds sponsored research agreements with Syndax Pharmaceuticals, Hibercell, Inc., Prospect Creek Foundation, Lustgarten Foundation for Pancreatic Cancer Research, Susan G. Komen Foundation, and the National Foundation for Cancer Research; is on the Advisory Boards for Syndax Pharmaceuticals, Dispatch Pharmateuticals, Carisma Therapeutics, Inc., CytomX Therapeutics, Inc., Shasqi, Kineta, Inc., Hibercell, Inc., Cell Signaling Technologies, Inc., Alkermes, Inc., NextCure, Guardian Bio, AstraZeneca Partner of Choice Network (OHSU Site Leader), Genenta Sciences, Pio Therapeutics Pty Ltd., and Lustgarten Foundation for Pancreatic Cancer Research Therapeutics Working Group, Inc. CLC serves on the SAB for Cepheid. GBM has licensed technologies to Myriad Genetics and NanoString/Bruker; is on the SAB or is a consultant to Amphista, Astex, AstraZeneca, BlueDot, Ellipses Pharmaceuticals, ImmunoMET, Leapfrog Bio, Bruker/Nanostring, Neophore, Nerviano, Nuvectis, Pangea, PDX Pharmaceuticals, Qureator, Rybodyne, Signalchem Lifesciences, Turbine, Zentalis Pharmaceuticals; has sponsored research from AstraZeneca, Zentalis, Nanostring and Nerviano and has stock/options/financial interests in ImmunoMet and SignalChem; financial interest in Nerviano Medical and Turbine that may have a commercial interest in the results of this research and technology. This potential conflict of interest has been reviewed and managed by OHSU.

## Methods

### TRIAL OVERSIGHT

All biospecimens and data were collected under Adaptive Molecular Therapy of Evolving Cancers (AMTEC) study, ClinicalTrials.gov identifier NCT03801369. Written informed consent was obtained from all participants prior to enrollment. The Oregon Health & Science University (OHSU) Institutional Review Board approved the trial (IRB #18504). The study adhered to the ethical principles of the Declaration of Helsinki and followed the International Council on Harmonization guidelines on Good Clinical Practice, in compliance with all applicable laws, regulatory requirements, and conditions mandated by regulatory authorities and/or institutional review boards.

### STUDY DESIGN

AMTEC was a single-center, single-arm, open-label phase II observational study evaluating the efficacy of olaparib in combination with durvalumab in participants with *BRCA1/2^wt^* mTNBC (Fig. 1a). Participants with biopsy-confirmed mTNBC first underwent a pre-treatment biopsy, followed by a 4-week induction phase with olaparib (300 mg orally twice daily). A repeat on-olaparib biopsy was performed at Cycle 1, Day 28 (±7 days). After completion of the induction phase, durvalumab (1500 mg intravenously every 4 weeks) was added to continued olaparib. Treatment was planned for 12 cycles, with continuation allowed beyond 12 cycles in the absence of disease progression or unacceptable toxicity. An optional biopsy was offered at the time of progression. Participants were followed for up to one year after completing protocol-directed therapy.

### ELIGIBILITY CRITERIA

Eligible participants must have biopsy-proven mTNBC defined as ER <10%, PR <10%, and HER2 non-amplified per ASCO/CAP guidelines^58^. Up to two prior chemotherapy regimens for metastatic disease were permitted. Additional inclusion criteria included ECOG performance status ≤1, tumors amenable to serial biopsies, and no prior exposure to PARP inhibitors or immune checkpoint inhibitors in the metastatic setting.

Key exclusion criteria were active autoimmune disease requiring systemic steroids or disease-modifying therapy; history of HIV; active hepatitis B or C infection; and symptomatic central nervous system metastases or carcinomatous meningitis.

Of the 24 enrolled participants, three were excluded from the efficacy endpoint analyses: one did not receive durvalumab due to rapid disease progression, one had an orthopedic procedure unrelated to trial therapy, and one was ineligible based on hormone receptor expression on the on-treatment biopsy. This resulted in a n=21 participants for the primary endpoint ORR analysis, and n=24 for the safety analysis group.

### ENDPOINTS

The primary endpoint was overall response rate (ORR), defined as the proportion of participants achieving complete response (CR) or partial response (PR) per RECIST v1.1 criteria^32^. Secondary efficacy endpoints included clinical benefit rate (CBR; CR, PR, or stable disease [SD] lasting ≥6 months), duration of response (DOR), progression-free survival (PFS), and overall survival (OS). Safety endpoints included the incidence of grade ≥3 adverse events as defined by CTCAE v5.0.

### STATISTICAL ANALYSIS

This study used a Simon 2-stage Minimax design^59^. The null (ICB alone) and alternative (Olaparib + Durvalumab) hypotheses were: H0: π = 0.15 and Ha: π = 0.35. For the primary endpoint, a total sample size of 28 participants will achieve 80% power to detect the ORR difference of 0.20 using the 2-stage Minimax design, with one-sided type I error =0.05.

In Stage 1, 15 evaluable participants (defined as those who received at least one dose of combination therapy) were enrolled (Extended Data Fig. 1a). The study would stop for futility if ≤2 responses were observed; otherwise, it would proceed to Stage 2. The regimen would be considered promising if ≥8 responses were observed among 28 participants. All Stage 1 participants were required to complete at least one post-baseline disease assessment prior to interim analysis.

AMTEC met criteria to proceed to Stage 2, with 4 of 15 evaluable participants achieving a PR. Ultimately, 24 participants were enrolled before the trial closed to accrual in August 2024 due to recruitment challenges and shifting strategic priorities of the pharmaceutical sponsor. With no additional enrollment planned, the current report summarizes outcomes for the full trial cohort.

### SURVIVAL ANALYSIS

Survival curves for both progression-free survival (PFS) and overall survival (OS) were calculated using the Kaplan-Meier estimator and plain or log 95% confidence intervals. Cox proportional hazards models were fitted to estimate hazard ratios (HR). Reported unadjusted p-values are indicated for both the logrank score test and Cox regression model test to evaluate statistical differences between groups. When evaluating OS, data were right-censored for events that had not occurred before completion of the study and are indicated on OS curves (+). For all survival analyses, living participants were censored at the data cutoff of April 15, 2025.

### SAMPLE AVAILABILITY

Pre-treatment and on-olaparib biopsies were obtained from the 21 participants evaluable for the primary endpoints. Samples were placed in appropriate preservative (FFPE or flash frozen for RPPA) within 3 minutes to preserve post-translational modification, particularly phosphoproteins^60^. Assay success varied across samples and platforms; Extended Data Fig. 1c summarizes which assays were completed for each sample. Neither the pre-treatment nor the on-olaparib biopsy for Participant C had adequate tumor content for analysis, so Genetrails and IHC data from a biopsy obtained 90 days previously was used for analysis. The pre-treatment biopsy from Participant N contained insufficient tumor for assays other than IHC, and RNAseq failed for both samples from this participant. NanoString DSP was successful on most samples, as it requires limited tumor content. mIHC failed in a subset of samples that became detached from the matrix, a known problem with these assay platforms^61^. RPPA, which requires larger amounts of material, was performed only on a subset of participant samples.

To increase statistical power for marker and feature analyses, samples from a three-participant pilot study were also included^14^. This IRB-approved study (OHSU IRB #18239; NCT03544125) served as a precursor to the AMTEC trial and followed the identical study design, with samples collected at the same time points and participants receiving the same drugs and dosing. The participant IDs from this pilot study were B, J, and U (Extended Data Fig. 1c).

### FEATURE SELECTION

Feature selection was conducted using two approaches for discrete vs. continuous data. For continuous values, the Boruta algorithm, a method built around random forests to compare feature importance against randomly permuted shadow features^62^. Unlike alternative feature selection methods that identify a minimal-optimal feature subset, Boruta identifies all features relevant for predicting outcome, which is useful for exploratory biomarker discovery in small sample sizes. This approach provides control against false positives while maintaining sensitivity to detect true associations. For each data type, Boruta was applied with Bonferroni multiple testing correction (mcAdj=TRUE) to adjust for the number of features tested at a significance threshold of p<0.05. To reduce stochasticity, an iterative approach was implemented where Boruta was executed 50 times with different random seeds (seeds 1-50) at a more stringent threshold (p<0.01) for both pre-treatment and on-olaparib biopsy timepoints. Features identified as "Confirmed" or "Tentative" across iterations were retained, and a final Boruta analysis (seed=42, p<0.05) was performed to determine the final feature set.

For data with discrete labels, predictive performance was assessed using F1 score through repeated stratified train-test splits. For continuous data that was subsequently discretized, only features that passed the initial Boruta analysis were considered in the F1 score evaluation. Prior to analysis, optimal label mapping was determined by evaluating which assignment (e.g., Positive=Responder vs. Negative=Responder) maximized F1 score. For each feature, 20 independent splits were performed with a 70:30 train-test ratio using stratified sampling to maintain class balance. Performance metrics including F1 score, precision, recall, and accuracy were calculated for both training and test sets across all splits. A final independent test was conducted using a 60:40 split. Median F1 scores across splits were used to rank feature predictive power. Only features with an F1 score ≥70 are shown.

### STASTICAL TESTING

Following feature selection with Boruta and F1 scores, statistical testing was conducted using student’s t-test for continuous datasets or Fisher’s exact tests for discrete datasets. Comparisons of continuous variables between responder (R) and non-responder (NR) groups were performed using two-sided t-tests, where non-responders were used as the reference group. Associations between categorical variables and treatment response were evaluated using Fisher’s exact test. Unadjusted p-values were reported with adaptive precision for up to 4 decimal places.

### CLINICAL IMMUNOHISTOCHEMISTRY (IHC)

Tissue was formalin-fixed for ∼12 h based on ASCO/CAP guidelines^58^. ER, PR, AR, HER2, and Ki-67 IHC stains were performed in the OHSU Knight Diagnostic Laboratories (KDL) on formalin-fixed, paraffin-embedded tissue, using pre-diluted antibodies intended for diagnostic use in a biotin-free protocol on a Ventana Benchmark instrument. All antibodies were sourced from Roche. Staining was interpreted and scored by a pathologist. <u>ER, PR, and AR:</u> Antibodies include anti-ER (CONFIRM clone SP1), #790-4324; anti-PR (CONFIRM clone 1E2), #790-2223; and anti-AR (Cell Marque clone SP107), # 760-4605. ER, PR, and AR stains are considered negative if there is nuclear staining in < 10% of the tumor cells. <u>HER2:</u> Staining was performed with Anti-HER2/neu (PATHWAY clone 4B5), #790-2991. HER2 scoring and interpretation are per the Ventana scoring guide as expanded by the ASCO/CAP 2013 guidelines. <u>Ki-67:</u> Staining was performed with anti-Ki-67 (CONFIRM clone 30-9) and reported as percent positive cells. <u>PD-L1:</u> Anti-PD-L1 staining with clone 22C3 was performed by PhenoPath Laboratories (Seattle, WA) using pharmDx (Agilent) or by KDL using Dako #M3653. Positive cells were scored by Tumor Proportion Score. Staining with clone SP263 (Roche Ventana #790-4905) was used when clone 22C3 staining was unavailable (Participants A, D, T, V).

### NGS: GENETRAILS® COMPREHENSIVE SOLID TUMOR PANEL

Targeted DNA sequencing was conducted at the CLIA-certified and CAP-accredited OHSU Knight Diagnostic Laboratories using the GeneTrails Solid Tumor Panel assay. DNA was extracted from tumor-rich regions that were macro-dissected from FFPE tissue. Library preparation utilized custom QiaSeq chemistry (QIAGEN) with multiplexed PCR, and sequencing was performed on an Illumina NextSeq 500/550. The DNA library was created with 9,228 custom-designed primer extension assays, covering 613,343 base pairs across 125 to 225 cancer-related genes. The panel is routinely sequenced at an average read depth exceeding 2,000 to ensure highly sensitive detection of SNVs, short insertions/deletions, and copy number alterations.

PI3K-AKT pathway genes include the following genes from the GeneTrails panel: *AKT1*, *AKT2*, *PIK3CA*, *PIK3CB, PIK3R1*, or *PTEN.* RTK-MAPK pathway genes include the following genes: *HRAS*, *KRAS*, *NRAS*, *ARAF*, *BRAF*, *RAF1*, *MAP2K1*, *MAP2K2*, *MAPK1*, *EGFR*, *ERBB2*, *ERBB3*, or *MET*. HR genes include the following*: ARID1A, ATM, ATR, ATRX, BAP1, BARD1, BRCA1, BRCA2, BRIP1, CDK12, CHEK1, CHEK2, FAM175A, FANCA, FANCC, FANCD2, FANCE, FANCF, FANCG, FANCM, MRE11A, NBN, PALB2, RAD50, RAD51, RAD51B, RAD51C, RAD51D, RAD52, RAD54L*, and *RECQL.* Immune genes include the following: *B2M, CD274, HLA-A, HLA-B, HLA-C, IDO1, IDO2, IFNGR1, IFNGR2, IRF1, JAK1, JAK2*, and *PDCD1LG*.

Only genomic alterations classified as Tier 1 (Strong Clinical Significance) or Tier 2 (Potential Clinical Significance), or gene copy number gains ≥7 or losses ≤0.5, were included in this study. For samples analyzed using earlier versions of the GeneTrails panel that lacked full coverage of these genes, whole-exome sequencing (WES) was used to determine gene status.

### WHOLE EXOME SEQUENCING (WES)

Preparation of DNA was described as above at the Knight Diagnostic Laboratories. Total nucleic acid from FFPE or buffy coat was sent to Tempus Labs Inc. (Tempus Labs, Inc., Chicago, IL, USA) for exome sequencing using the Tempus xE panel. Raw sequence reads were reprocessed at OHSU following the GATK Best Practices for Somatic Variant Calling. Briefly, raw sequence FASTQ reads were aligned to the UCSC GRCh37/hg19 human genome build using BWA-MEM (0.7.12, GATK, Broad Institute) ^63,64^, followed by marking duplicate reads (Mark Duplicates, GATK) and base recalibration (BQSR, GATK). Somatic variants were called using MuTect2 (4.0.4.0, GATK, Broad Institute) between the FFPE (tumor) and the participant’s matched normal from buffy coat. A panel of normal (PON) and the gnomAD (2.0.1) ^65^ germline reference resource were used to filter out technical sequencing artifacts and common polymorphisms, respectively. All analysis tools were run using an OHSU Galaxy instance (v19.01) ^66^. On-olaparib WES was not performed for most participants, given the short duration between biopsies.

### MUTSIG3 ANALYSIS

COSMIC mutation signatures (v2) were calculated from the somatic variants (VCF) from WES using the MutationalPatterns R package with the hg19 genome reference (BSgenome.Hsapiens.UCSC.hg19) ^40,67^. The Signature 3 (MutSig3) scores were discretized into High or Low using a threshold of 40.

### HR DETECTION ASSAY

HR efficiency was assessed as in Ozmen et al.^39^, whereby RAD51 foci, which represent functional HR repair complexes, are assessed in cells that are both Geminin positive (G2 phase) and γH2AX foci positive (double strand DNA breaks). For IF staining of formalin-fixed, paraffin-embedded (FFPE) tissue samples, 5 µm sections were deparaffinized using xylene twice for 10 minutes, 100% ethanol twice for 10 minutes, 95% ethanol for 5 minutes, 70% ethanol for 5 minutes, and 50% ethanol for 5 minutes, and then left in PBS. Tissues were antigen-retrieved in a pressure cooker using Citrate (pH 6) and DAKO antigen retrieval (pH 9) buffers. Tissues were washed twice in PBS and then were further permeabilized with 0.4% Triton X-100 in PBS for 1 hour. The excess permeabilization solution was removed by washing three times in PBS, and then a hydrophobic barrier was placed around the mounted tissue. Tissues were blocked with 3% BSA in PBS prior to antibody labeling. The tissues were then incubated with primary antibodies Rad51 (1:1500; Abcam #ab133534), Geminin (1:15, #NCL-L, Leica), and γH2AX (1:400; EMD #05-636) on a rocker for 2 hours at room temperature in a humidity chamber. After three PBS washes, secondary antibodies (1:500, Invitrogen #A21428, 1:500, Biotium #20253-1, 1:1000, Invitrogen #A21131) were applied. After three final PBS washes, all tissue samples were postfixed with 4% PFA for 10 minutes at room temperature. Hoechst staining solution (1:5000) was used and mounted in Prolong Gold Antifade mounting media. Sample images were acquired using a spinning disk confocal microscope with a 40x dry objective. Raw image files from Zen (.czi) were imported into custom ImageJ macro software for image segmentation. WEKA machine learning plugin was used for both nuclei and foci segmentation. HRC nuclei were defined as having ≥5 RAD51 foci, while HRD cells were defined as RAD51 negative nuclei with ≥3 γH2AX+ foci. The relative contributions of both HRC and HRD nuclei were used to stratify participants.

### RNA TRANSCRIPTOME SEQUENCING

Preparation of RNA and transcriptome sequencing was performed at the Knight Diagnostic Laboratories. Total nucleic acid was extracted and purified from macrodissected, tumor-rich areas of FFPE sections. Libraries were prepared using a TruSeq RNA Access library preparation kit and sequenced on an Illumina NextSeq500/550. Approximately 100 million reads were generated per sample. Gene expression was quantified relative to Gencode v24 transcripts using Kallisto^68,69^.

### GENE SET VARIATION ANALYSIS (GSVA)

GSVA, a non-parametric unsupervised method to calculate gene set or pathway scores on a per-sample basis, was conducted on the RNAseq data using the GSVA package (v1.44.5) with the 1) Molecular Signatures DataBase (MSigDB) Cancer Hallmarks (v7.5.1) and 2) 16 immune cell types gene sets^70–72^. GSVA scores values were discretized into High or Low using a threshold of 0.

### MULTIPLEX IMMUNOHISTOCHEMISTRY (mIHC)

Multiple immunohistochemistry (mIHC) was performed as previously described^73^ and following (https://www.protocols.io/view/mihc-staining-ohsu-coussens-39-lab-sop-3i6gkhe). Antibody information is listed in Supplementary Table 1. Briefly, FFPE human tissues were sectioned at 5 microns and mounted on positively charged slides (Tanner Adhesive Slides, Mercedes Medical, TNR WHT45AD). Slides were baked overnight at 55°C followed by an additional baking step for 30 minutes at 58-60°C. They were then deparaffinized in xylene and graded ethanol (Xylene 2 x 5 min, 100% EtOH 2 x 2 min, 95% EtOH 2 x 2 min, 70% EtOH 2 x 2 min, 50% EtOH 1 x 2 min, diH2O 2 x 2 min). Slides were counterstained in Hematoxylin (Dako S3301), underwent heat-mediated antigen retrieval in pH 6.0 Citra solution (BioGenex, Fremont, CA, USA), blocked in Dako Dual Endogenous Enzyme Block (S2003, Dako, Santa Clara, CA, USA), and subjected to protein blocking with 5% normal goat serum and 2.5% bovine serum albumin (BSA) in tris-buffered saline-0.1% Tween (TBST). Slides were incubated with primary antibodies for 1 hour at room temperature or 16–17 hours at 4°C. Primary antibody was washed off in TBST, and either anti-rat, anti-mouse, or anti-rabbit Histofine Simple Stain MAX PO horseradish peroxidase-conjugated polymer (Nichirei Biosciences, Tokyo, Japan) was applied for 30 min at room temperature, followed by AEC chromogen (Vector Laboratories, Burlingame, California, USA).

Images were acquired using the Aperio ImageScope AT (Leica Biosystems) at ×20 magnification. Following acquisition, coverslips were gently removed in 1×TBST while agitating. Removal of AEC and HRP inactivation was accomplished by incubating the slides in 0.6% fresh H2O2 in methanol for 15 minutes. AEC removal and stripping of antibodies was accomplished by ethanol gradient incubation and heat-mediated antigen retrieval such as described above between cycles. After washing and protein blocking, samples were subjected to the next round of staining. Some samples failed due to tissue loss during staining and image acquisition. Cell phenotypes were assigned with hierarchically gating (see Supplementary Table 2 for gating strategy) and quantified to cell densities (cells/mm^2^). Cell phenotype densities were log2 transformed followed by scaling and centering (i.e. standard scaled or z-scored) as continuous variables and discretized as “High” for samples ≥0 and “Low” for <0.

### NANOSTRING GEOMX DIGITAL SPATIAL PROFILING (DSP)

DSP was performed by the OHSU Knight Diagnostic Laboratories. A single FFPE slide per sample was used for DSP protein panel analysis. FFPE sections (4-5 micron) were deparaffinized and subjected to antigen retrieval in Citrate buffer, pH 6.0 for 15 minutes using a pressure cooker. The slides were then incubated with a cocktail of oligonucleotide-tagged antibodies together with two fluorescent-tagged morphological markers, pan-CK (Alexa 532nm) and CD45 (Alexa 594nm) at 4°C overnight, followed by blocking for 1 hour with Buffer W. After antibody incubation, slides were washed with TBS-T and post-fixed with 10% neutral buffered formalin for 30 minutes. Following a brief wash with TBS-T, the sections were incubated with SYTO-13 (FITC 525nm) for 30 minutes and then placed on the DSP instrument for sample collection. Slides were scanned and tumor regions were identified based on nuclei staining and visualization markers.

A minimum of three regions of interest (ROIs; 660 micron diameter) were selected for each sample under the guidance of a board-certified pathologist in accordance with recommended practices^74^. ROIs were chosen with the goal of maximizing tumor cell content, avoiding any areas of necrosis or sectioning artifacts on the slide. UV-released oligonucleotide barcodes were collected automatically in a 96-well plate, then hybridized with GeoMx hyb codesets at 67°C for 16 hours and quantitated using the MAX nCounter system.

Assays were run on two versions of the assay platform: 1.2 and 2.0. Some antibodies differed between platform versions (see Supplementary Table 3 for DSP antibody list), but the same analysis pipeline was used for all. A control tissue microarray was included in every run to correct for batch effects. Based on comparison with normalized signals from a 19-sample mixed breast cancer cohort, sample values were discretized with z-score percentiles. To determine appropriate thresholds for DSP protein markers, we performed a density-based bimodal distribution analysis. Density curves were computed for each protein marker using a Gaussian kernel density function where the majority of DSP protein markers followed a bimodal distribution. Using a rolling window approach, the local maximums and minimums were calculated for each marker where peaks were defined as local maximums exceeding both the leading and trailing edges of the window and valleys defined as the minimum density value between the first two identified peaks. The median valley across all markers was defined as the global threshold for either signaling associated or immune markers as density distributions for tumor signaling associated markers tended to be shifted higher. The final thresholds were set to 60 for signaling markers and 50 for immune markers.

### IMMUNE CONSENSUS SIGNATURES

For the RNAseq-based Immune Consensus (“IMMUNE RNA SUMMARY”), a sample was classified as “Positive” if at least 10 of 17 High Immune RNA Signatures (Extended Data Fig. 1c) were present. These 17 RNA signatures were identified as predictive of clinical outcomes (p <0.05), selected using the Boruta feature-selection algorithm, and demonstrated an F1 score ≥70. The signatures included: Activated CD8^+^ T Cells, Allograft Rejection, B Cells, Complement, Cytotoxic Cells, Gamma Delta T Cells, Immature Dendritic Cells, IL2/STAT5 Signaling, IL6/JAK/STAT3 Signaling, Inflammatory Response, Interferon Alpha Response, Interferon Gamma Response, NK CD56^dim^ Cells, Regulatory T Cells, T Helper Cells, Central Memory T Cells (TCM), and Effector Memory T Cells (TEM).

For the mIHC-based Immune Consensus (“IMMUNE mIHC SUMMARY”), a sample was classified as “Positive” if at least 9 of 10 High Immune RNA Signatures (Extended Data Fig. 1c) were present. These 10 mIHC Cell Types were identified as predictive of clinical outcomes (p <0.05), selected using the Boruta feature-selection algorithm, and demonstrated an F1 score ≥70. The signatures included: CD20⁺ B Cells, CD4⁺ T Cells, CD8⁺ T Cells, Immune Cells, Ki-67⁺ CD8⁺ T Cells, Ki-67⁺ Immune Cells, Ki-67⁺ TBET/EOMES/PD-1⁻ CD8⁺ T Cells, Other Immune Cells, TBET⁻ FOXP3⁻ CD4⁺ T Cells, and TBET/EOMES/PD-1⁻ CD8⁺ T Cells.

For the DSP-based Immune Consensus (“IMMUNE DSP SUMMARY”), a sample was classified as “Positive” if at least 40% High Immune DSP Proteins (Extended Data Fig. 1c) were present. For samples in which the stroma compartment was not assayed, only tumor compartment proteins were included. Eleven Immune DSP Proteins were identified as predictive of clinical outcomes (p <0.05), selected using the Boruta feature-selection algorithm, and demonstrated an F1 score ≥70. The proteins included: CD3 (Stroma), CD3 (Tumor), CD4 (Tumor), CD45 (Stroma), CD45 (Tumor), CD8 (Stroma), GZMA (Stroma), GZMA (Tumor), HLA-DR (Stroma), HLA-DR (Tumor), and PD-L1 (Tumor).

The MULTI-OMIC IMMUNE CONSENSUS was defined by integrating RNAseq, mIHC, and DSP immune consensus results. A sample was classified as “Positive” if it was positive in at least one of the individual measures: IMMUNE RNA SUMMARY, IMMUNE mIHC SUMMARY, or IMMUNE DSP SUMMARY.

### REVERSE PHASE PROTEIN ARRAY (RPPA)

Protein extracts from tumor samples were analyzed with a panel of 480 proteins and phosphoproteins as previously described^75,76^. To scale the protein expression values, the RPPA data from each participant sample was merged within the TCGA primary breast cancer and SMMART-program metastatic breast cancer RPPA datasets, using the replicate-based normalization method^57,77^. The protein expression values were then *z*-scored by the Median Absolute Deviation (MAD) from a Basal classified subset of TCGA samples.

### TNBC SUBTYPE CALLING

CLIA TNBC subtypes were computed using a refined list of 77 genes using a novel algorithm^51,52^. Sample gene expression was batch corrected using Removal of Unwanted Variation (RUVg) ^78^. Spearman’s correlation was computed between the gene expression values and each centroid. Patient samples were classified as immune activated (BLIA), basal immune suppressed (BLIS), luminal androgen receptor (LAR), or mesenchymal (MES) subtypes based correlation to the corresponding centroids. In cases where the difference between the top two centroids was less than 0.1, samples were considered indeterminate (IND).

### SINGLE-CELL RNA SEQUENCING (scRNAseq)

Thirteen biospecimens were collected from participants with metastatic breast cancer under an IRB-approved MMTERT observational study^53^ (OHSU IRB #16113). Fresh core needle biopsies were disaggregated using gentleMACS kits (Miltenyi Biotec, #130-095-929) following the manufacturer’s instructions and filtered through a 70-µm cell strainer. Cells were collected by centrifuging (300g for 7 min at 4°C) and resuspended at 700–1200 cells/µl. Live cells were isolated with a EasySep Dead Cell Removal (Annexin V) Kit (STEMCELL Technologies, #17899). Single cell suspensions of viable cells were processed using a Chromium Next GEM Single Cell 3’ Kit v3.1 (10xGenomics) following the manufacturer’s protocol. The entire mixed cell population was analyzed without sorting or enriching specific cell subtypes. Briefly, viable single cells were encapsulated into GEMs containing barcoded oligonucleotides. 5,000-10,000 GEMs were then subjected to cell lysis, reverse transcription, and cDNA amplification. Libraries were constructed from the amplified cDNA, and sequencing was performed on the Illumina NovaSeq platform. Raw sequencing data were aligned to GRCh38 genome reference using 10X software CellRanger (Version 6.1.2) with default parameters.

To ensure high-quality data, we used Scanpy^79^ to implement three quality control measures on the raw gene-cell-barcode matrix for each cell: 1) the proportion of mitochondrial genes (≤20%), 2) unique molecular identifiers (UMIs, ranging from 400 to 120,000), and 3) gene count (ranging from 200 to 10,000). Doublets were identified and removed from each sample using Scrublet^80^. Normalization of total counts per cell was performed using the normalize_total function in Scanpy, followed by log-normalization with the log1p function. Clustering was conducted using the Leiden algorithm at a resolution of 1 in Scanpy.

Marker genes for each cluster were identified using a t-test implemented in Scanpy. These marker genes were then used to annotate each cluster using publicly available databases including CellMarker^81^ and PanglaoDB^82^ to determine cell identities. These annotations were later refined using the CellTypist^83^ package with the celltypist.annotate function. The model ’Immune_All_High.pkl’ was specified, with majority voting enabled. Through this process, seven major cell type clusters were annotated: epithelial cells, natural killer cells, myeloid cells, T cells, B cells, fibroblasts, and endothelial cells. To distinguish malignant cells, we calculated and identified large-scale chromosomal copy number variation (CNV) using inferCNV^84^ and CaSpER^85^ tools for each sample based on transcriptomes. T cells and myeloid cells were used as reference cells, and epithelial cells with differing CNV patterns and higher CNV score were annotated as malignant cells.

Datasets were merged across different samples using SCVI^86^ based integration. The top 4,000 highly variable genes were used to train the VAE models, with each biopsy as covariate key. After training the initial VAE model, the annotated cell types were used to build an extended model with scANVI^87^ for better integration. After integration, NK cells, myeloid cells, T cells, B cells, and fibroblasts were further classified using the following publicly available scRNAseq datasets: NK cells (GSE212890^88^); T cells, myeloid cells, and B cells (GSE169426^89^) and fibroblasts (GSE103322, GSE132465, GSE154778, and GSE212966^90^). The label transfer method was performed using the scArches^91^ algorithm, following the best practices described previously^93^. Dot plots were generated using the Scanpy DotPlot function to visualize the expression patterns of a panel of previously defined TNBC subtype markers^52^ across various cell types.

### mTNBC3s OUTCOME CLASSIFIER

A 34 gene classifier, mTNBC3s, designed to improve upon the CLIA TNBC BLIA/BLIS subtype classifier by reducing the number of indeterminate calls was previously developed and applied to a subset of the AMTEC cohort (see Bottomly et al. ^54^ for detailed methods). The same classifier was applied to the additional AMTEC samples in this paper. Because this model was trained on datasets that did not include LAR tumors, LAR-classified samples were excluded (Participants P and U).

### THREE-PROTEIN RESPONSE PREDICTOR

Using the normalized GeoMx DSP (z-score percentiles), the on-olaparib samples were randomly split into a 65/35 train-test set for feature selection and model development. Using the training set, combinatorial feature selection with ANOVA was used to identify the set of protein markers (continuous measurements) with the highest discriminatory power for predicting best response (categorical outcome). Combinations of 2, 3, and 4 proteins were systematically evaluated and ranked using ANOVA F-statistics (f_classif) with the top-performing feature combinations retained for model building.

A logistic regression classifier with L2 (Ridge) regularization (C=1.0, lbfgs solver) was trained using the same training set as above and the top-performing protein marker combination, including CD3 (Tumor), CD45 (Tumor), and PD-L1 (Stroma). Since feature selection was previously performed with ANOVA, L2 regularization was selected over L1 (LASSO) to retain protein features with weighted coefficients. The classifier was implemented with a fixed random seed (seed=42) and evaluated on the independent test set with performance assessed by precision, recall, F1 score, and accuracy.

### VISUALIZATIONS

Boxplots, bar plots, and Kaplan-Meier curves were plotted with R using ggplot2, ggpubr, and ggstatsplot. Swimmer plots were generated in Python using matplotlib. The CoMut plot was generated using the Python library CoMut^92^.

### DATA AVAILABILITY

All data are available through the HTAN Data Portal as part of the HTAN OHSU Atlas (https://data.humantumoratlas.org/). Raw sequencing data have been deposited to dbGAP (Project phs002371.v1.p1). Images can be viewed at the Imaging Data Commons (IDC) https://portal.imaging.datacommons.cancer.gov/.

### CODE AVAILABILITY

No new tools were developed for the analyses in this study.

## EXTENDED DATA

**Extended Data Figure 1.**
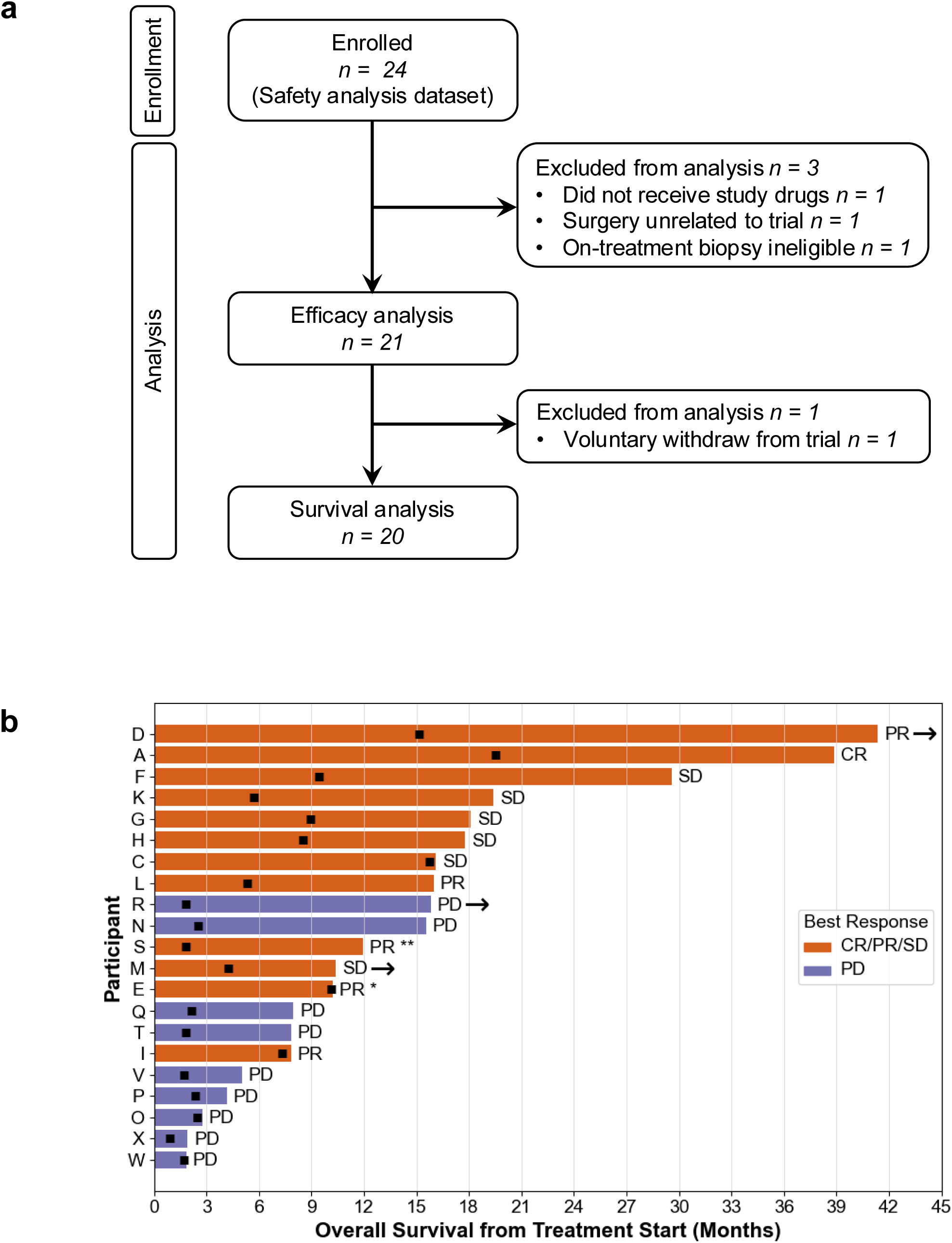
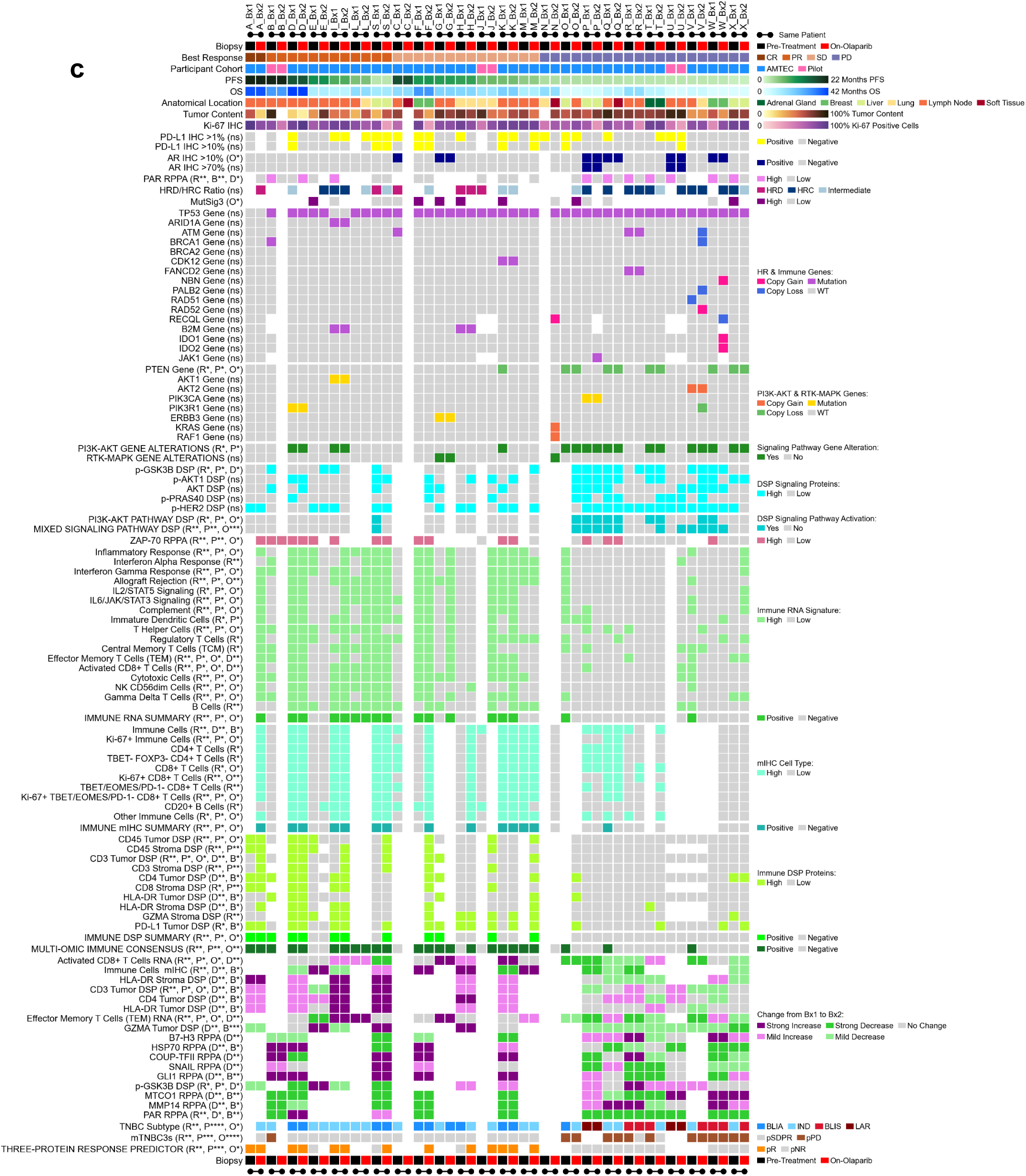
Overview of the AMTEC trial and clinical outcomes. **a)** *Trial design and enrollment.* CONSORT flow diagram summarizing participant enrollment, exclusions, and analyses in the AMTEC trial. **b)** *Overall Survival (OS).* Swimmer plot for 21 efficacy-evaluable participants. Best response by RECIST v1.1^33^, is indicated adjacent to each treatment duration bar. OS measured from olaparib start until death or data cutoff. Orange bars: participants with Complete Response (CR), Partial Response (PR), or Stable Disease (SD); purple bars: Progressive Disease (PD). * = passed away from a trial-unrelated accident; ** = voluntarily withdrew from the study. Black squares = treatment end. Arrows = alive at time of data cutoff. **c)** *Multi-omic clinical features and signatures from serial biopsies.* Summary analysis results for all clinical and molecular features in each sample. Columns are sample identifiers (participant identifier appended to biopsy number; Bx1 = pre-treatment biopsy; Bx2 = on-olaparib biopsy). Rows correspond to clinical or molecular feature and signature data. White boxes denote missing data. Features with parenthetical notations were found to be significant in one or more analyses: R = RECIST best response t-test; P = Kaplan-Meier analysis of Progression Free Survival (PFS); O = Kaplan-Meier analysis of Overall Survival (OS); B = paired biopsy t-test for responders and non-responders; D = pre-treatment to on-olaparib change t-test for responders vs non-responders. * p <0.05; ** p <0.005; *** p <0.0005; ns = not significant. See also Extended Data Tables 1c,d for feature significance summaries. **Best Response:** determined by RECIST v1.1. CR = Complete Response; PR = Partial Response; SD = Stable Disease; PD = Progressive Disease. **Participant Cohort:** AMTEC trial or Pilot study enrollment. Includes all efficacy-evaluable AMTEC participants (see Extended Data Fig. 1a). **PFS**: measured from start of olaparib until end of olaparib plus durvalumab (see Fig. 1d). **OS:** measured from olaparib start until death or data cutoff (see Fig 1e). **Anatomical Location:** organ site of the metastatic lesion from which the biopsy was obtained. **Tumor Content**: pathologist-estimated percentage of tumor cells after microdissection of tumor-rich regions from FFPE slides. **Ki-67**, **PD-L1**, and **AR IHC:** assayed clinically and defined as defined as follows: Ki-67 = % positive tumor cells; PD-L1 = tumor proportion score (TPS) ≥1% or ≥10%; AR = nuclear staining tumor cells ≥10% or ≥70% (see Fig. 2c). **PAR RPPA**: PAR protein levels measured by RPPA. High = median absolute deviation >0 relative to a Basal-classified subset of TCGA samples (see Fig. 2b, Extended Data Fig. 2e, Fig. 4h). **HRC/HRC Ratio**: the ratio of HRC to HRD nuclei. Ratios <15% = HRD, >65% = HRC, 15–65% = Intermediate (see Extended Data Fig. 2a–c). **MutSig3:** COSMIC Mutational Signature 3, discretized into High or Low using a threshold of 40 (see Fig. 2a). **HR & Immune Genes** and **PI3K-AKT & RTK-MAPK Genes:** variants reported clinically by GeneTrails. Copy Gain is ≥7 copies and Copy Loss is ≤0.5 copies. Mutations include missense mutations, nonsense mutations, indels, and splice site substitutions. See *Methods* for pathway gene inclusions. **Signaling Pathway Gene Alteration:** the presence of least one DNA alteration among the PI3K-AKT or the RTK-MAPK pathway genes (see Extended Data Fig. 2f). Note that participants K and W had only one of two biopsies with clinically reported *PTEN* loss, possibly due to tumor content variability constraining copy number loss detection. **DSP Signaling Proteins:** phosphoprotein levels within tumor ROIs measured by NanoString DSP. High = z-score percentile ≥50 relative to a mixed breast cancer cohort (see Extended Data Fig. 2h, Fig. 4g). **DSP Signaling Pathway Activation:** PI3K-AKT PATHWAY DSP activation = both p-AKT1 and p-GSK3B high. MIXED SIGNALING PATHWAY DSP activation is ≥3 high among p-GSK3B, p-PRAS40, AKT, or p-HER2 (see Fig. 2d,e and Extended Data Fig. 2i,j). **ZAP-70 RPPA:** ZAP-70 protein levels measured by RPPA. High = median absolute deviation >0 relative to a Basal-classified subset of TCGA samples (see Extended Data Fig 3mm). **Immune RNA Signature:** from GSVA scores of MSigDB Cancer Hallmarks and Inferred Immune Cell Type gene sets. Discretized into High or Low using a threshold of zero (see Extended Data Fig. 3e-u, Fig. 4a,c). **IMMUNE RNA SUMMARY:** the number of High and Low Immune RNA Signatures per sample. Positive is ≥10 High signatures (see Fig. 3a and Extended Data Fig. 3a). **mIHC Cell Type**: immune cell phenotypes assigned by hierarchical gating of protein co-expression and quantified to cell densities. High is logged/scaled z-score ≥0 (see Extended Data Fig. 3v-ee, Fig. 4a, Extended Data Fig. 4a). **IMMUNE mIHC SUMMARY:** the number of High and Low mIHC Cell Types per sample. Positive is ≥9 High cell types (see Fig. 3b and Extended Data Fig. 3b). **Immune DSP:** levels of immune proteins measured within tumor and stroma ROIs by NanoString DSP. High is z-scores percentile ≥60 (see Extended Data Fig. 3ff-ll, Fig. 4a-c, Extended Data Fig. 4a-c,g). **IMMUNE DSP SUMMARY:** the number of High Immune DSP proteins per sample. Positive is ≥40% High proteins. Stroma compartment was unmeasurable for some samples (see Fig. 3c and Extended Data Fig. 3c). **MULTI-OMIC IMMUNE CONSENSUS:** the overall immune status in a combined three-platform signature. Positive is at least one High measurement was observed for IMMUNE RNA SUMMARY, IMMUNE mIHC SUMMARY, or IMMUNE DSP SUMMARY (see Fig. 3d and Extended Data Fig. 3d). **Change from Bx1 to Bx2:** the difference in feature score between the pre-treatment and on-olaparib biopsies (see Fig. 4). For RNA, mIHC, and RPPA features, Strong Increase ≥ +0.25; Mild Increase > +0.05 and < +0.25; Strong Decrease ≤ -0.25; Mild Decrease < -0.05 and > -0.25; No Change > -0.05 and < +0.05. For DSP proteins, Strong Increase is z-score percentile change ≥ +25; Mild Increase > +5 and < +25; Strong Decrease ≤ -25; Mild Decrease < -5 and > -25; No Change > -5 and < +5. **TNBC Subtype**: Basal-Like Immune Activated (BLIA), Basal-Like Immune Suppressed (BLIS), Luminal Androgen Receptor (LAR), or Indeterminate (IND), assigned from RNAseq using a revised 77-gene classifier (see Fig. 5a-c, Extended Data Fig. 5a). **mTNBC3s:** predicted responder (pSDPR) or non-responder (pPD), assigned from RNAseq analysis of a 34-gene set predictive of response to therapy in a preliminary study of 13 AMTEC trial participants^54^ (see Fig. 5d, Extended Data Fig. 5d). **THREE-PROTEIN RESPONSE PREDICTOR:** predicted responder (pR) or predicted non-responder (pNR), assigned from NanoString DSP combined protein expression of CD45 (tumor ROI), CD3 (tumor ROI), and PD-L1 (stroma ROI; see Fig. 6).

**Extended Data Figure 2.**
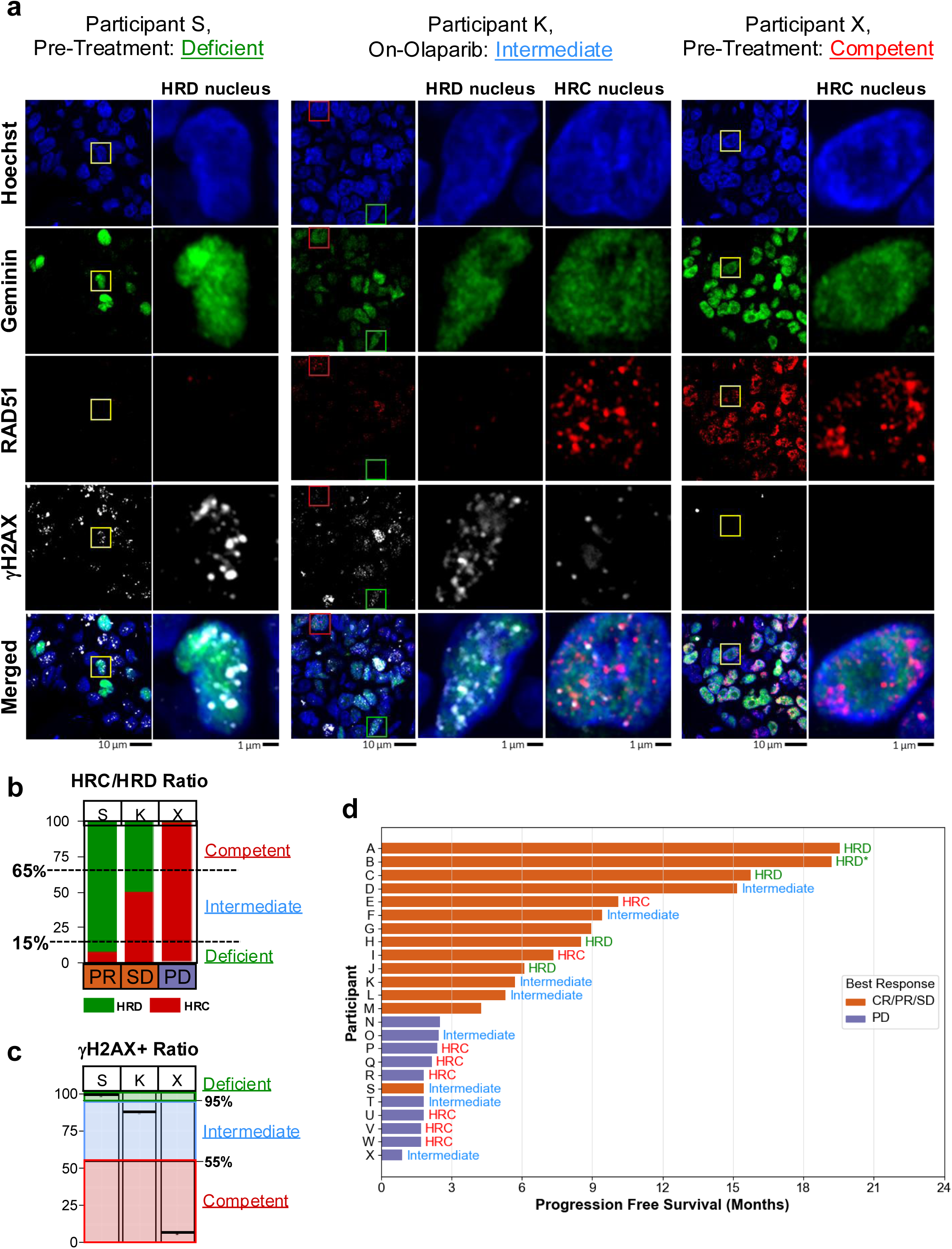
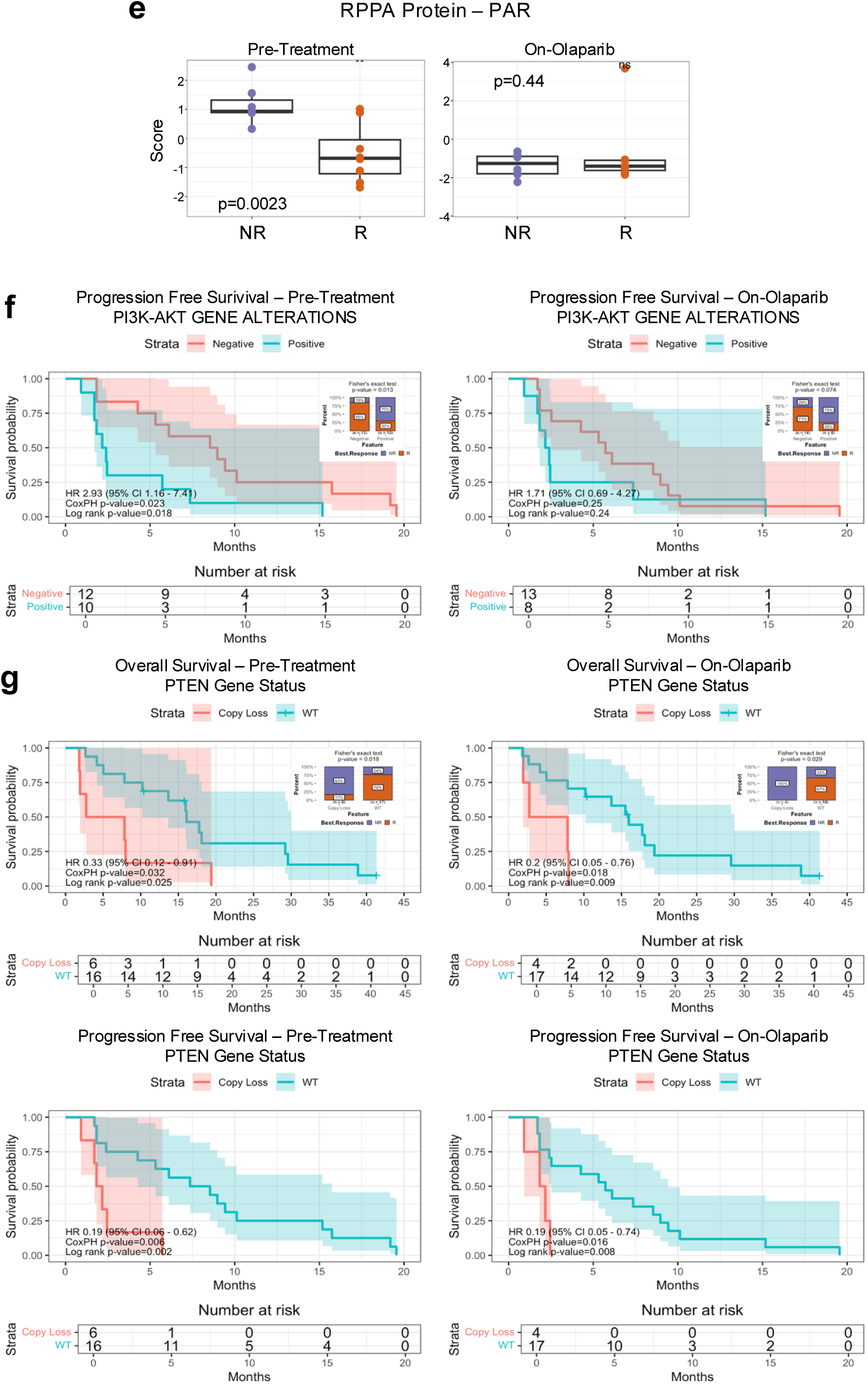
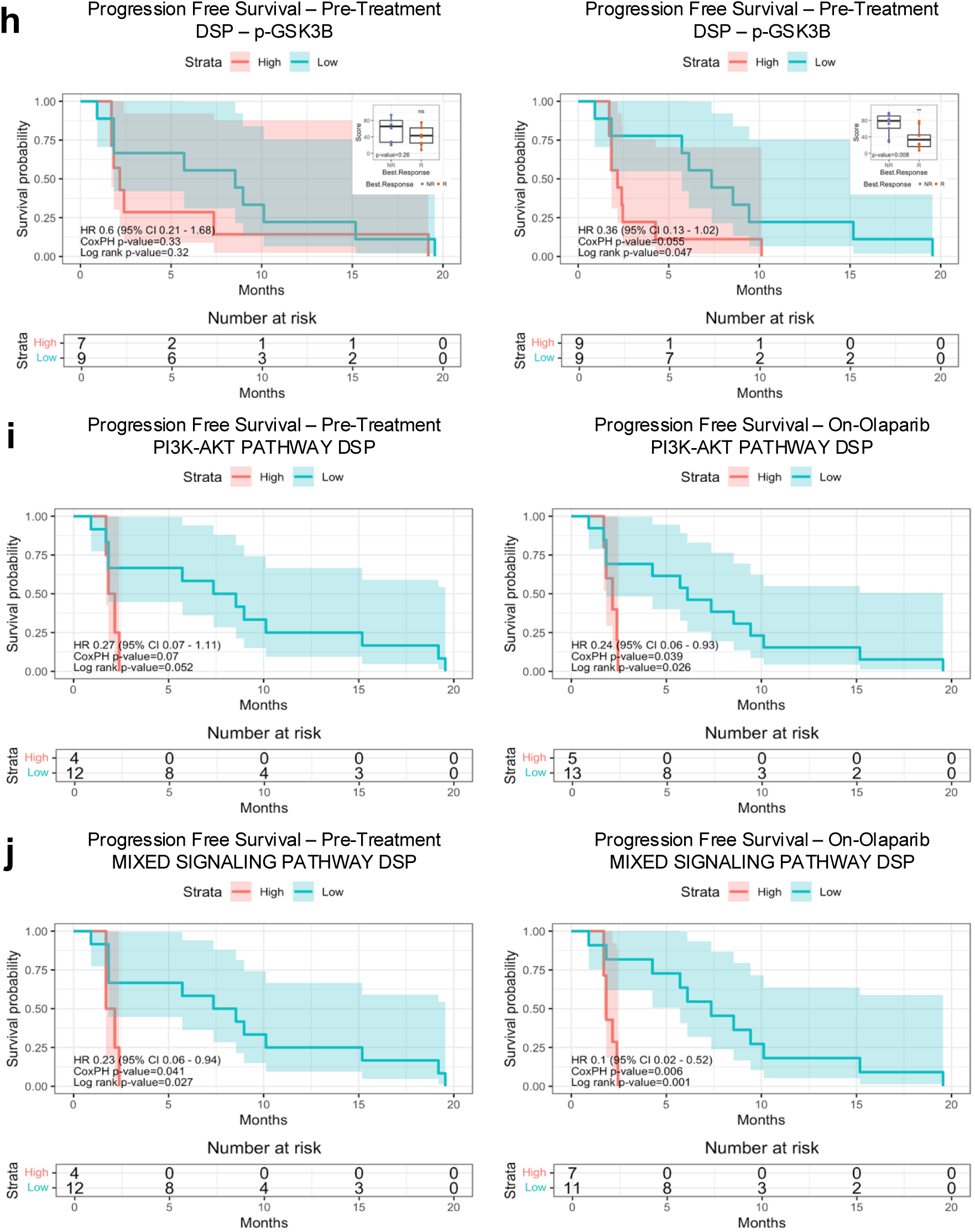
Malignant cell-intrinsic signatures of response to olaparib and durvalumab. **a-d)** *Homologous recombination (HR) repair proficiency.* Quantification of DNA repair complexes and HR status in cycling cells by immunocytochemistry. **a)** RAD51, γH2AX, and Geminin nuclear co-staining. Representative images from three samples: HR deficient (HRD; Participant S), HR competent (HRC; Participant X), and intermediate (Participant K). RAD51 and γH2AX foci were counted in Geminin-positive nuclei. HRD: <5 RAD51 foci and >3 γH2AX foci; HRC >5 RAD51 foci. Scale bar is shown as indicated. **b)** Machine learning-based HR classification. Custom machine learning segmentation results from the samples shown in (d). Sample HR classifications were defined using the ratio of HRC to HRD. Deficient is <15%; Competent is >65%, and Intermediate is 15-65%. RECIST best response is shown in orange for responders (PR, SD) and purple for the non-responder (PD). **c)** γH2AX positivity ratios. γH2AX positivity ratios calculated in the samples in (d) are represented as horizontal black bars. Samples falling within the green area are HR Deficient (>95% γH2AX^+^/Geminin^+^ co-positive nuclei) and in the red area are HR Competent (<55% co-positive nuclei). Samples falling within the blue area were Intermediate, tending toward higher γH2AX positivity and a mixture of HRC and HRD nuclei. **d)** *Progression-free survival with HRD status.* Swimmer plot showing PFS (time on olaparib ± durvalumab) for 24 participants (21 efficacy + 3 pilot; Extended Data Figure 1c). HRD/HRC ratio score is annotated, except for participant B who was germline BRCAm. Scoring was from the on-olaparib biopsy except for Participants C, D, J, P, and Q, where Pre-Treatment was used (no on-olaparib sample available). **e)** *PAR protein.* Boxplots of PAR protein levels measured from RPPA in Pre-Treatment (left) and On-Olaparib (right) biopsies from RECIST non-responders (NR, purple) and responders (R, orange). P-values from Student’s t-test compare NR and R protein levels at each timepoint. **f-g)** *Gene alterations.* Kaplan-Meier analyses of Overall Survival (OS) and Progression-Free Survival (PFS) stratified by presence of gene alterations in Pre-Treatment (left) and On-Olaparib (right) biopsies. Hazard Ratios (HR) and p-values from Cox proportional hazards (CoxPH) and log-rank tests are shown; shaded areas indicate 95% confidence intervals (CI) for the log survival probability. OS measured from olaparib start until death or data cutoff; right-censoring denoted by “+”; PFS measured from start of olaparib until end of olaparib plus durvalumab. Insets: stacked bar plots of percent NR (purple) and R (orange) in groups with or without gene alterations; p-values from Fisher’s exact test comparing each group. **f)** PI3K-AKT gene alterations. Positive is ≥1 PI3K-AKT pathway gene alteration (copy gain ≥7, copy loss ≤0.5, missense mutations, nonsense mutations, indels, or splice site substitutions). **g)** PTEN gene status. Copy loss: *PTEN* DNA copy number ≤0.5. **h-j)** *Signaling proteins.* Kaplan-Meier analysis of PFS stratified by High (z-score percentile ≥50, red) or Low (<50, blue) DSP signaling pathway protein expression. **h)** p-GSK3B protein levels. Insets: boxplots of protein levels from NR (purple) and R (orange); P-values from Student’s t-test for NR vs. R. **i)** PI3K-AKT pathway activity. Positive: p-AKT1 and p-GSK3B ≥50 percentile. **j)** Mixed signaling pathway activity. Positive: at least 3 of p-GSK3B, p-PRAS40, AKT, or p-HER2 ≥50 percentile.

**Extended Data Figure 3.**
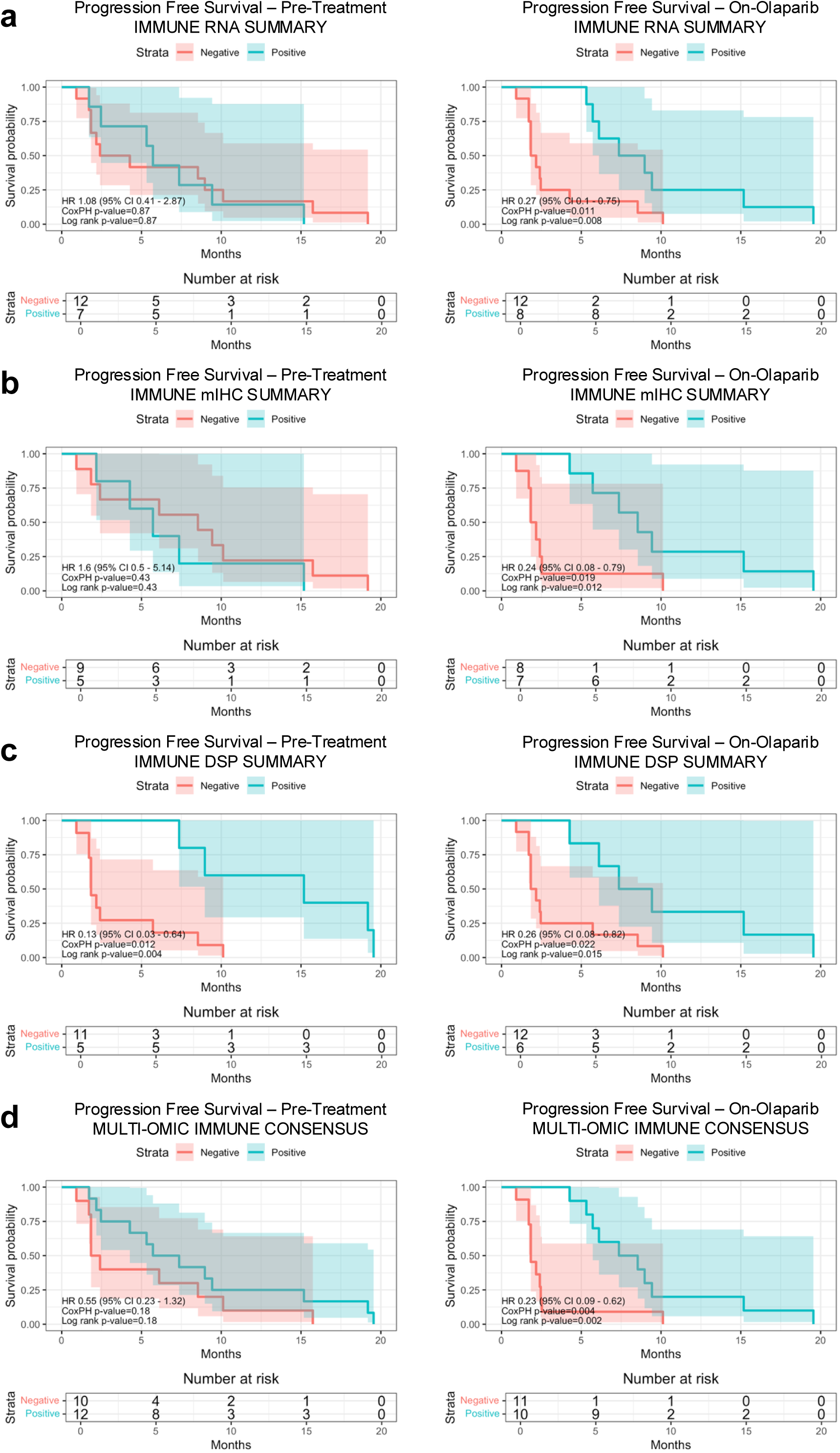
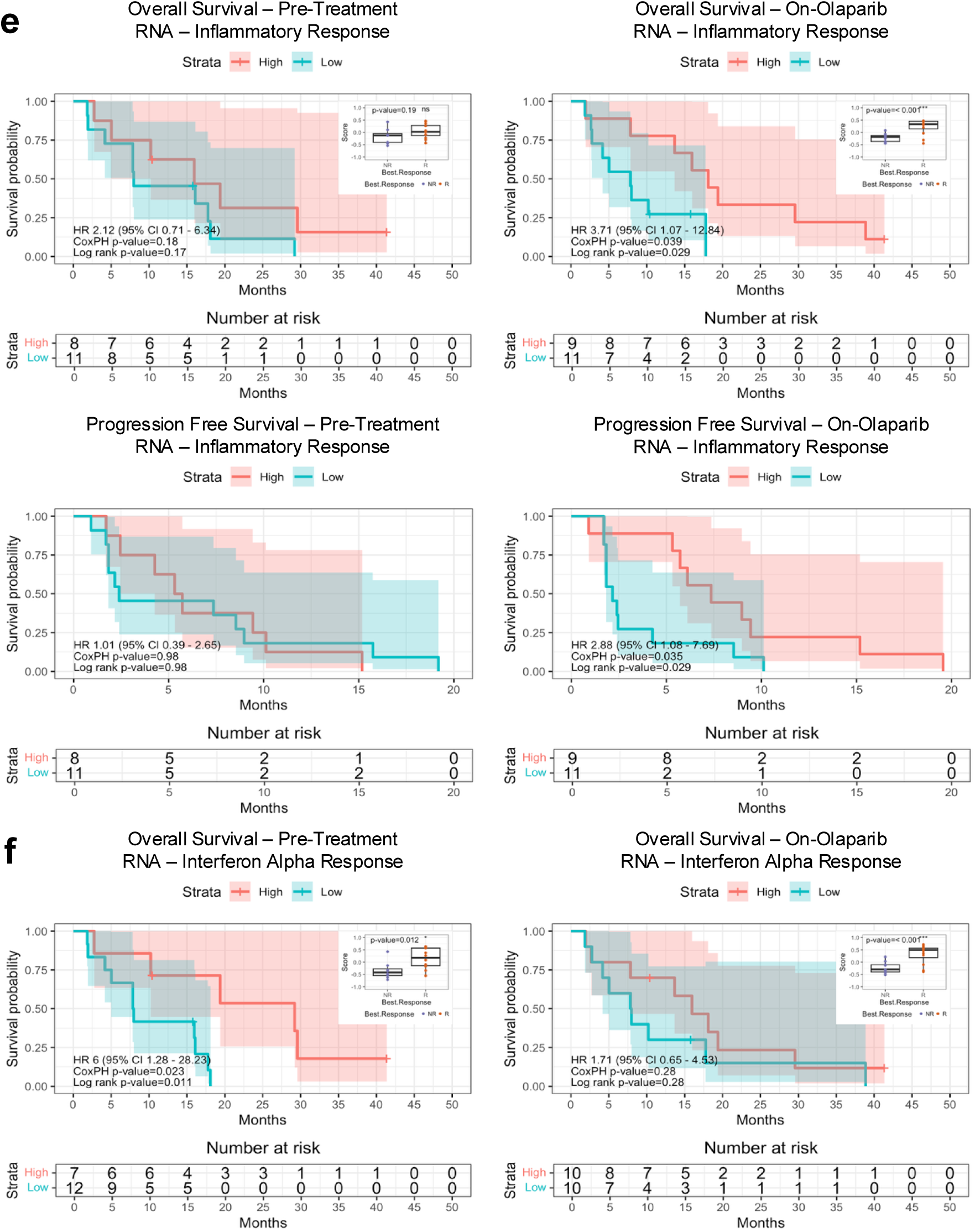
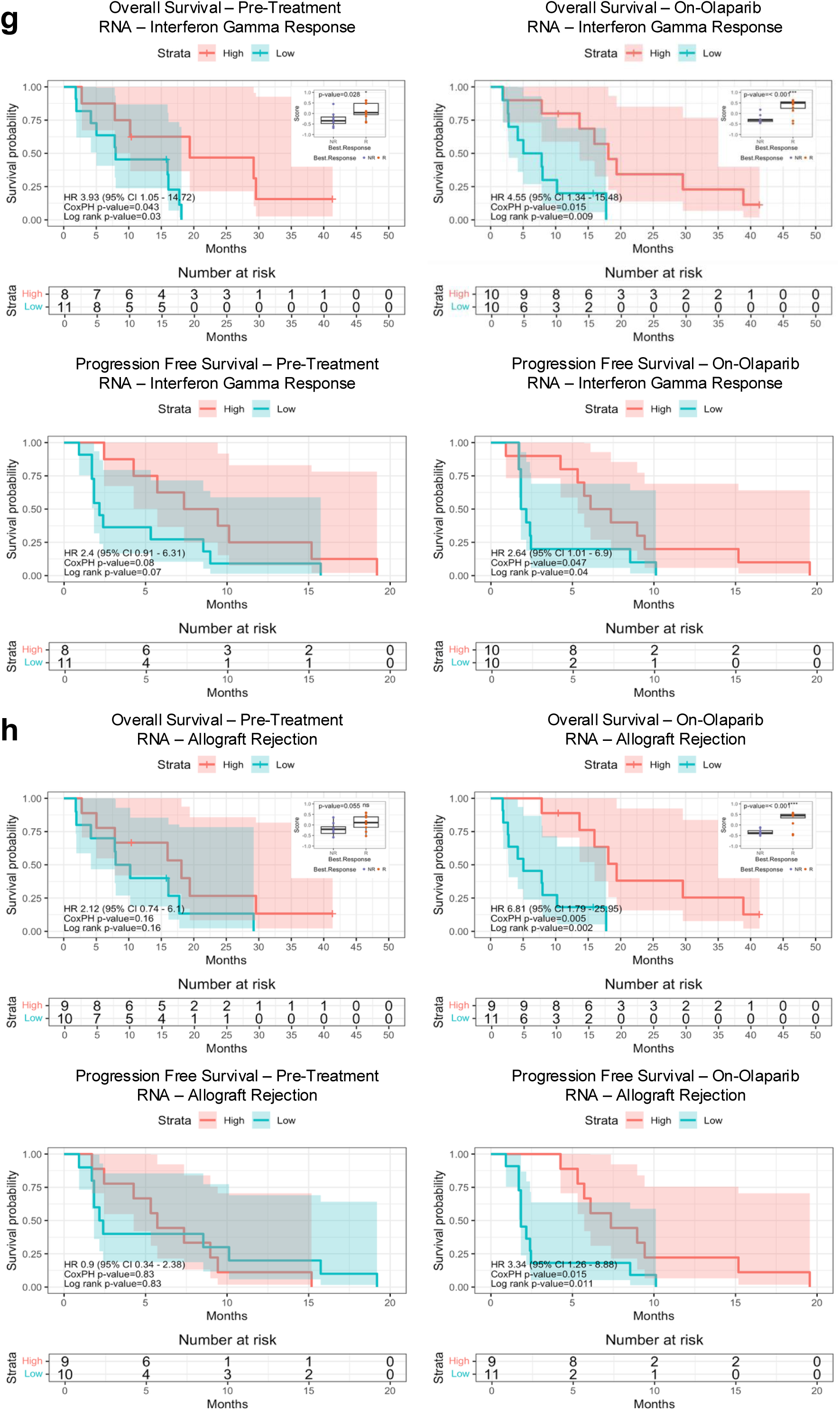
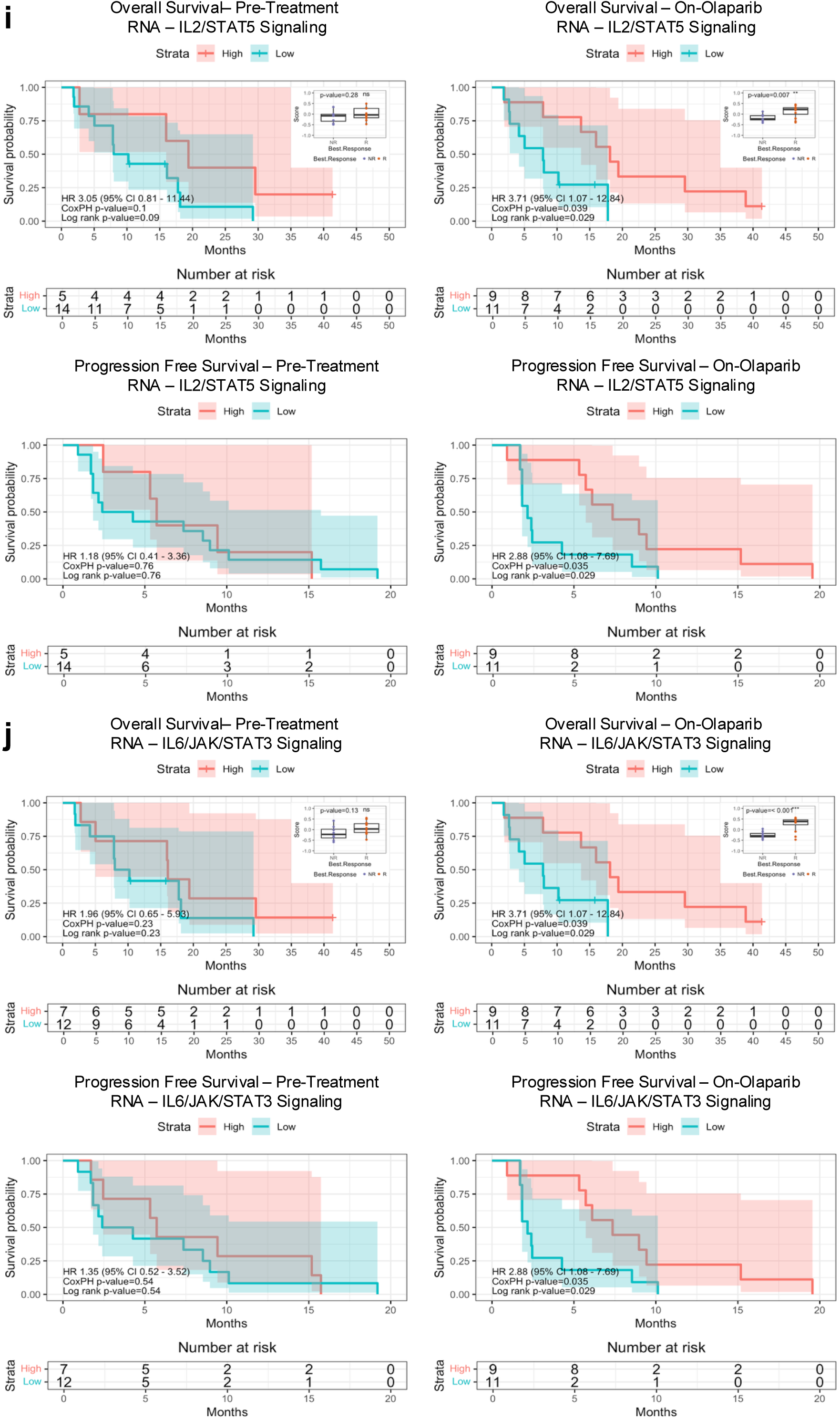
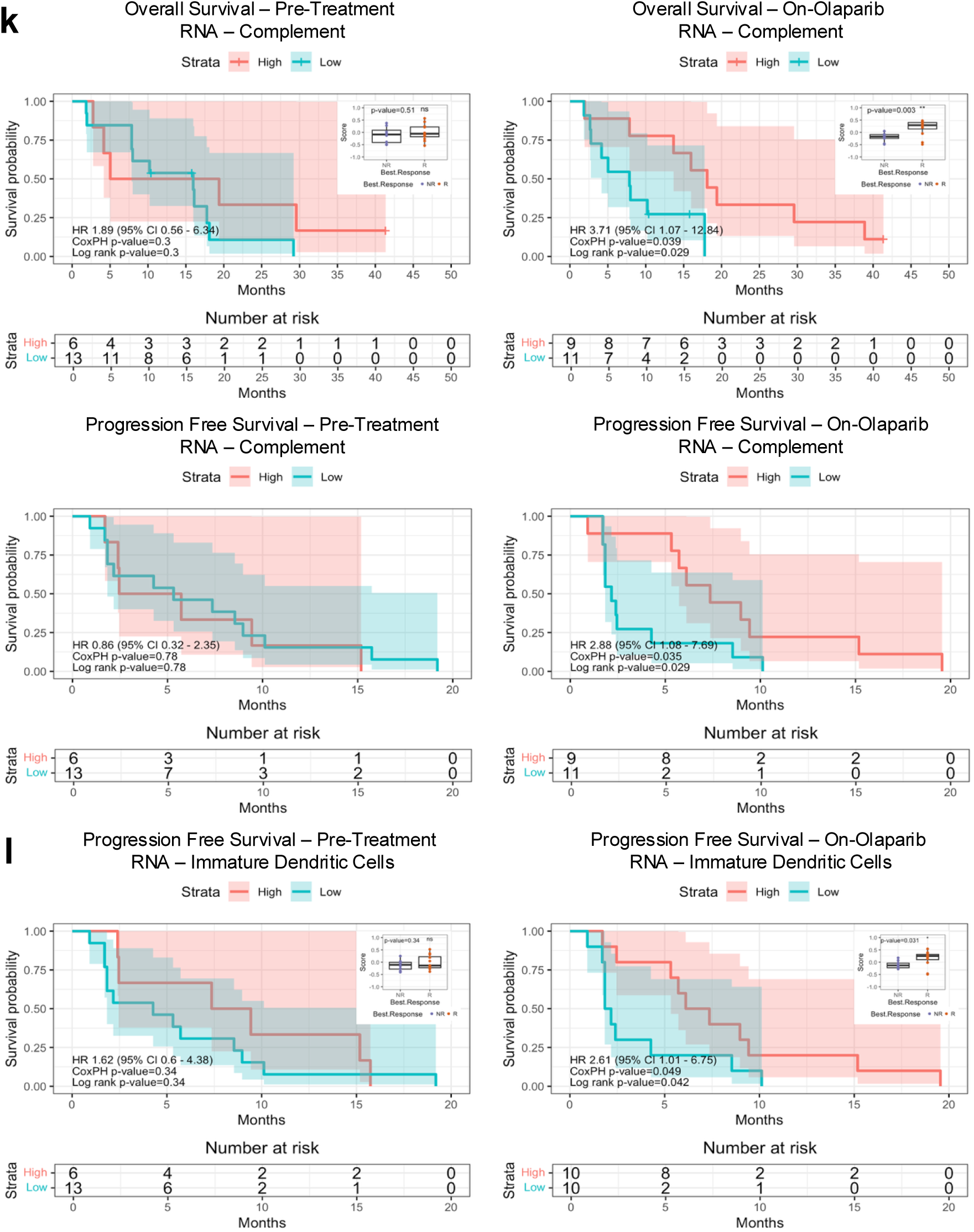
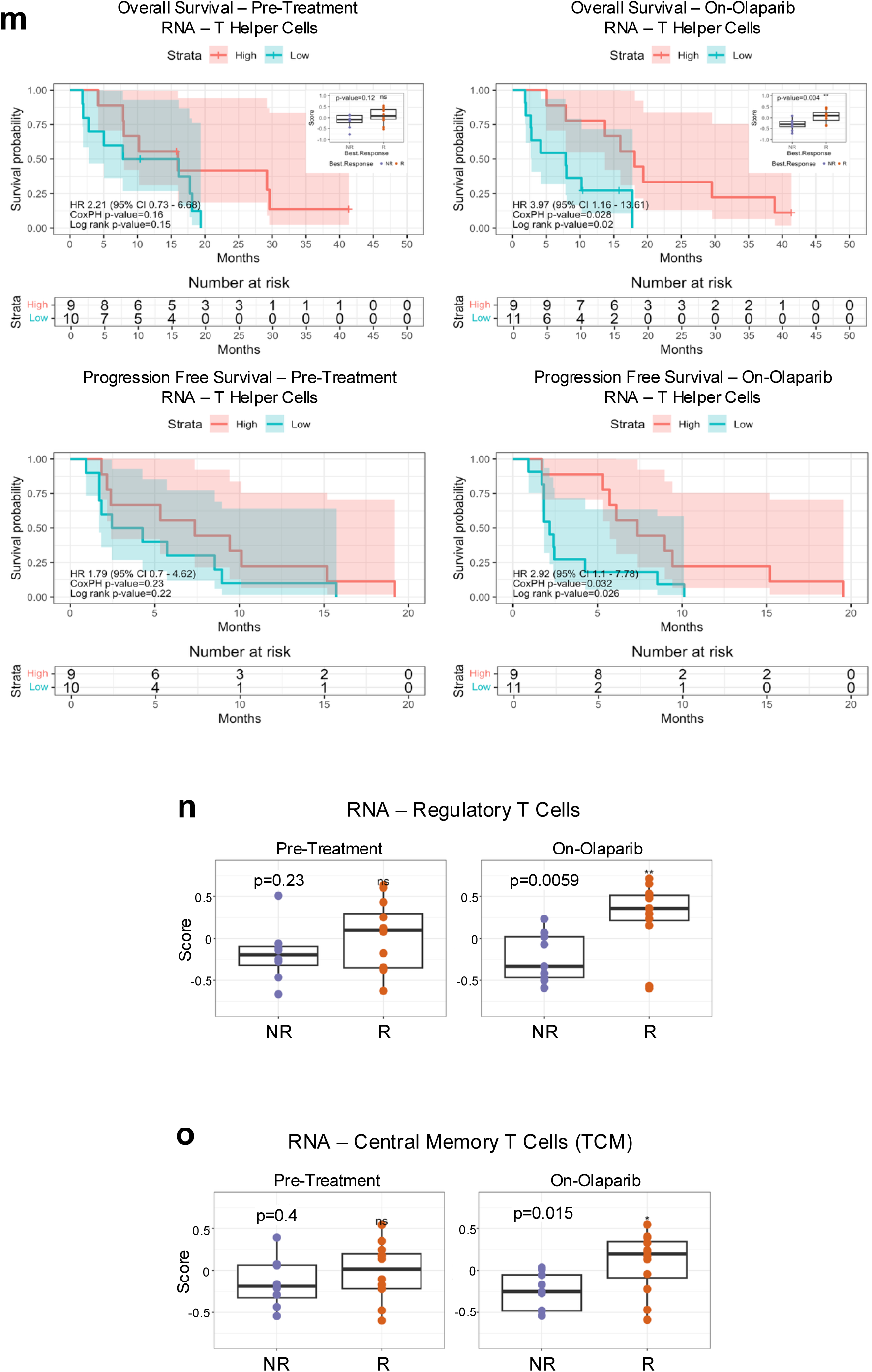
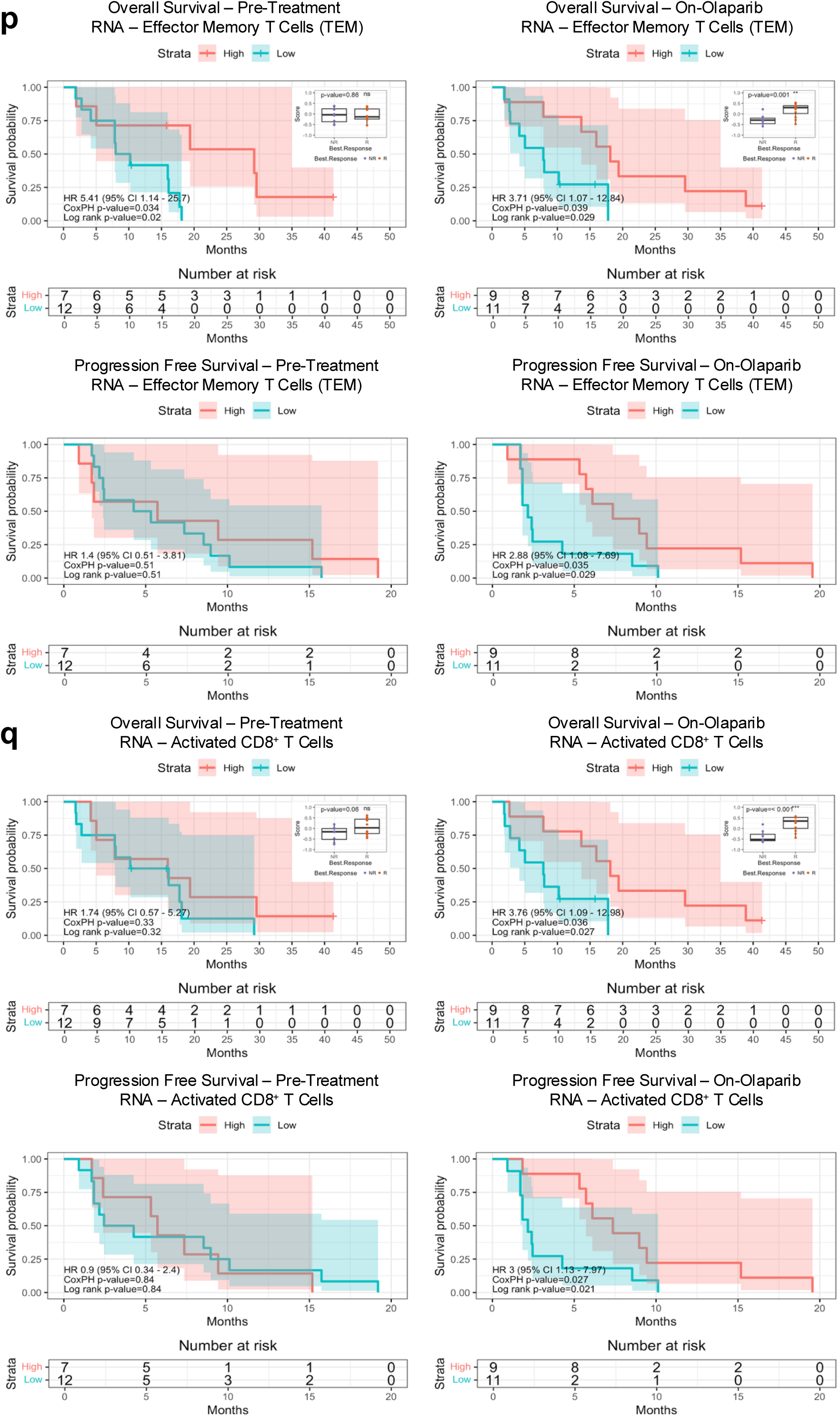
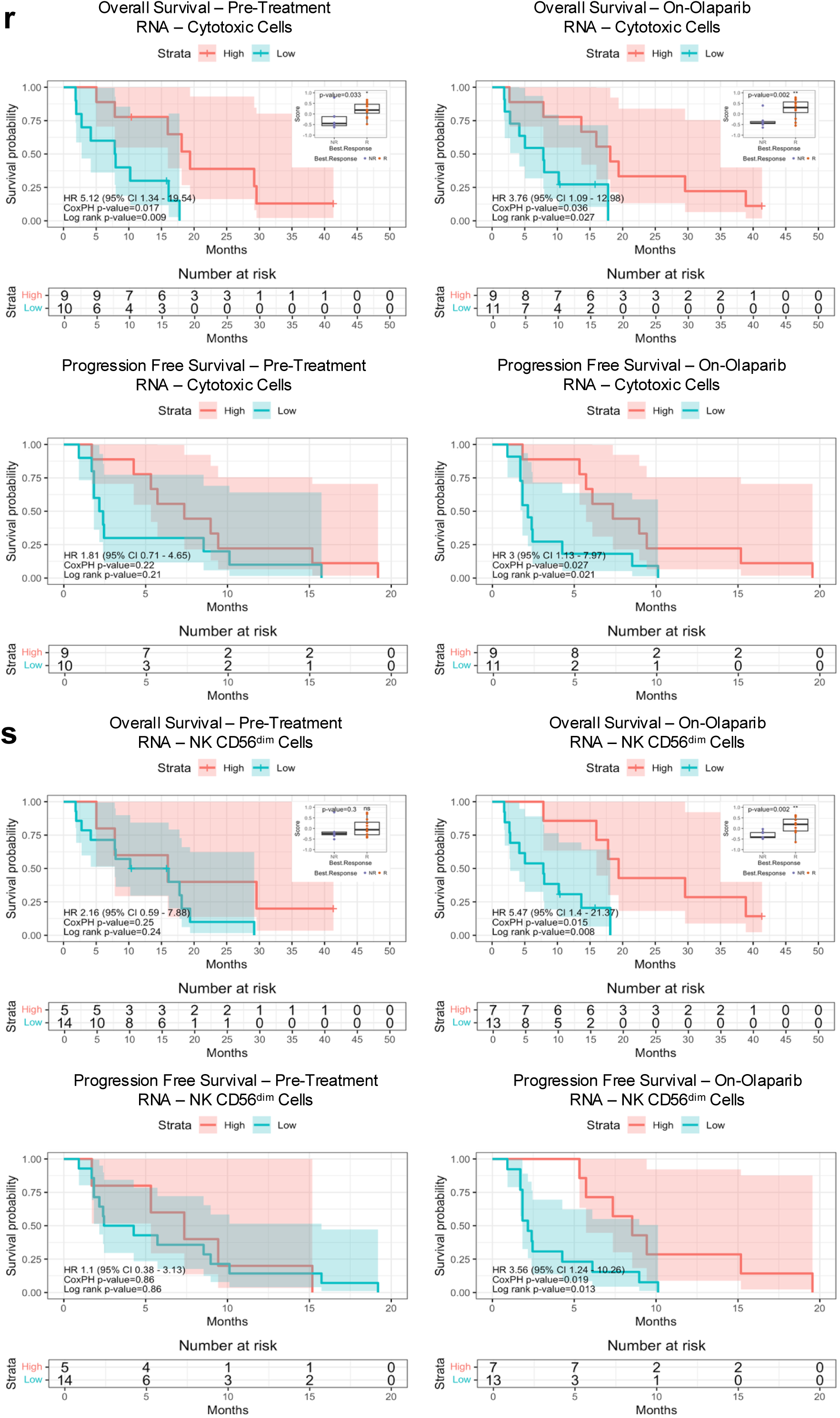
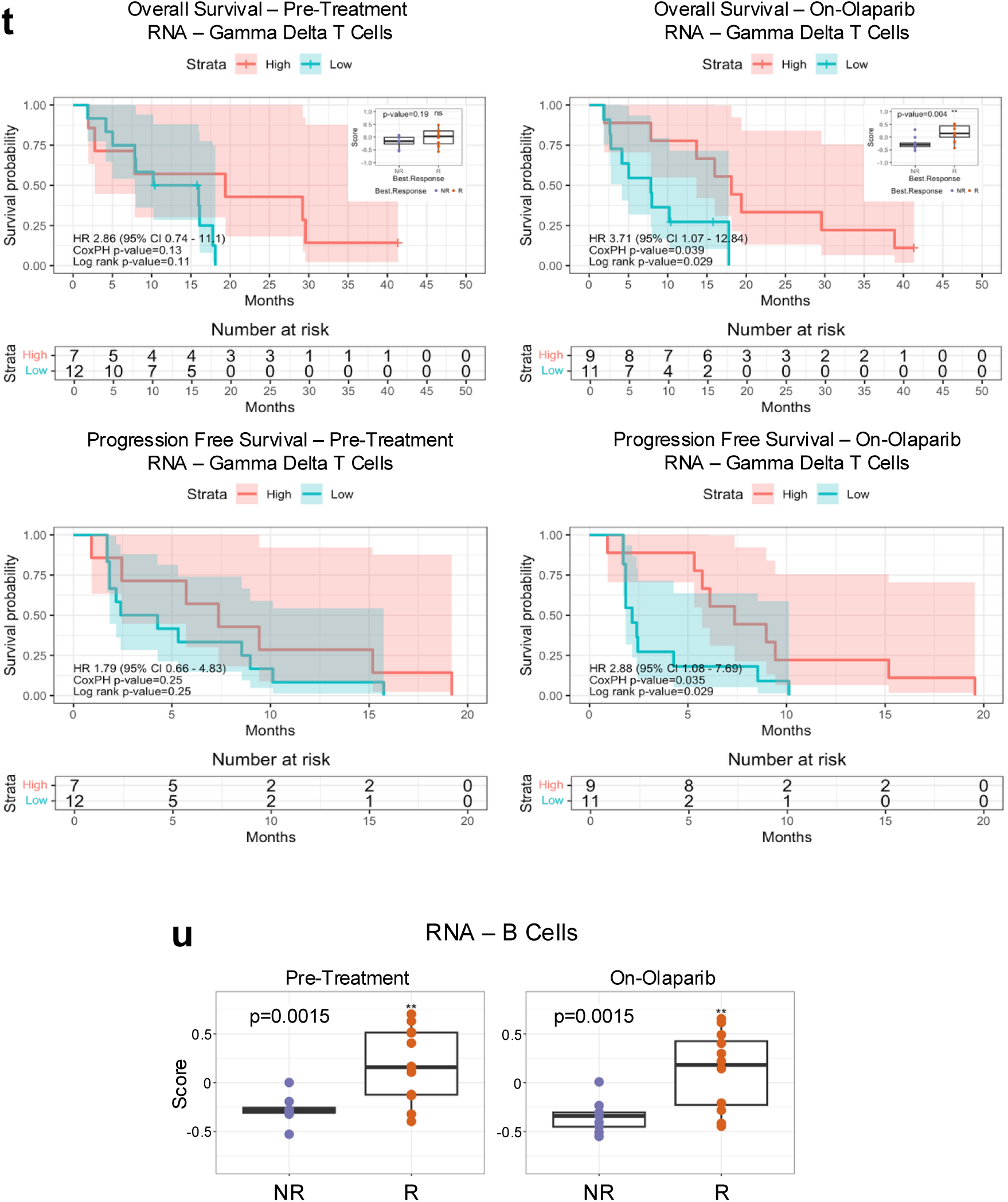
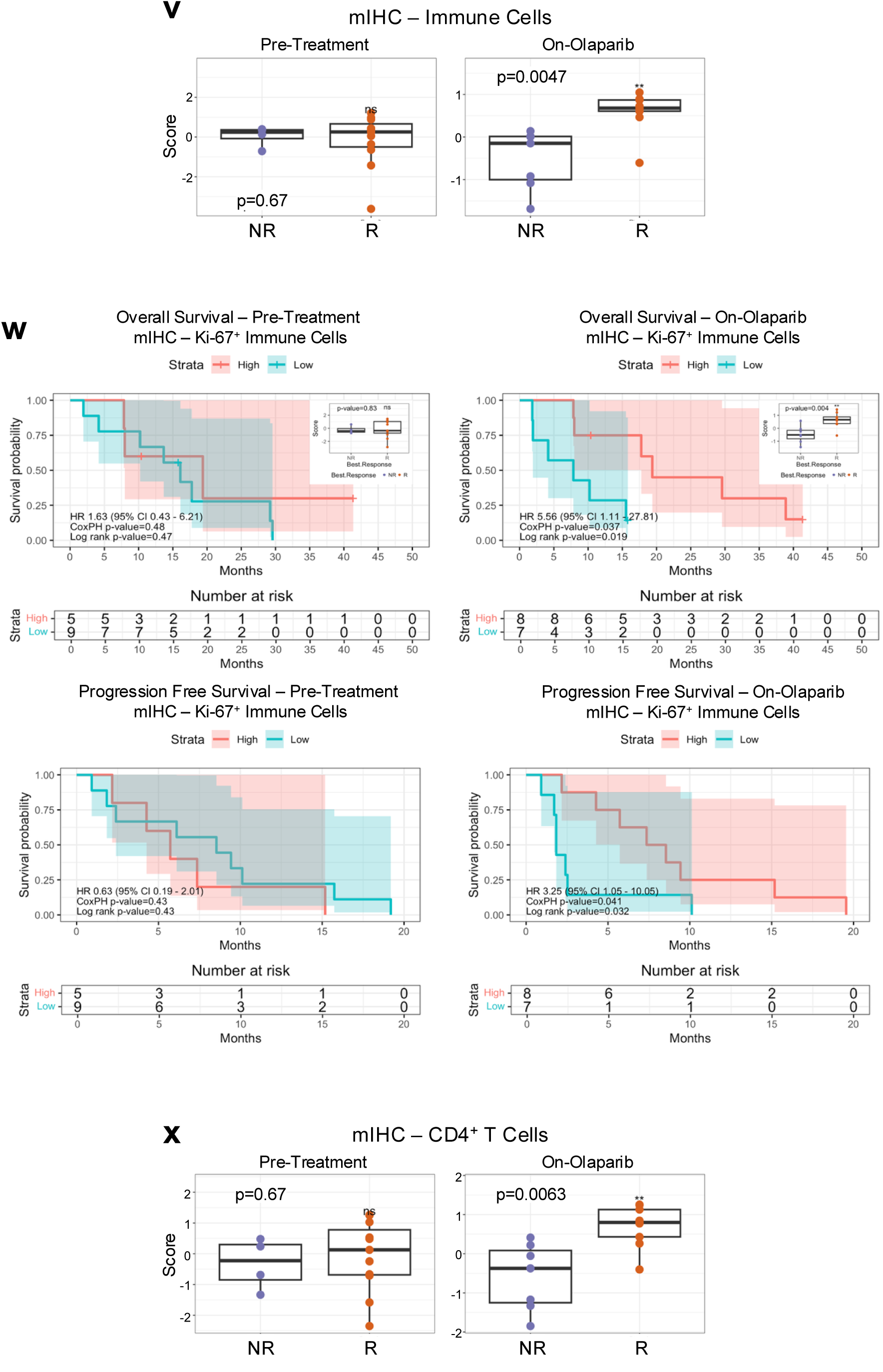
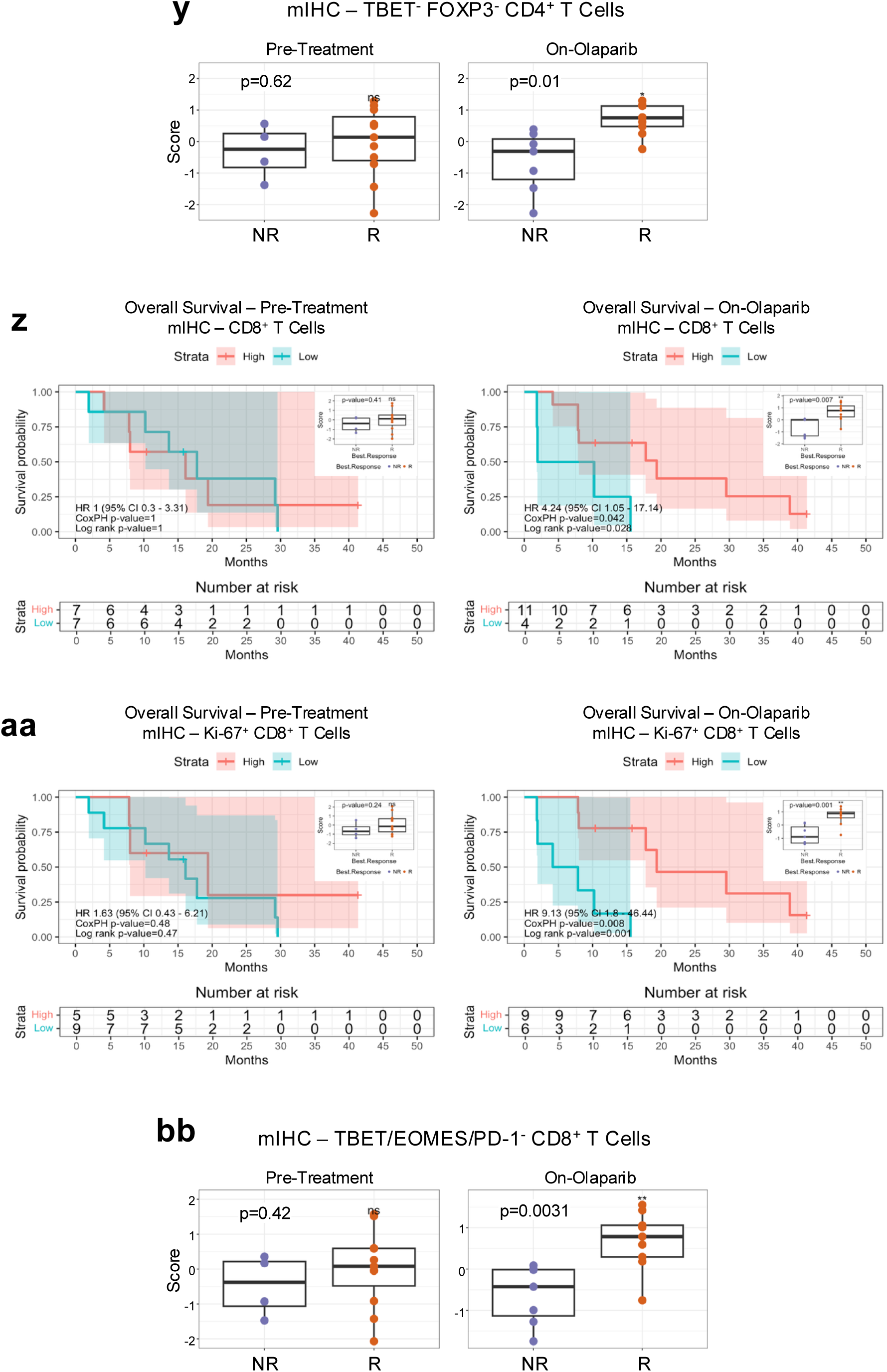
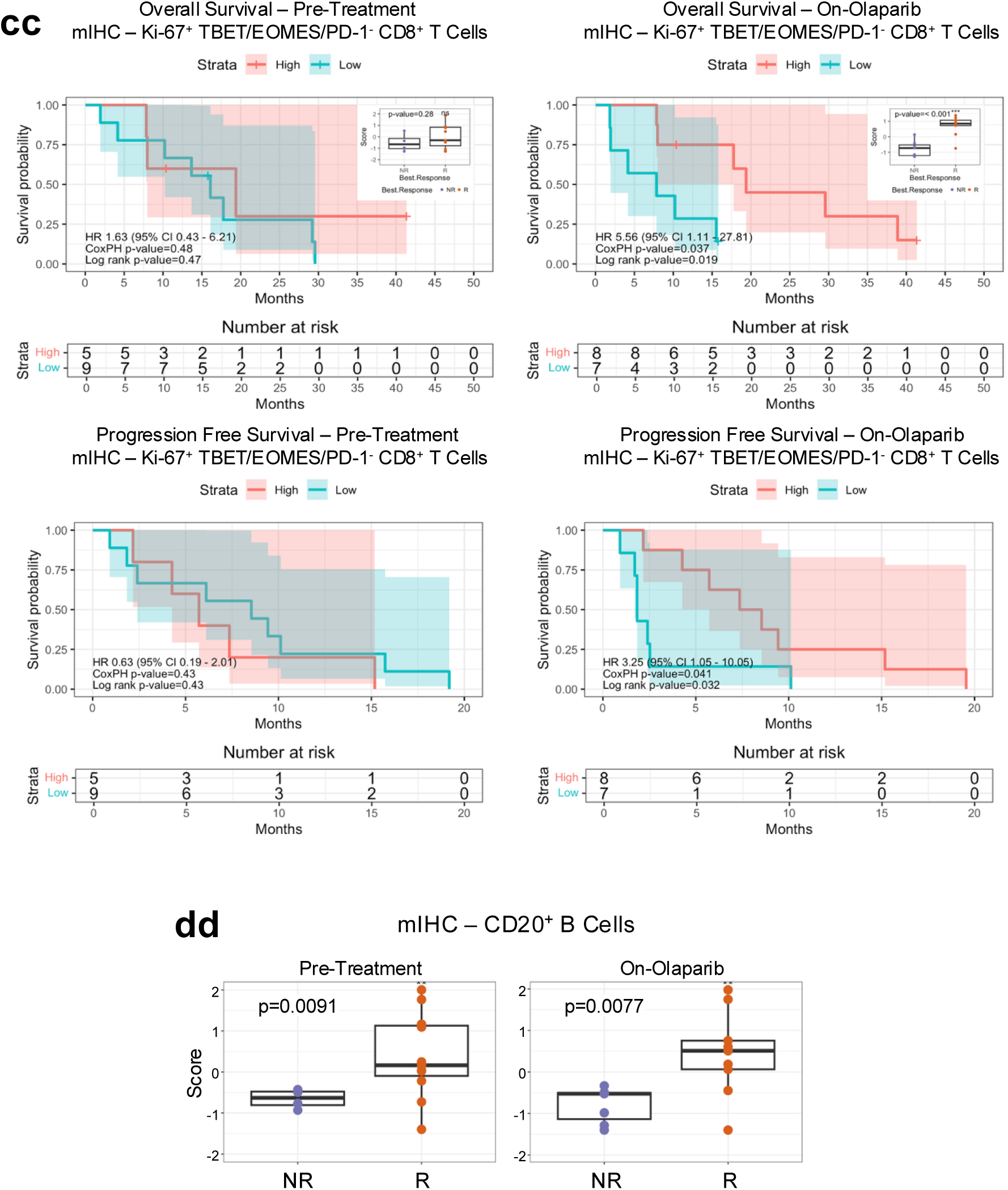
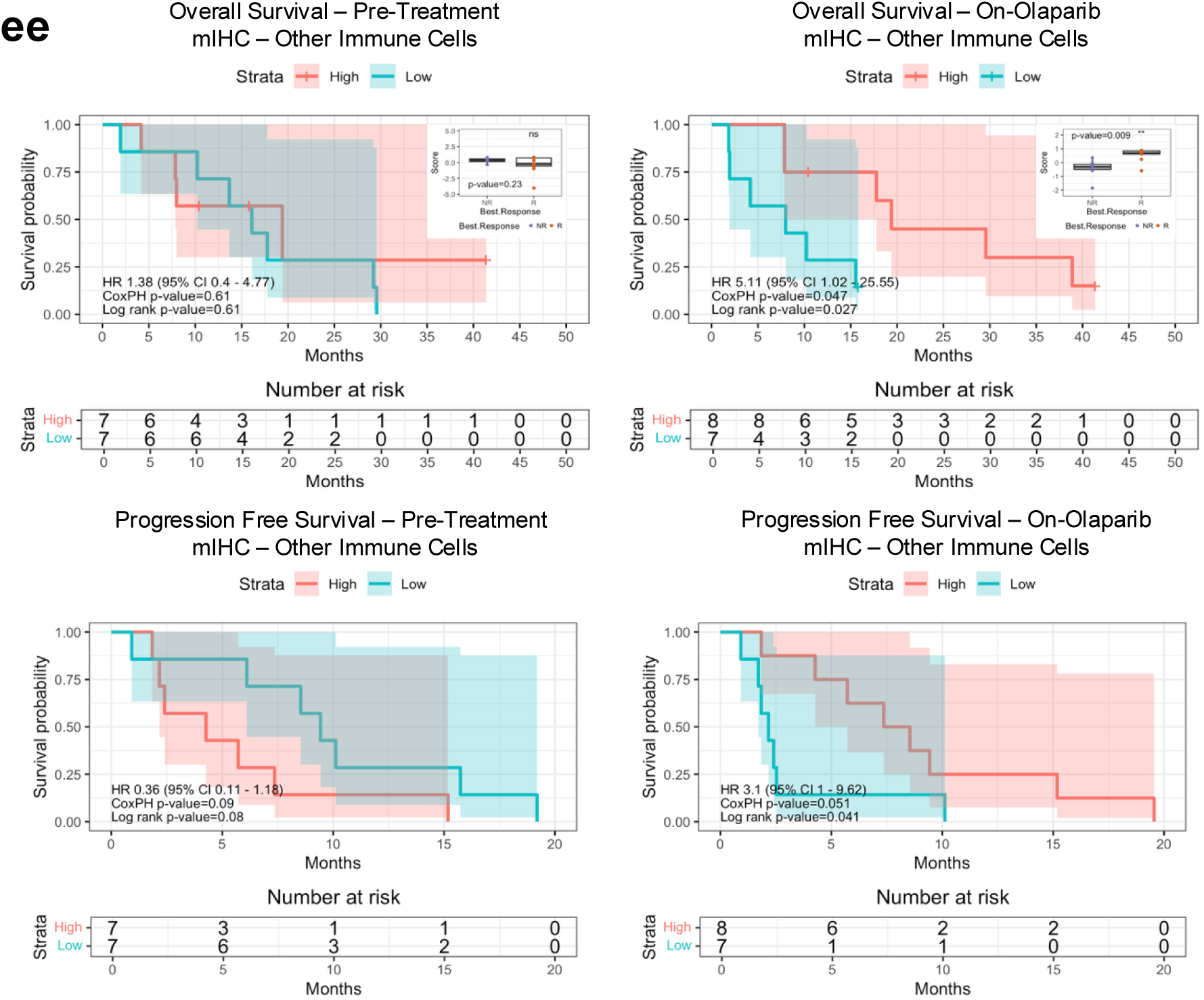
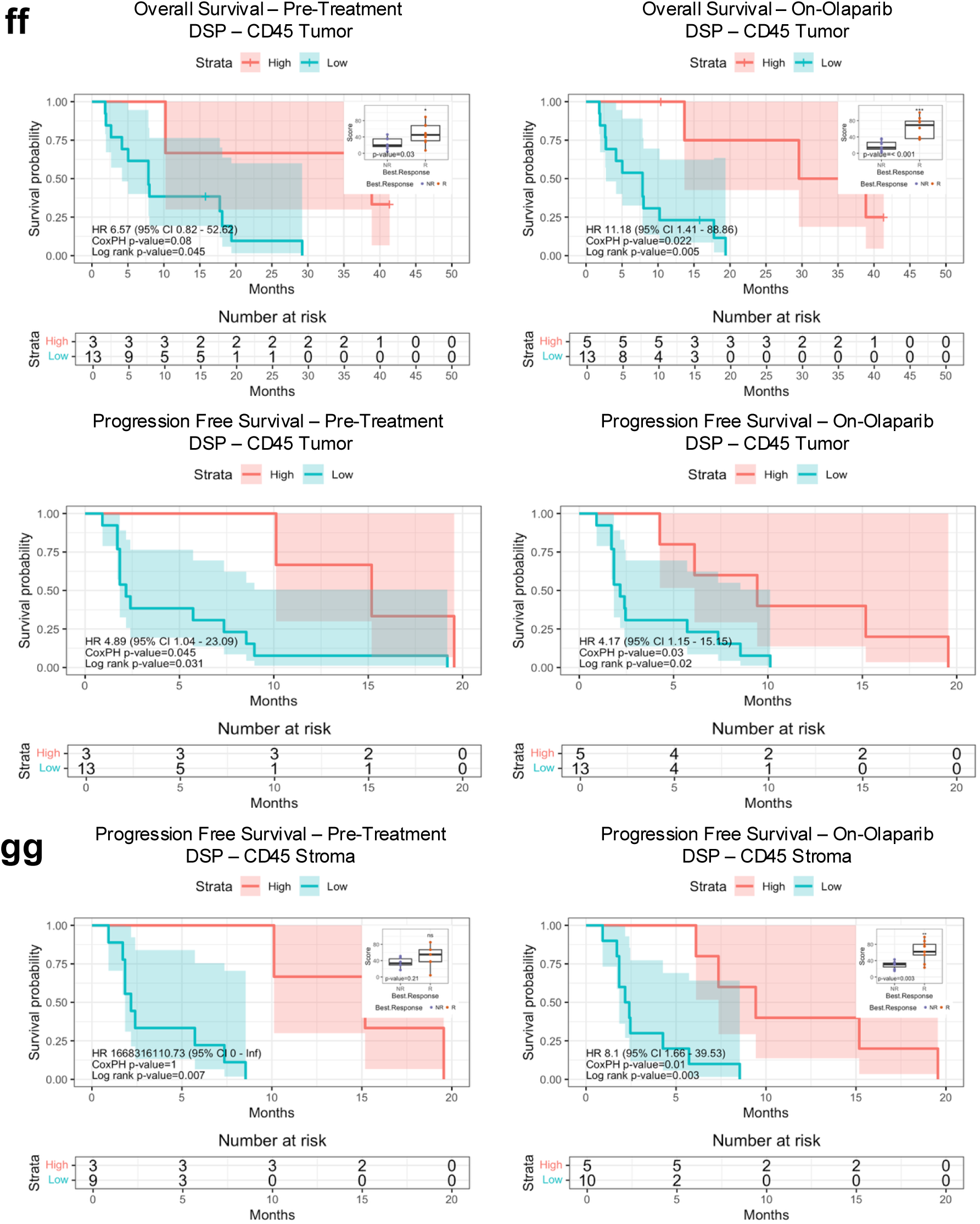
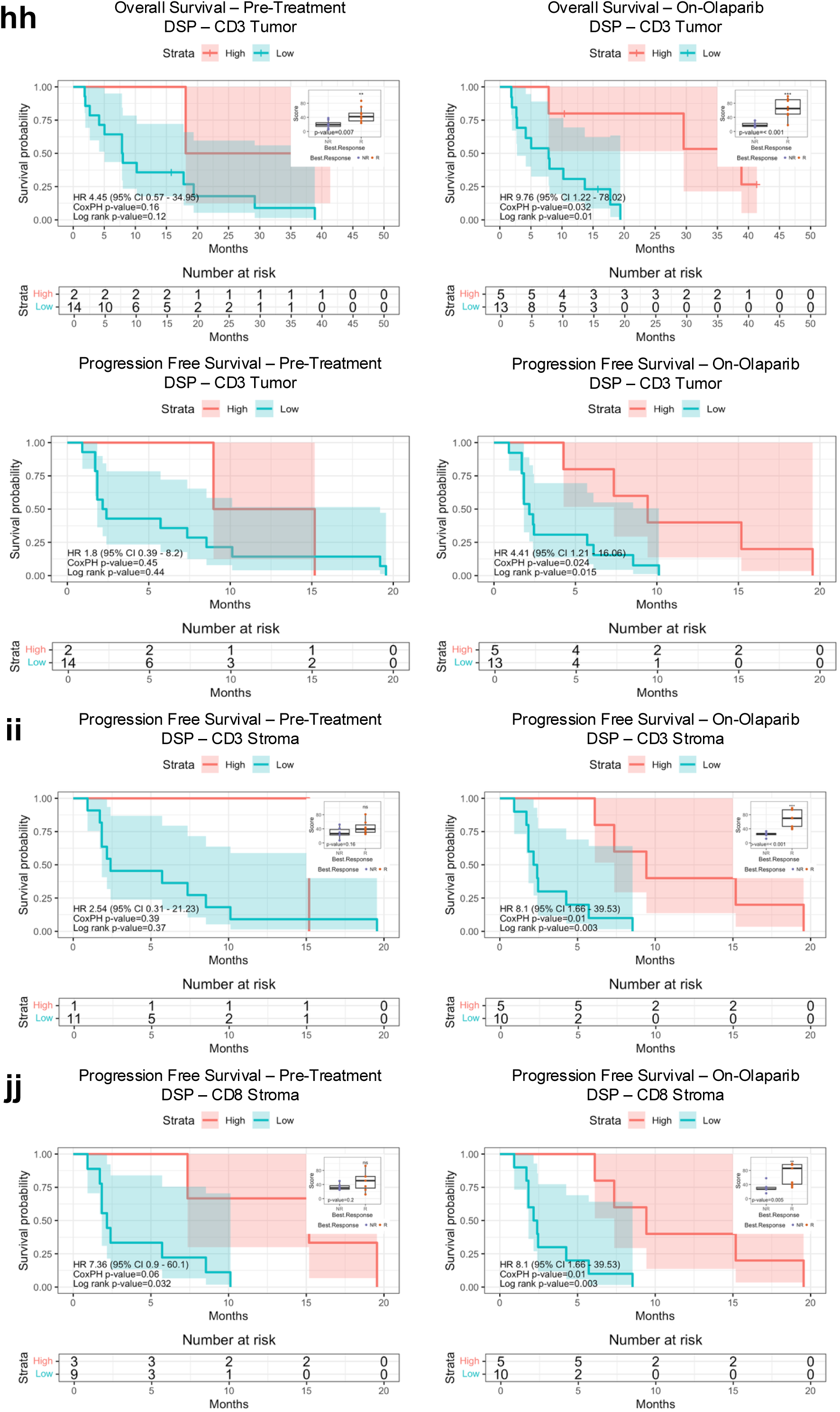
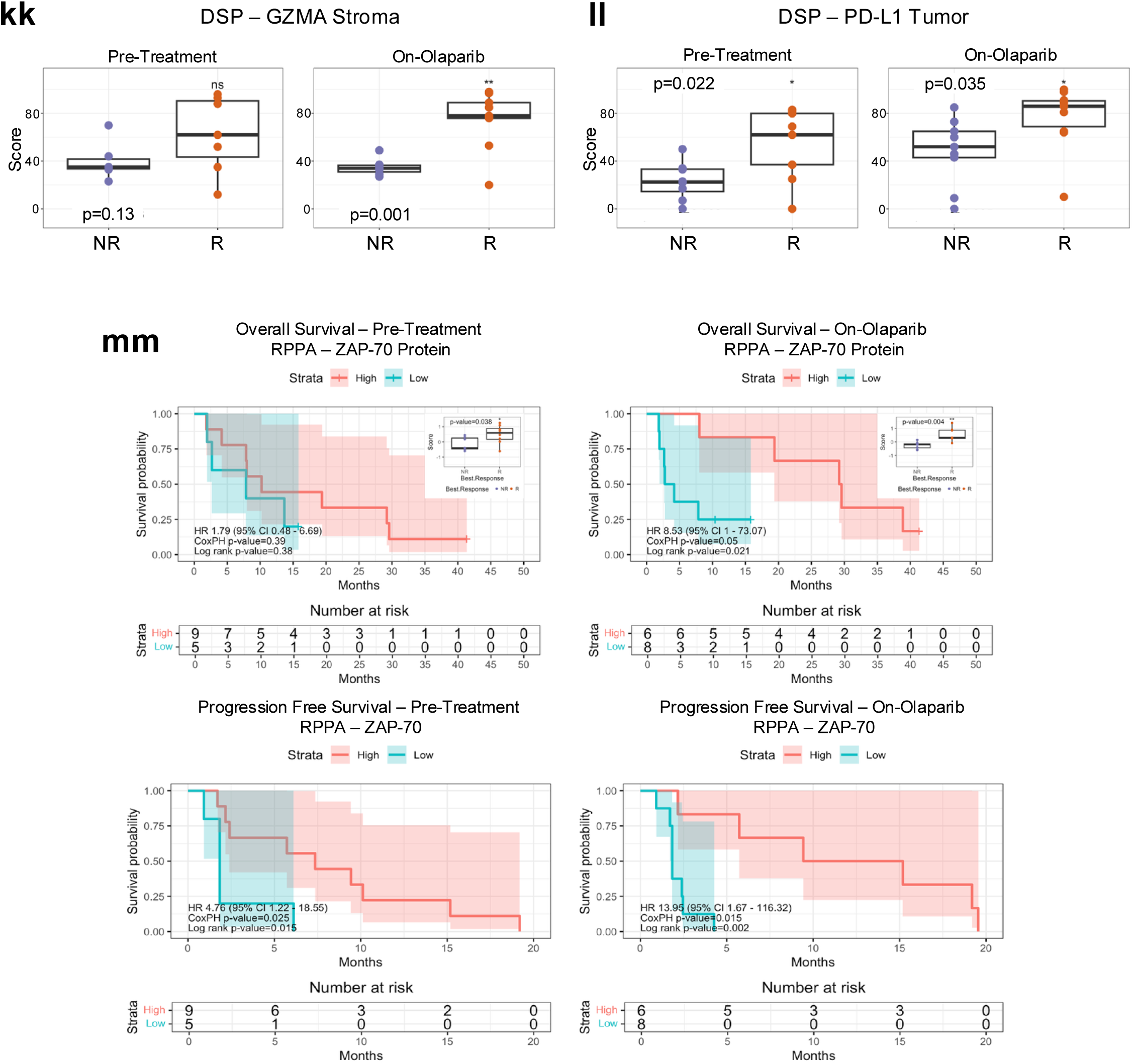
Microenvironment signatures of response to olaparib and durvalumab. **a-d)** *Multi-omic immune summary signatures*. Kaplan-Meier PFS in Pre-Treatment (left) and On-Olaparib (right) biopsies. Samples classified as Positive (blue) or Negative (red) based on predefined signature thresholds. HRs and p-values from CoxPH and log-rank tests are shown; shaded areas indicate 95% CI for the log survival probability. PFS measured from start of olaparib until end of olaparib plus durvalumab. Features shown achieved an F1 score >0.7 from random train/test split analysis and were significant (p <0.05) in pre-treatment and/or on-treatment samples (see Extended Data Table 1c). **a)** Immune RNA summary signature. Positive: ≥10 of 17 Immune RNA Signatures (Extended Data Figure 1c). **b)** Immune mIHC summary signature. Positive: ≥9 of 10 mIHC Cell Types (Extended Data Figure 1c). **c)** Immune DSP summary signature. Positive: ≥40% of Immune DSP proteins (Extended Data Figure 1c). **d)** Multi-Omic Immune Consensus signature. Positive: at least one positive summary signature (Extended Data Figure 1c). **e-u)** *Immune gene sets and signatures.* Kaplan-Meier OS and PFS in Pre-Treatment (left) and On-Olaparib (right) biopsies. Enrichment scores (treated as continuous variables) were categorized as High (≥0, red) and Low (<0, blue). HRs and p-values from CoxPH and log-rank tests are shown; shaded areas indicate 95% CI for the log survival probability. OS measured from olaparib start until death or data cutoff; right-censoring denoted by “+”. PFS as above. Boxplots: enrichment scores from from NR (purple) and R (orange); p-values from Student’s t-test. Features shown were identified by Boruta feature selection within MSigDB Cancer Hallmark or Inferred Immune Cell Type gene sets, achieved an F1 score >0.7 from random train/test split analysis, and were significant (p <0.05) in pre-treatment and/or on-treatment samples (see Extended Data Table 1c). **e)** Inflammatory Response (MSigDB Cancer Hallmark). Kaplan-Meier OS and PFS, and boxplots. **f)** Interferon Alpha Response (MSigDB Cancer Hallmark). Kaplan-Meier OS and PFS, and boxplots. **g)** Interferon Gamma Response (MSigDB Cancer Hallmark). Kaplan-Meier OS and PFS, and boxplots. **h)** Allograft Rejection (MSigDB Cancer Hallmark). Kaplan-Meier OS and PFS, and boxplots. **i)** IL2/STAT5 Signaling (MSigDB Cancer Hallmark). Kaplan-Meier OS and PFS, and boxplots. **j)** IL6/JAK/STAT3 Signaling (MSigDB Cancer Hallmark). Kaplan-Meier OS and PFS, and boxplots. **k)** Complement (MSigDB Cancer Hallmark). Kaplan-Meier OS and PFS, and boxplots. **l)** Immature Dendritic Cells (Inferred Immune Cell Type). Kaplan-Meier PFS, and boxplots. **m)** T Helper Cells (Inferred Immune Cell Type). Kaplan-Meier OS and PFS, and boxplots. **n)** Regulatory T Cells. Inferred Immune Cell Type). Boxplots. **o)** Central Memory T Cells (TCM; Inferred Immune Cell Type). Boxplots. **p)** Effector Memory T Cells (TEM; Inferred Immune Cell Type). Kaplan-Meier OS and PFS, and boxplots. **q)** Activated CD8^+^ T Cells (Inferred Immune Cell Type). Kaplan-Meier OS and PFS, and boxplots. **r)** Cytotoxic Cells (Inferred Immune Cell Type). Kaplan-Meier OS and PFS, and boxplots. **s)** NK CD56^dim^ Cells (Inferred Immune Cell Type). Kaplan-Meier OS and PFS, and boxplots. **t)** Gamma Delta T Cells (Inferred Immune Cell Type). Kaplan-Meier OS and PFS, and boxplots. **u)** B Cells (Inferred Immune Cell Type). Boxplots. **v-ee)** *mIHC immune cell lineages and states.* Kaplan-Meier OS and PFS in Pre-Treatment (left) and On-Olaparib (right) biopsies. Cell phenotype densities (treated as continuous variables) discretized as High (≥ 0, red) and Low (< 0, blue). HRs and p-values from CoxPH and log-rank tests are shown; shaded areas indicate 95% CI for the log survival probability. OS and PFS were measured as above. Boxplots: cell phenotype densities in biopsies from NR (purple) and R (orange); p-values from Student’s t-test. Features shown were identified by Boruta feature selection, achieved an F1 score >0.7 from random train/test split analysis, and were significant (p <0.05) in pre-treatment and/or on-treatment samples (see Extended Data Table 1c). **v)** Immune Cells. Boxplots. **w)** Ki-67^+^ Immune Cells. Kaplan-Meier OS and PFS and boxplots. **x)** CD4^+^ T Cells. Boxplots. **y)** TBET^-^ FOXP3^-^ CD4^+^ T Cells. Boxplots. **z)** CD8^+^ T Cells. Kaplan-Meier OS and boxplots. **aa)** Ki-67^+^ CD8^+^ T Cells. Kaplan-Meier OS and boxplots. **bb)** TBET/EOMES/PD-1^-^ CD8^+^ T Cells. Boxplots. **cc)** Ki-67^+^ TBET/EOMES/PD-1^-^ CD8^+^ T Cells. Kaplan-Meier OS and PFS and boxplots. **dd** CD20^+^ B Cells. Boxplots. **ee)** Other Immune Cells. Kaplan-Meier OS and PFS and boxplots. **ff-ll)** *DSP immune protein expression.* Kaplan-Meier OS and PFS in Pre-Treatment (left) and On-Olaparib (right) biopsies. Protein levels in both tumor and stroma regions of interest categorized as High (≥60 z-score percentile, red) and Low (<60 percentile, blue). HR and p-values from CoxPH and log-rank tests are shown; shaded areas indicate 95% CI for the log survival probability. OS and PFS were measured as above. Boxplots: protein levels in biopsies from NR (purple) and R (orange); p-values from Student’s t-test. Features shown were identified by Boruta feature selection, achieved an F1 score >0.7 from random train/test split analysis, and were significant (p <0.05) in pre-treatment and/or on-treatment samples (see Extended Data Table 1c). **ff)** CD45 Tumor. Kaplan-Meier OS and PFS and boxplots. **gg)** CD45 Stroma. Kaplan-Meier PFS and boxplots. **hh)** CD3 Tumor. Kaplan-Meier OS and PFS and boxplots. **ii)** CD3 Stroma. Kaplan-Meier PFS and boxplots. **jj)** CD8 Stroma. Kaplan-Meier PFS and boxplots. **kk)** GZMA Stroma. Boxplots. **ll)** PD-L1 Tumor. Boxplots. **mm)** *ZAP-70 protein expression from RPPA.* Kaplan-Meier OS and PFS in Pre-Treatment (left) and On-Olaparib (right) biopsies. Scaled and normalized immune protein expression categorized as High (MAD z-score ≥0, red) and Low (MAD z-score <0, blue). HRs and p-values from CoxPH and log-rank tests are shown; shaded areas indicate 95% CI for the log survival probability. OS and PFS were measured as above. Boxplots: protein expression in biopsies from NR (purple) and R (orange); p-values from Student’s t-test.

**Extended Data Figure 4.**
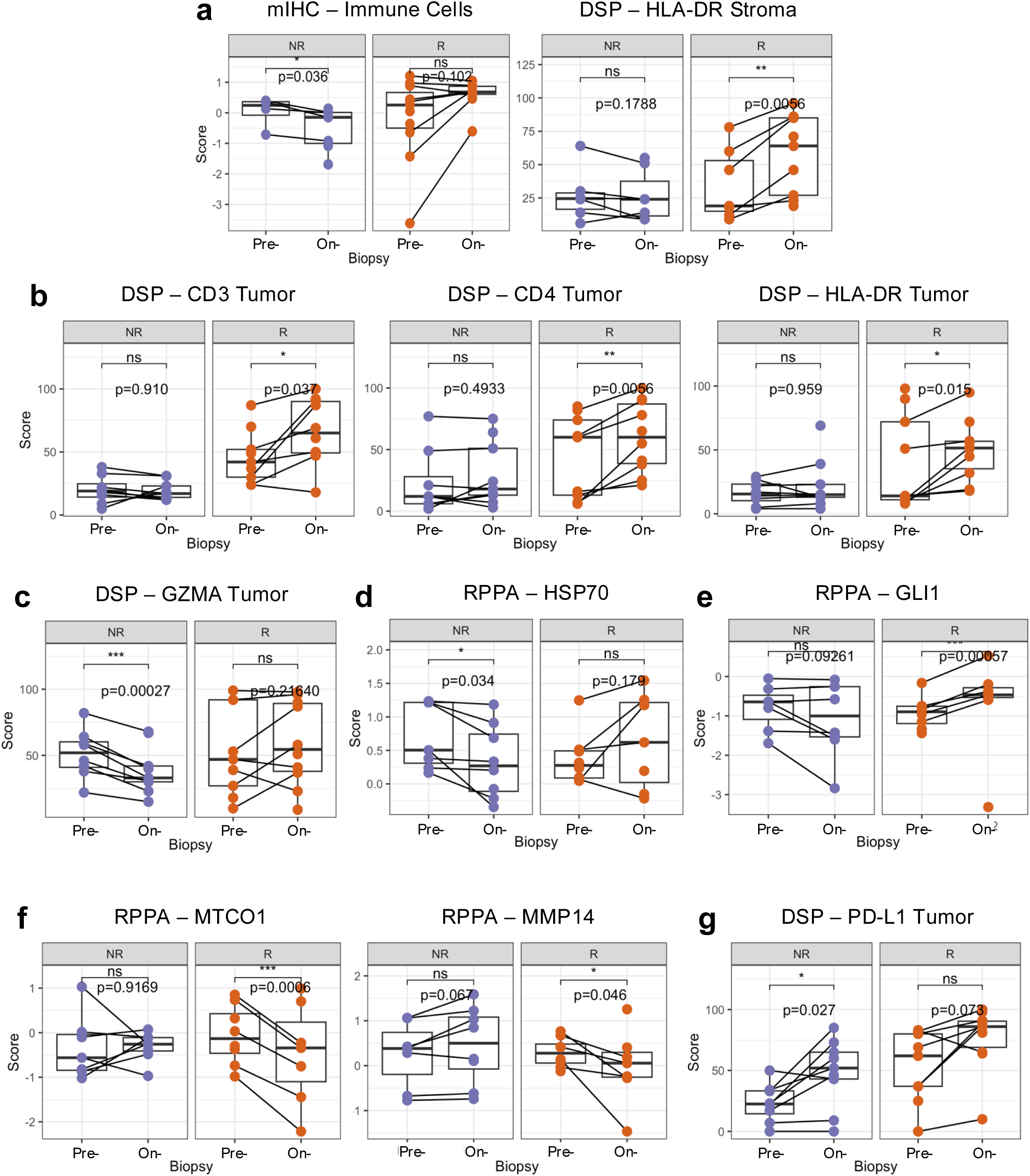
Olaparib-induced response markers and signatures. **a-g)** *Changes following olaparib treatment.* Boxplots comparing feature scores in Pre-Treatment (Pre-) and On-Olaparib (On-) biopsies, separated by non-responders (NR, purple, left) and responders (R, orange, right). Lines connect paired Pre- and On- samples from the same participant. P-values from paired Student’s t-test for Pre- vs. On- biopsies. Markers from Fig. 4 and p<0.05 are shown. **a)** Immune markers increased in responders, decreased in non-responders (see Fig. 4a). **b)** Immune markers increased in responders, unchanged in non-responders (see Fig. 4b). **c)** Immune markers unchanged in responders, decreased in non-responders (see Fig. 4c). **d)** Malignant cell markers increased in responders, decreased in non-responders (see Fig. 4e). **e)** Malignant cell markers increased in responders, unchanged in non-responders (see Fig. 4f). **f)** Malignant cell markers decreased in responders, increased in non-responders (see Fig. 4g). **g)** PD-L1 Tumor DSP.

**Extended Data Figure 5.**
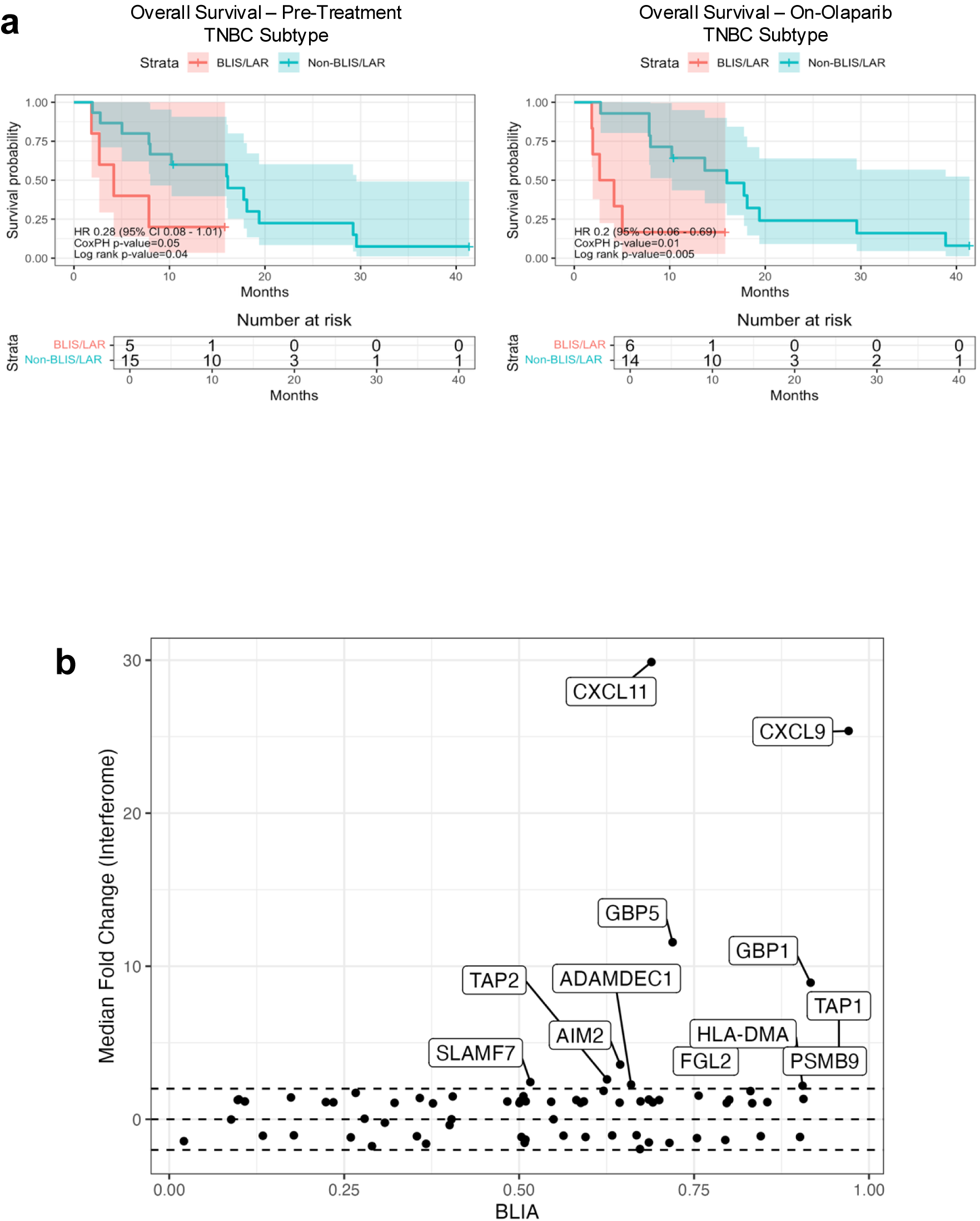
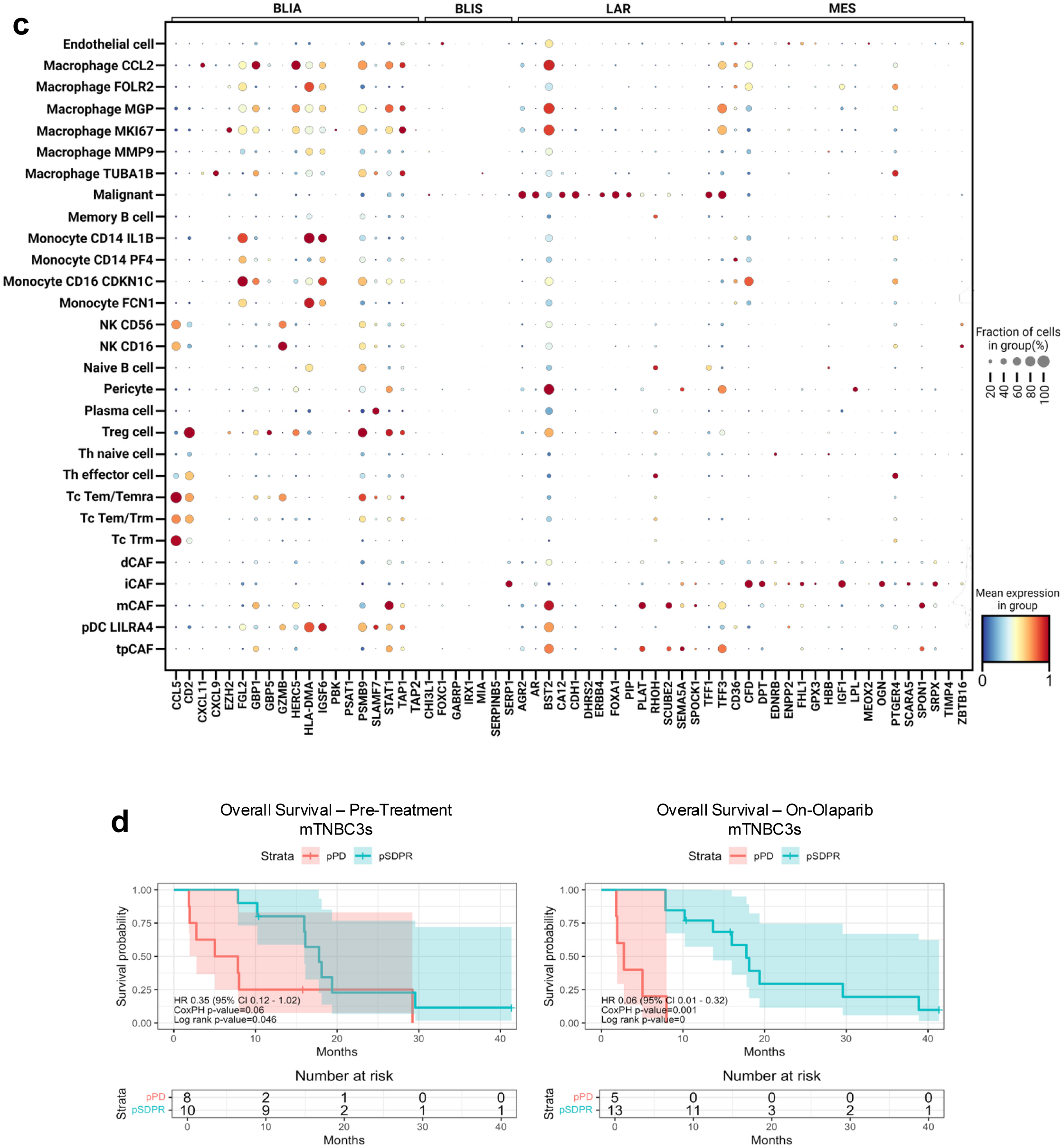
TNBC subtype correlates of response to olaparib and durvalumab. **a)** *TNBC subtypes.* Kaplan-Meier OS stratified by TNBC RNA subtype (BLIS/LAR, red; Non-BLIS/LAR, blue) in Pre-Treatment (left) and On-Olaparib (right) biopsies. HR and p-values from CoxPH and log-rank tests are shown; shaded areas indicate 95% CI for the log survival probability. OS measured from olaparib start until death or data cutoff; right-censoring denoted by “+”. **b)** *BLIA interferon-regulated genes.* The Interferome web tool (http://www.interferome.org), a database of interferon-regulated genes, was used to identify a subset of highly ranked genes (median Interferome gene change >2, dashed lines, Y-axis) associated with the BLIA subtype (>0.5, X-axis). **c)** *TNBC subtype genes within cell types.* Dot plot visualizing expression of representative marker genes (x-axis, bottom) within TNBC subtypes (x-axis, top) across distinct cell type clusters (y-axis). Data are from a single-cell RNAseq breast cancer dataset (see *Methods*). Relative gene expression levels are represented by the color scale, and the fraction of cells within each cell type expressing the gene is represented by the dot size. The plot highlights differential expression patterns of key TNBC markers within each cell cluster, providing insight into the heterogeneity and distribution of TNBC-related gene expression profiles at the single-cell level. *mTNBC3s gene signature.* Kaplan-Meier OS stratified by mTNBC3s^54^ predicted non-responders (pPD, red) and predicted responders (pSDPR, blue). OS was measured as above.

**Extended Data Table 1.**
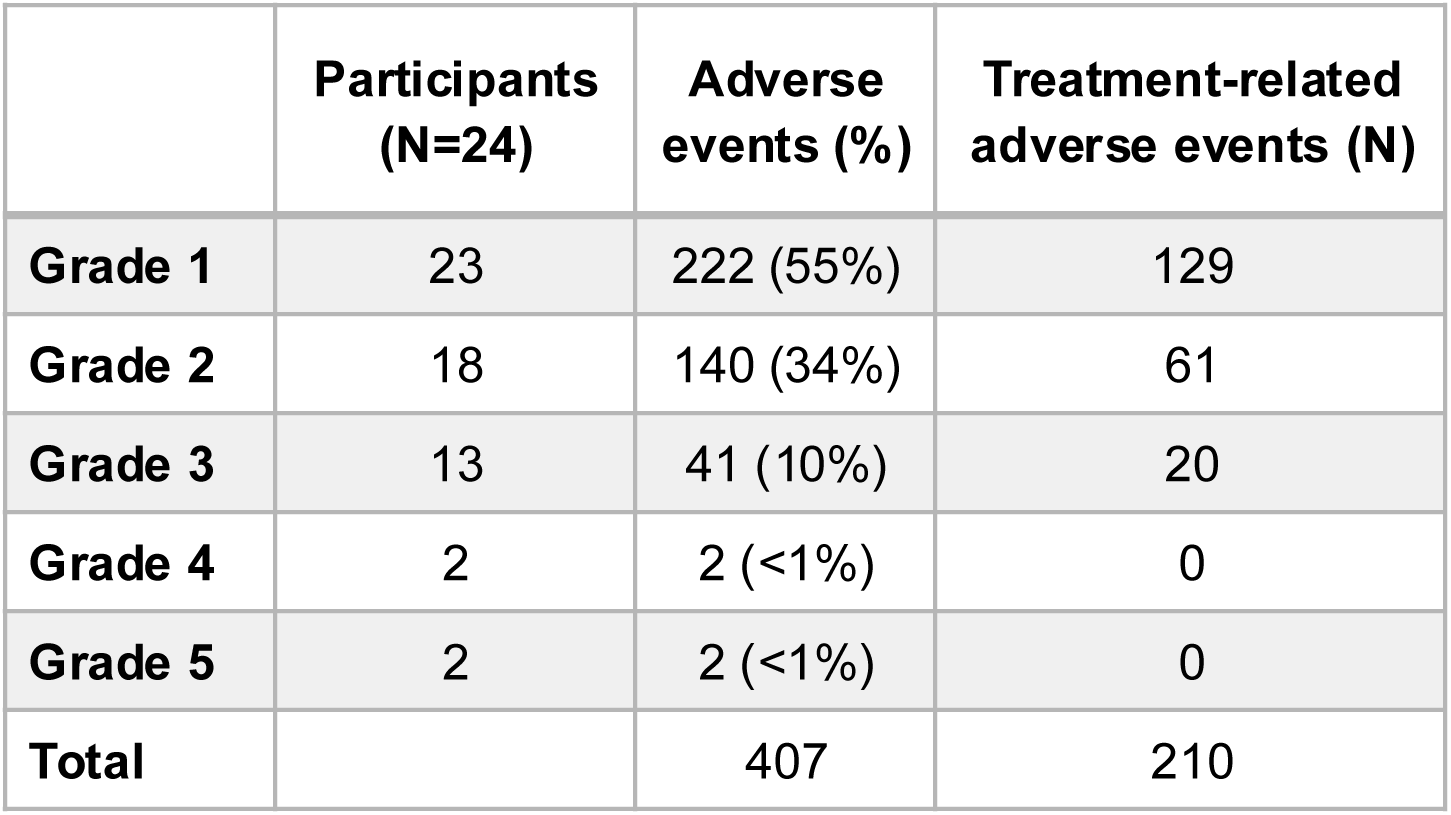
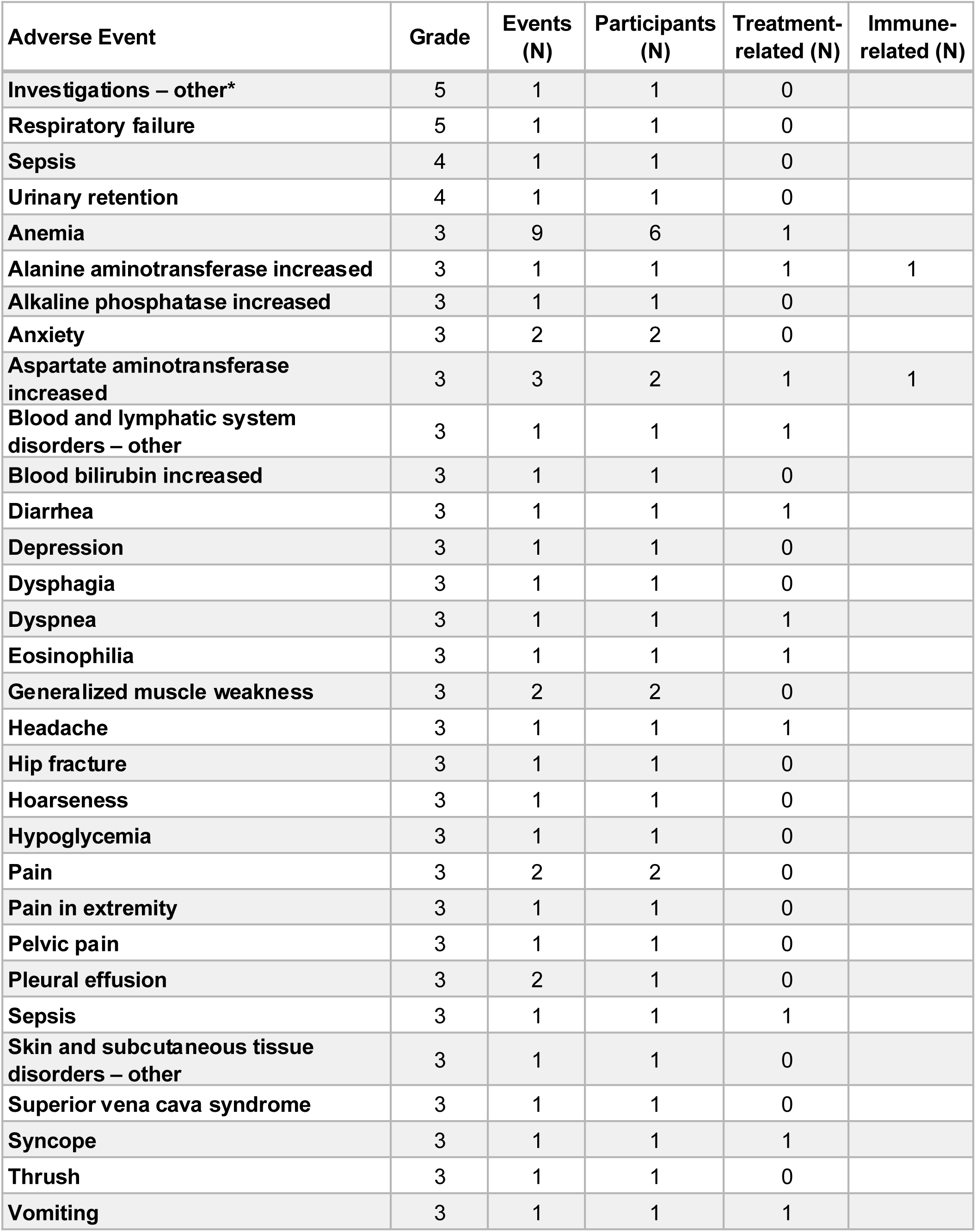
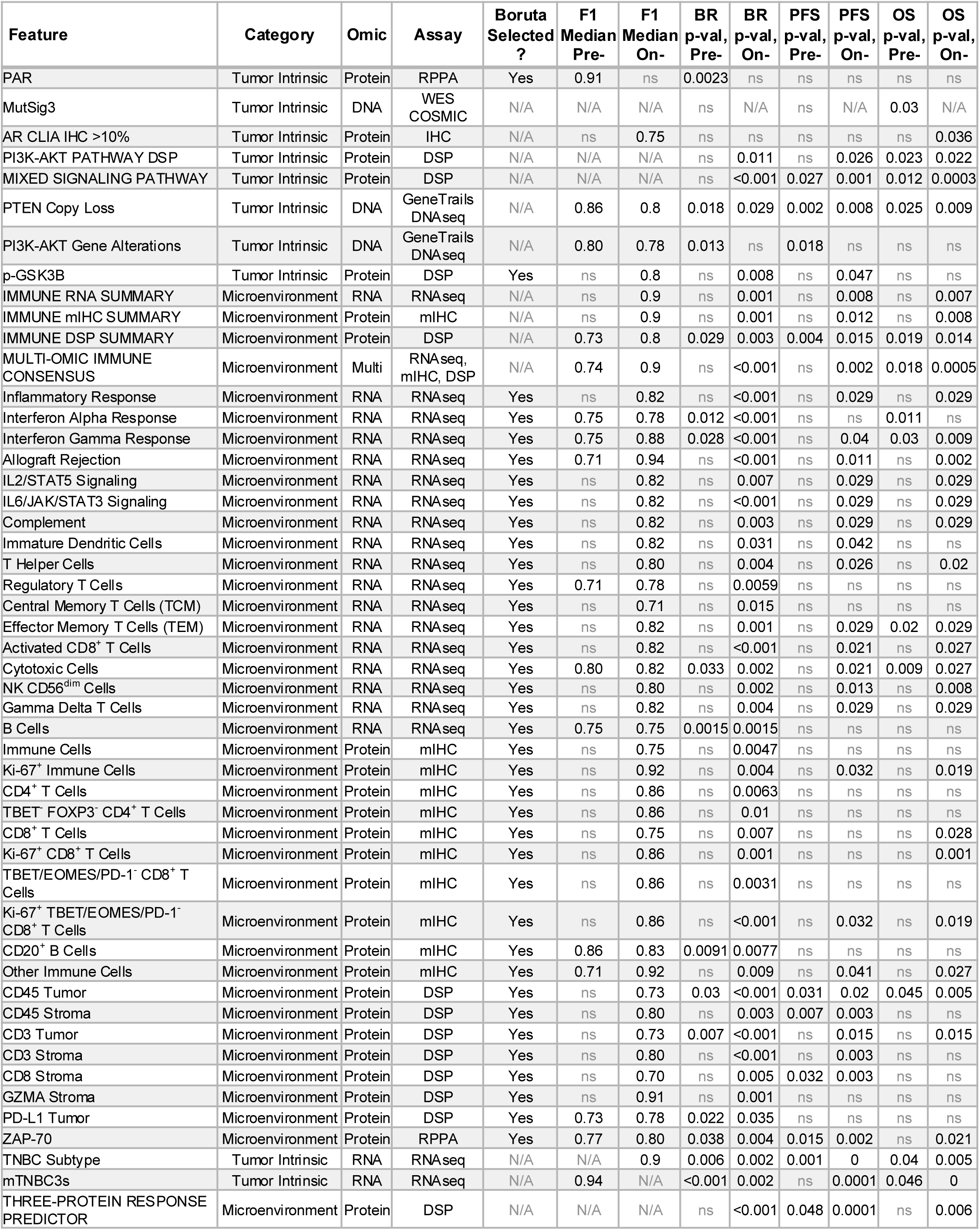
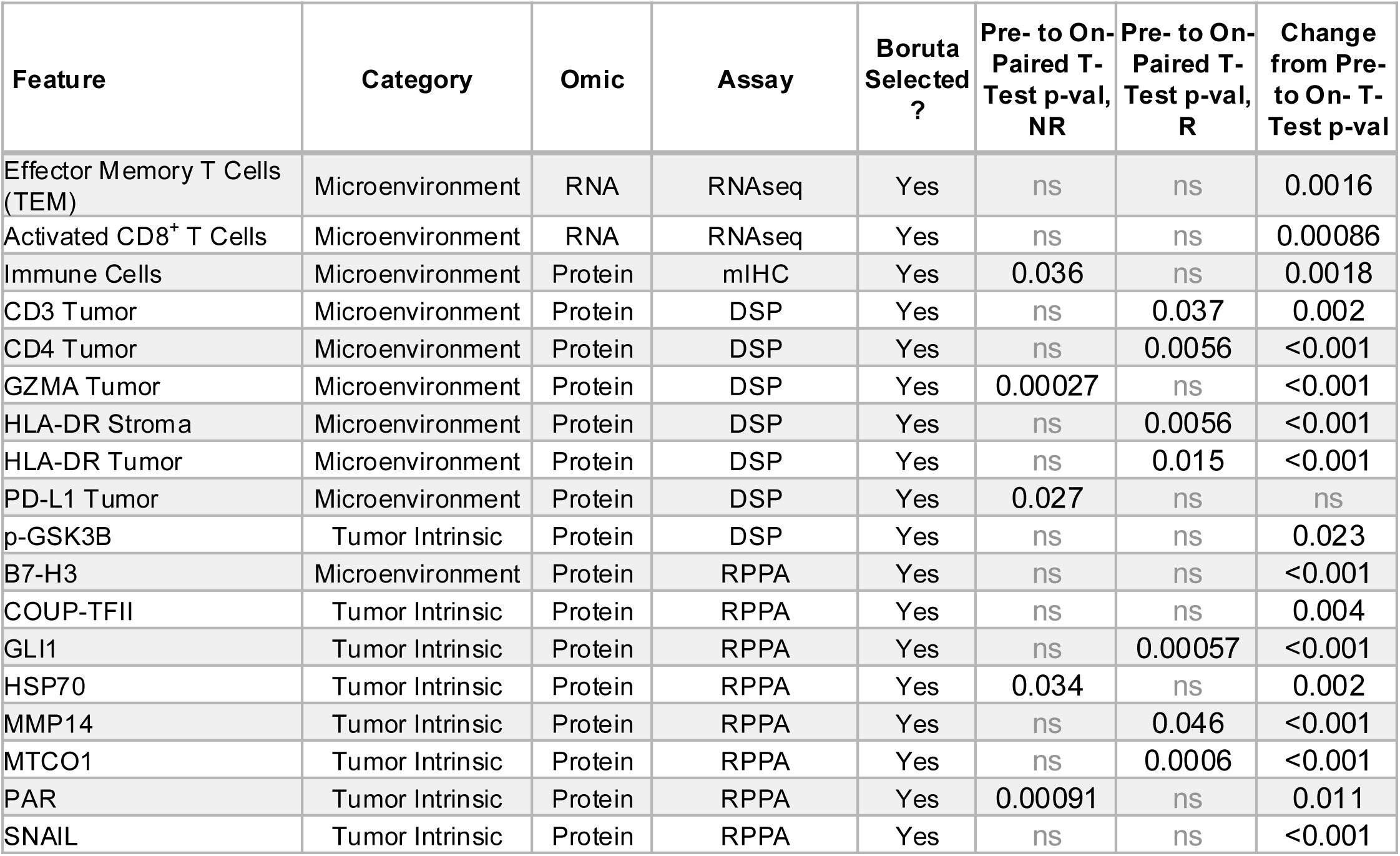
Clinical summary of AMTEC trial participants. **a)** *Adverse events.* Safety analysis performed on data from 24 participants who received at least one dose of either drug. Most treatment-related adverse events (89%) were Grade 1-2. Of the 45 Grade ≥3 events, 20 (44%) were treatment-related. **b)** *Grades 3-5 adverse events.* Data are from 15 of 24 participants. The two immune-related events were resolved with a dose hold. The two Grade 5 events were unrelated to protocol therapy: one due to respiratory failure and one (Investigations – other*) resulting from a motor vehicle accident. **c)** *Response-associated features.* Summary of statistically significant features correlating with response to olaparib and durvalumab. The biopsy in which each feature was observed (Pre-Treatment, Pre-; On-Olaparib, On-) is indicated for significant associations with Best Response (BR), Progression Free Survival (PFS), and Overall Survival (OS). Features identified by Boruta feature selection are noted. Median F1 scores for random training/test split analysis are reported. ns = not significant (p-value >0.05); N/A = not available. **d)** *Olaparib-induced features.* Summary of features significantly changed in olaparib-treated samples. All features were identified by Boruta feature selection performed separately for each dataset: Inferred Immune Cell Type gene sets, mIHC Cell Phenotypes, DSP Signaling or Immune Protein Expression, and RPPA Protein Expression.

